# What is the effectiveness of community diagnostic centres: a rapid review

**DOI:** 10.1101/2022.12.07.22283199

**Authors:** Alesha Wale, Chukwudi Okolie, Jordan Everitt, Amy Hookway, Hannah Shaw, Kirsty Little, Ruth Lewis, Alison Cooper, Adrian Edwards

## Abstract

The COVID-19 pandemic directly impacted diagnostic services in the UK and globally. This exacerbated the rapid rise in demand for diagnostics that existed before the pandemic, resulting in significant numbers of patients requiring various diagnostic services and increased waiting times for diagnostics and treatment. In 2021, community diagnostic centres were launched in England. As diagnostic services account for over 85% of clinical pathways within the NHS and cost over six billion pounds per year, diagnostic centres across a broader range of diagnostic services may be effective, efficient, and cost-effective in the UK health sector.

This rapid review aimed to identify and examine the evidence on the effectiveness of community diagnostic centres. A prior Research Evidence Map was used, along with the stakeholder input, to select a substantive focus for the rapid review. Comparative studies examining community diagnostic centres that accept referrals from primary care as a minimum were included. Prioritised outcomes included those relating to impact on capacity and pressure on secondary care, ensuring equity in uptake or access, and economic outcomes

The review included evidence available up until August 2022. Twenty primary studies were included. Twelve individual diagnostic centres were evaluated across the 20 studies. Most studies evaluated diagnostic centres located within hospital settings. One study evaluated a mobile diagnostic ultrasound service. Most studies were specific to cancer diagnoses. Six studies covered multiple health conditions, which will have also included cancer. Other conditions reported included: severe anaemia, fever of uncertain nature, and multiple sclerosis. A range of outcomes was identified. 11 studies conducted in Spain evaluated the same type of clinic i.e. Quick Diagnostic Unit and seven studies evaluated the same centre at different time intervals. No evidence relating to equity of access was identified.

The evidence relating to effectiveness appeared mixed. There is evidence to suggest that diagnostic centres can reduce various waiting times, including time to surgical consultation, time from consultation to treatment, time from cancer suspicion to treatment, time from diagnosis to specialist consultation and time from diagnosis to treatment. Diagnostic centres could help reduce pressure on secondary care by avoiding hospitalisations in stable patients.

Cost-effectiveness may depend on whether the diagnostic centre is running at full capacity. Factors that could determine the costs incurred by a centre include the diagnostic and clinical complexity of patients, and the characteristics of the unit including the number of staff and contribution of staff time.

## Wales COVID-19 Evidence Centre (WCEC) Rapid Review

### Rapid Review Details

**Review conducted by:** Public Health Wales (PHW)

**Review Team:**

- Alesha Wale, Public Health Wales,
- Chukwudi Okolie, Public Health Wales,
- Jordan Everitt, Public Health Wales,
- Amy Hookway, Public Health Wales,
- Hannah Shaw, Public Health Wales,
- Kirsty Little, Public Health Wales

**Review submitted to the WCEC:** November 2022

**Stakeholder consultation meeting**: 22nd November 2023

**Rapid Review report issued by the WCEC:** December 2022

### WCEC Team

Ruth Lewis, Adrian Edwards, Alison Cooper involved in drafting the Topline Summary and editing.

**This review should be cited as:** RR00043_Wales COVID-19 Evidence Centre_What is the effectiveness of community diagnostic centres: a rapid review. November 2022

**Disclaimer:** The views expressed in this publication are those of the authors, not necessarily Health and Care Research Wales. The WCEC and authors of this work declare that they have no conflict of interest.

### TOPLINE SUMMARY

#### What is a Rapid Review?

Our rapid reviews (RR) use a variation of the systematic review approach, abbreviating or omitting some components to generate the evidence to inform stakeholders promptly whilst maintaining attention to bias. They follow the methodological recommendations and minimum standards for conducting and reporting rapid reviews, including a structured protocol, systematic search, screening, data extraction, critical appraisal, and evidence synthesis to answer a specific question and identify key research gaps. They take 1-2 months, depending on the breadth and complexity of the research topic/ question(s), extent of the evidence base, and type of analysis required for synthesis.

This report is linked to a **rapid evidence map** published as: REM00043_ Wales COVID-19 Evidence Centre. A rapid evidence map of what evidence is available on the effectiveness of community diagnostic centres. September 2022

#### Who is this summary for?

Diagnostics Strategy Board

#### Background / Aim of Rapid Review

The COVID-19 pandemic directly impacted diagnostic services in the UK and globally. This exacerbated the rapid rise in demand for diagnostics that existed before the pandemic, resulting in **significant numbers of patients requiring various diagnostic services and increased waiting times for diagnostics** and treatment. **In 2021, community diagnostic centres were launched in England.** As diagnostic services account for over 85% of clinical pathways within the NHS and cost over £6 billion per year, **diagnostic centres across a broader range of diagnostic services may be effective, efficient, and cost-effective** in the UK health sector.

This rapid review aimed **to identify and examine the evidence on the effectiveness of community diagnostic centres.** The prior REM was used, along with the stakeholder input, to select a substantive focus for the RR. It was decided that only **comparative studies** examining **community diagnostic centres that accept referrals from primary care as a minimum** would be included. Prioritised **outcomes included those relating to impact on capacity and pressure on secondary care, ensuring equity in uptake or access, and economic outcomes**.

#### Key Findings

##### Extent of the evidence base

- Twenty **primary studies** were included: 16 quasi-experimental studies (all comparative studies using cross sectional post-test only designs), three economic evaluations and one randomised controlled trial (the latter published in 1998, and now superseded by more recent research).
- Twelve individual diagnostic centres were evaluated across the 20 studies.
- Most studies (n=19) evaluated diagnostic centres **located within hospital settings**. One study evaluated a mobile diagnostic ultrasound service.
- Most studies (n=10) were specific to **cancer diagnoses.** Six studies covered **multiple health conditions,** which will have also included cancer. Other conditions reported included: severe anaemia (n=1), fever of uncertain nature (n=1), and multiple sclerosis (n=1). One study did not report a particular health condition of interest.
- A range of outcomes was identified from studies conducted in Canada (n=4), UK (n=5), and Spain (n=11). The 11 studies conducted in Spain evaluated the same type of clinic – Quick Diagnostic Unit (QDU), and seven of these studies evaluated the same centre at different time intervals.
- **No evidence relating to equity of access** was identified.
- **No comparative ongoing studies** were identified.

##### Recency of the evidence base

The review included evidence available up until August 2022. Included studies were published between 1998 and 2021, with data collection between 1995 and 2018.

##### Evidence of effectiveness

- The evidence relating to effectiveness appeared mixed.
- There is evidence to suggest that diagnostic centres can reduce various waiting times, **including time to surgical consultation, time from consultation to treatment, time from cancer suspicion to treatment, time from diagnosis to specialist consultation** and **time from diagnosis to treatment**.
- Diagnostic centres could help reduce pressure on secondary care by **avoiding hospitalisations in stable patients, reducing the number of visits required to receive a definite diagnosis, and increasing the number of patients in whom a definite outcome (discharged or scheduled for surgery) was reached**.
- Cost-effectiveness may depend on whether the diagnostic centre is running at full capacity. Factors that could determine the costs incurred by a diagnostic centre include **the diagnostic and clinical complexity of patients, and the characteristics of the unit including the number of staff and contribution of staff time**.

##### Best quality evidence

- All studies had considerable methodological limitations. The three economic evaluation studies (Bosch et al 2021, Sewell et al 2020, Sanclemente-Ansó et al 2016) were considered the most robust.

#### Policy Implications

- This rapid review highlighted **possible benefits of diagnostic centres**, particularly with regards to **reducing waiting times and pressure on hospitals**.
- As the data collection dates of included studies are wide-ranging, and many of the diagnostic centres included were from other countries where the healthcare system is different to that of the UK, **the results may not be generalisable**.
- Further research is needed to determine the optimum location for diagnostic centres.
- Further research is required to evaluate the effectiveness of diagnostic centres for conditions other than cancer.
- Further well-designed robust research from the UK and other comparable countries is needed to better understand the effectiveness and accessibility of diagnostic centres within Wales. (Only one UK study was published after 2013)
- Comparative impact evaluations should be incorporated into service development plans from the onset, to assess the effectiveness of newly opened diagnostic centres in the UK over time.

#### Strength of Evidence

Most study designs used weak methods that may be less appropriate for inferring effectiveness. Studies varied by countries, designs, definitions and often had inconsistent findings, so results should be interpreted with caution.

## 1. BACKGROUND

### 1.1 Who is this review for?

This Rapid Review (RR) was conducted as part of the Wales COVID-19 Evidence Centre Work Programme (WCEC). The above question was suggested by the Welsh Government Technical Advisory Cell (TAC) to inform the Diagnostics Strategy Board and support the implementation of community diagnostic centres across Wales.

### 1.2 Purpose of this review

The COVID-19 pandemic has had a direct impact on diagnostic services in the United Kingdom (UK) and globally. This, in addition to the rapid rise in demand for diagnostics that existed prior to the pandemic, has resulted in a significant backlog of patients requiring various diagnostic services and increased waiting times for diagnostics and treatment. The most recently published data shows that in Wales, the number of patients waiting longer than the target of eight weeks for diagnostics rose from 10.8% (7,964) in March 2020 to 41.5% (44,489) in August 2022 (StatsWales 2022). An Independent Review of Diagnostic Services for NHS England chaired by Professor Sir Mike Richards, called for significant reform and investment in diagnostic services, and recommended the establishment of community diagnostic centres to aid in tackling the backlog and delays to diagnostic services (National Health Service England 2020). Community diagnostic centres aim to provide patients with quicker and more convenient direct access to diagnostic services than is currently available, and reduce pressure on hospitals (Department of Health and Social Care 2021). With an emphasis on direct patient access to services from primary care, these centres can be located within hospital settings or within the community. In Wales, community diagnostic centres are generally referred to as **Regional Diagnostic Hubs** (but are referred to here as Community Diagnostic Centres, for the purpose of this report).

In England, community diagnostic centres were first launched in 2021 in a range of settings including hospitals, football stadiums, and repurposed retail outlets (Department of Health and Social Care 2021). At present, over 90 community diagnostic centres have been opened, with plans to open up to 160 centres by 2025 (National Health Service England 2022). In Wales, a plan to create a network of community diagnostic centres was outlined by the Welsh Government in April 2022 (Welsh Government 2022). As diagnostic services currently account for over 85% of clinical pathways within the NHS and cost over £6 billion a year (National Health Service 2022), community diagnostic centres across a broader range of diagnostic services may be an effective, efficient, and cost-effective introduction to the UK health sector. These services could ensure timely diagnoses and reduced waiting times in a convenient location, ensuring people receive the treatment they need. Furthermore, community diagnostic centres could help address inequalities by providing accessible diagnostic services to people from particular social groups who may be less likely to engage with the healthcare system (The King’s Fund 2022).

The purpose of this RR is to draw on the earlier work undertaken to identify and examine the evidence on the effectiveness of community diagnostic centres. This preliminary work included a rapid evidence map (REM) that identified a large body of evidence relevant to community diagnostic centres: REM00043_ Wales COVID-19 Evidence Centre. A rapid evidence map of what evidence is available on the effectiveness of community diagnostic centres. September 2022. In order to measure effectiveness as part of the current RR, stakeholders prioritised outcomes, identified during the REM, that evaluated whether community diagnostic centres can impact capacity and pressure on secondary care, as well as ensuring equity in uptake or access as most important, along with economic outcomes. Therefore, our RR focussed on outcomes that were best able to demonstrate this. Due to the large number of studies identified during the initial investigation into this topic, **only comparative studies examining community diagnostic centres that accept referrals from primary care as a minimum, were included in this RR**. Comparative studies are defined here as studies that investigated some aspect of a diagnostic centre and compared it with usual care, another diagnostic centre or some other service.

### 1.3 Definition of community diagnostic centres

Community diagnostic centres are described within the international literature using a variety of terms and definitions. For the purposes of this RR, community diagnostic centres are defined as health services aimed at improving population health outcomes by providing quicker and easily accessible diagnostic services in the community, which are accessible to primary care practitioners/services, thereby relieving pressure on secondary care services. In Wales, community diagnostic centres are generally referred to as Regional Diagnostic Hubs to avoid confusion with the descriptors or acronyms used for other similar services. For the purposes of this RR, we use the descriptor ‘community diagnostic centres’ to incorporate the range of terms used for these services. However, the different descriptors used within individual studies are also outlined and discussed in this report.

## 2. RESULTS

### 2.1 Overview of the Evidence Base

A total of 20 primary studies met our inclusion criteria. Sixteen of these were quasi-experimental studies comprising cross sectional post-test only designs, three were economic evaluations and one was a randomised controlled trial (RCT). Included studies were conducted in Spain (n=11), UK (n=5), and Canada (n=4), and were published between 1998 and 2021. Ten studies were specific to cancer diagnoses, six studies reported on multiple health conditions (rather than a specific condition) and three studies covered a single health condition including severe anaemia (n=1), fever of uncertain nature (FUN) (n=1), and multiple sclerosis (MS) (n=1). One study did not report a particular health condition of interest. Included studies varied in their reporting of outcomes, with some being purely descriptive and others offering inferential statistical findings. A detailed matrix of the outcomes reported by each study presented in this RR can be found in Figure 1. A detailed summary of included studies organised by country, diagnostic centre and comparator, can be found in Table 1.

**Figure 1.**
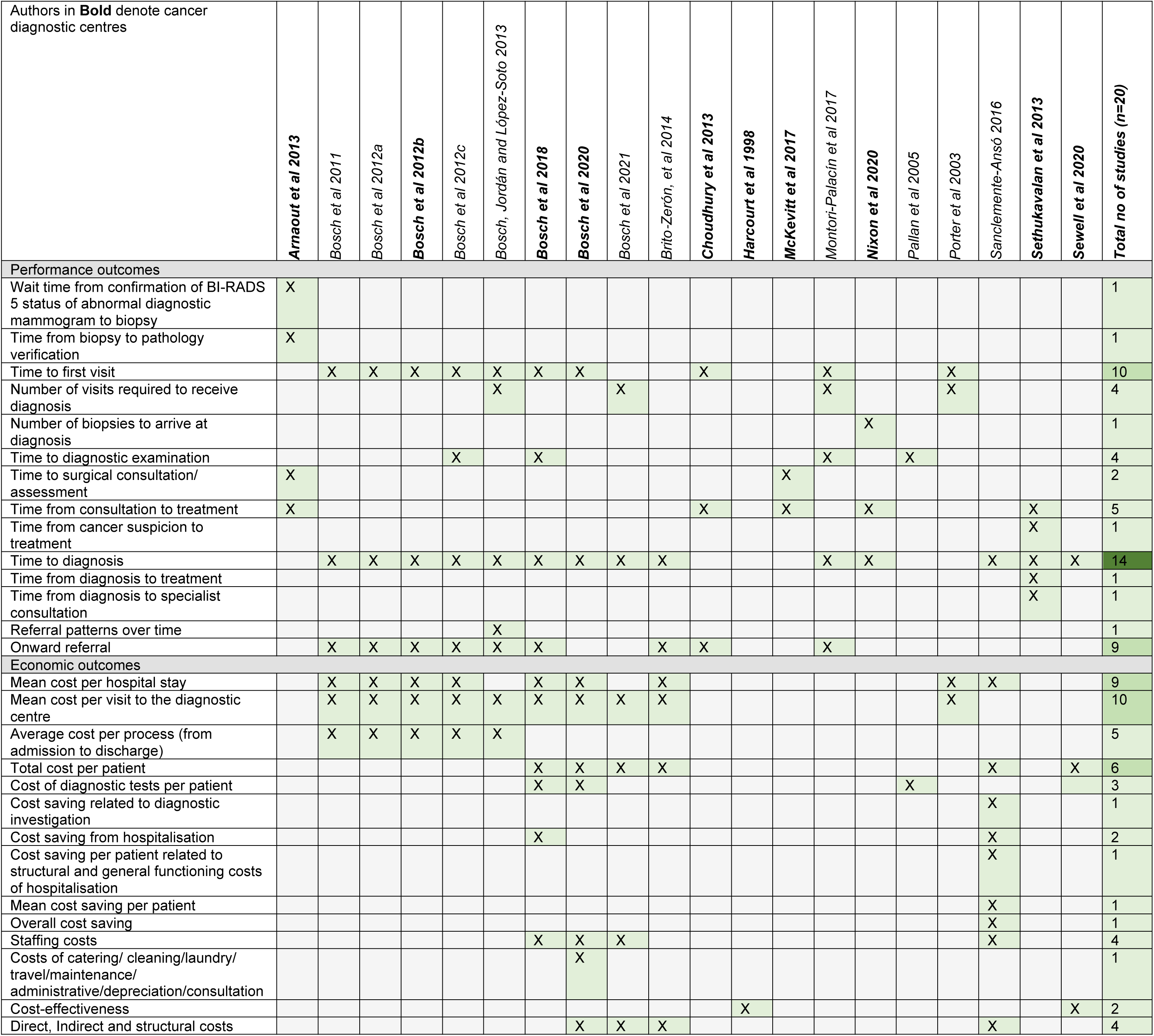
Outcomes matrix of included studies as reported in the RR (all outcomes reported by each study can be seen in the rapid evidence map (REM)).

**Table 1:**
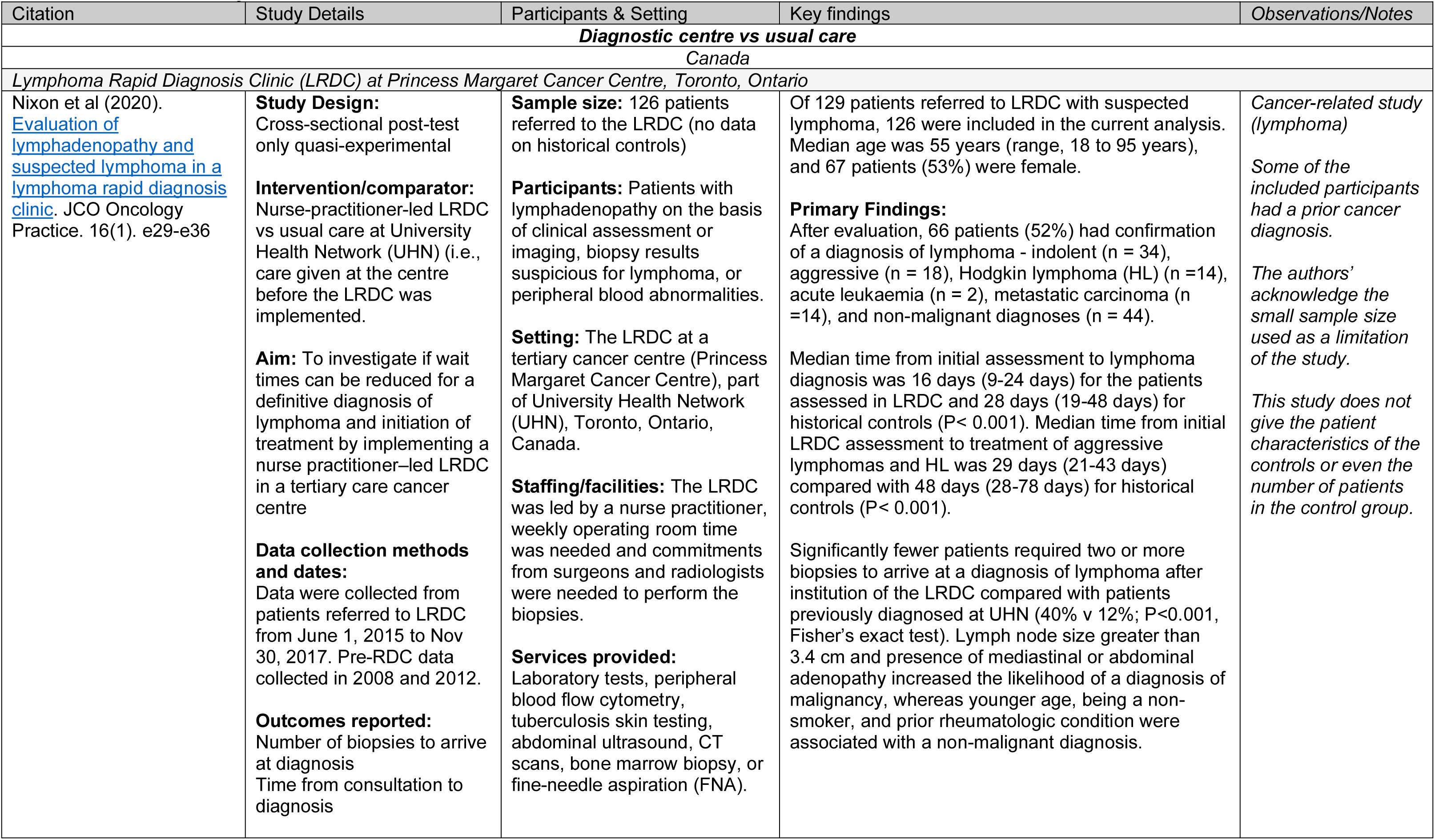
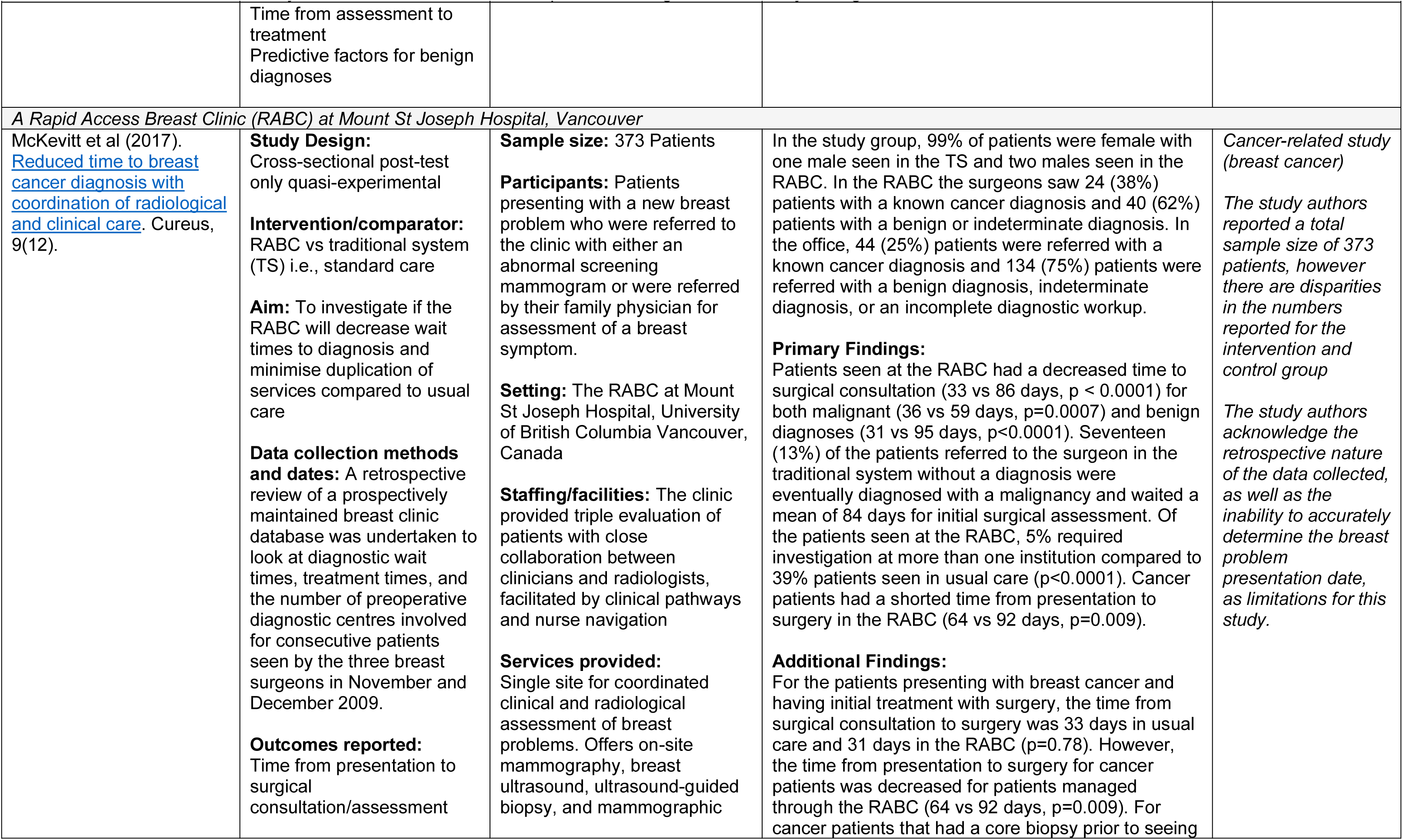
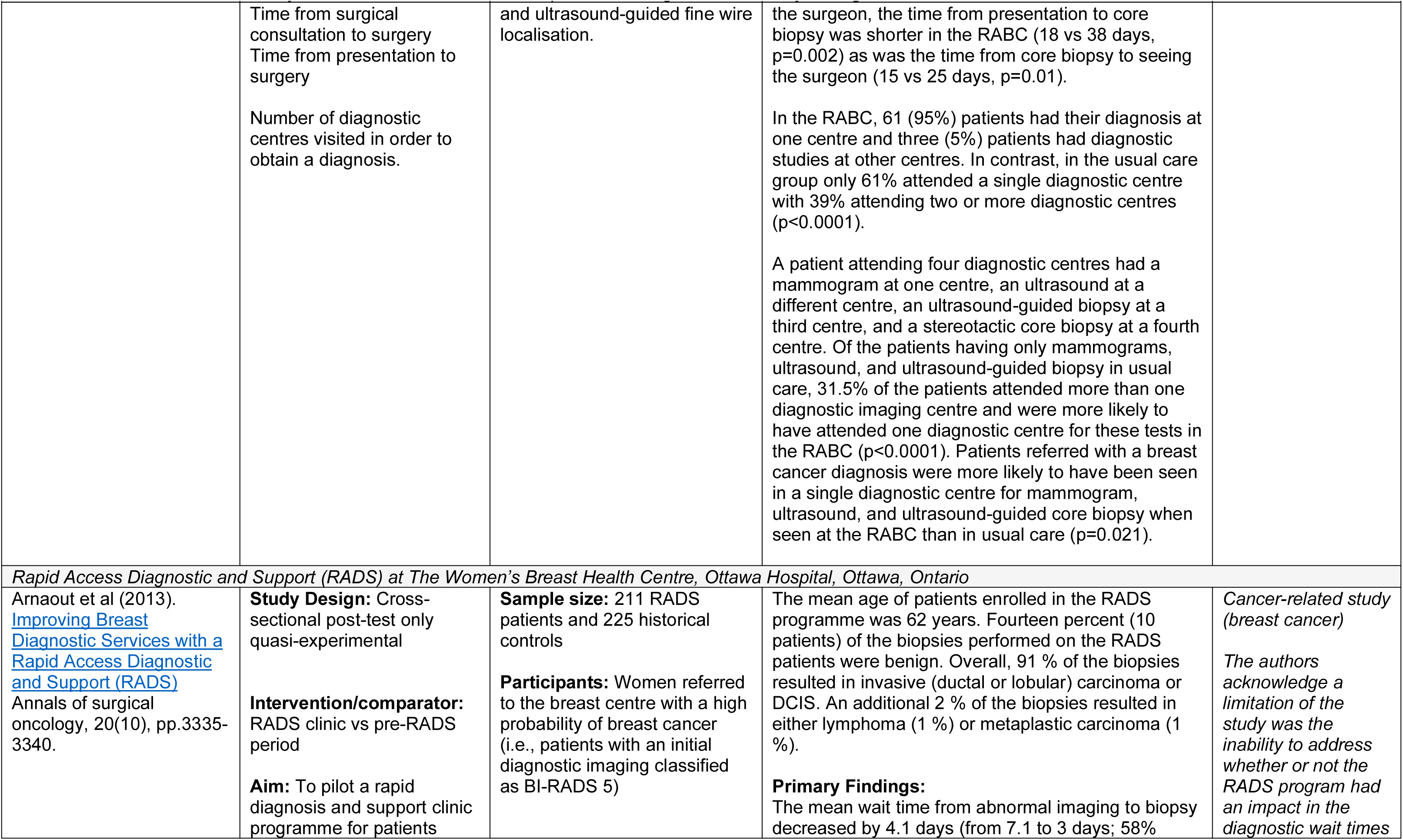
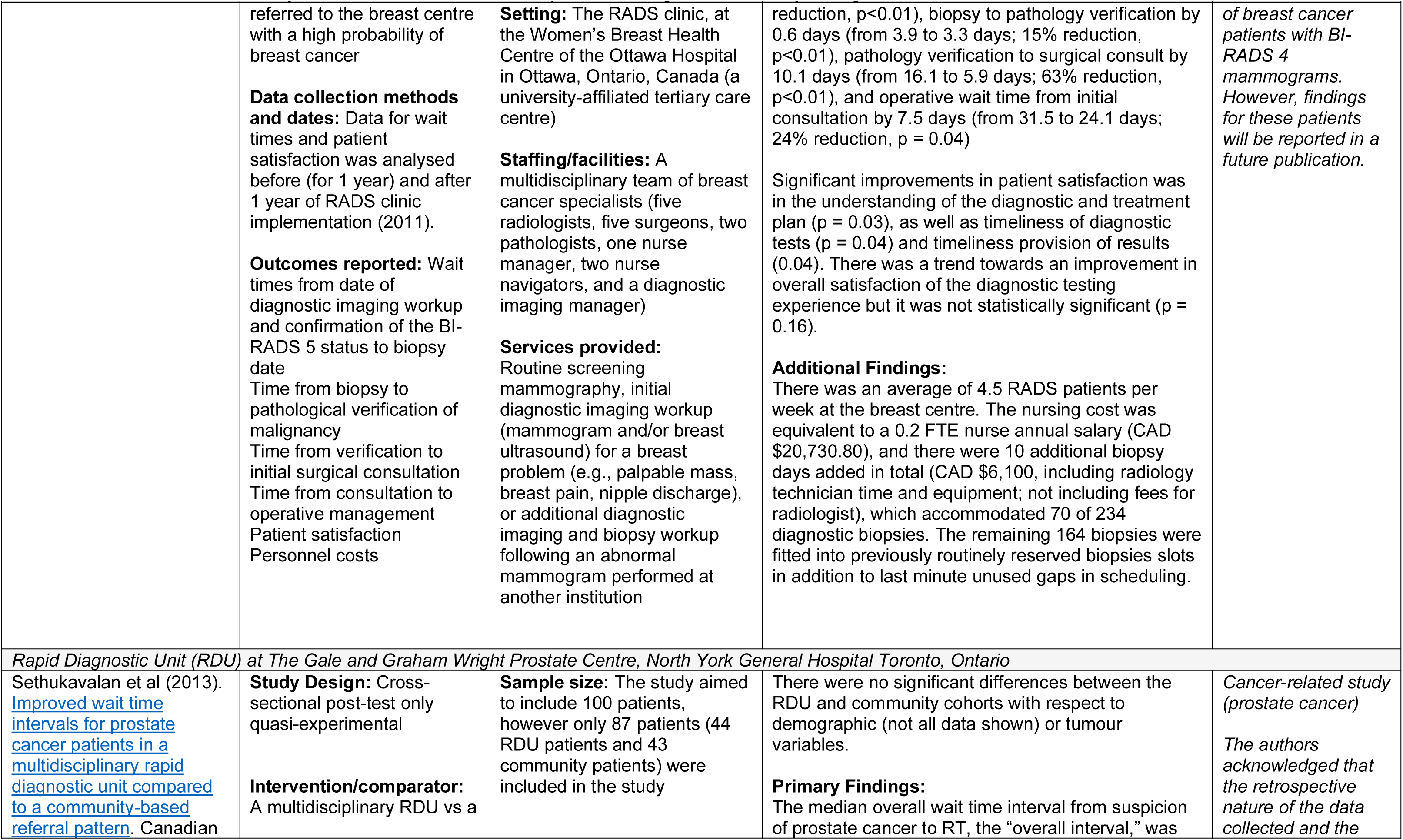
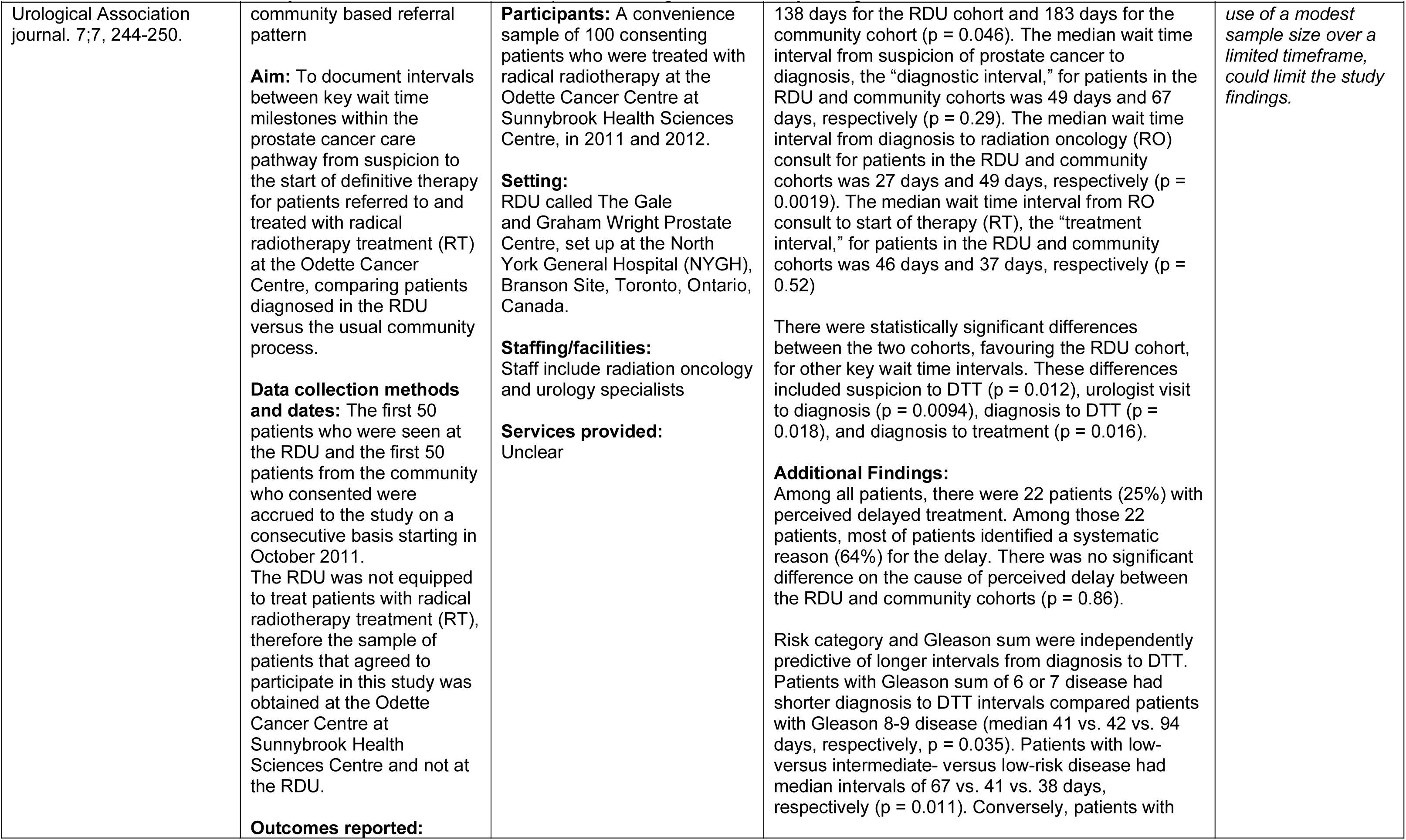
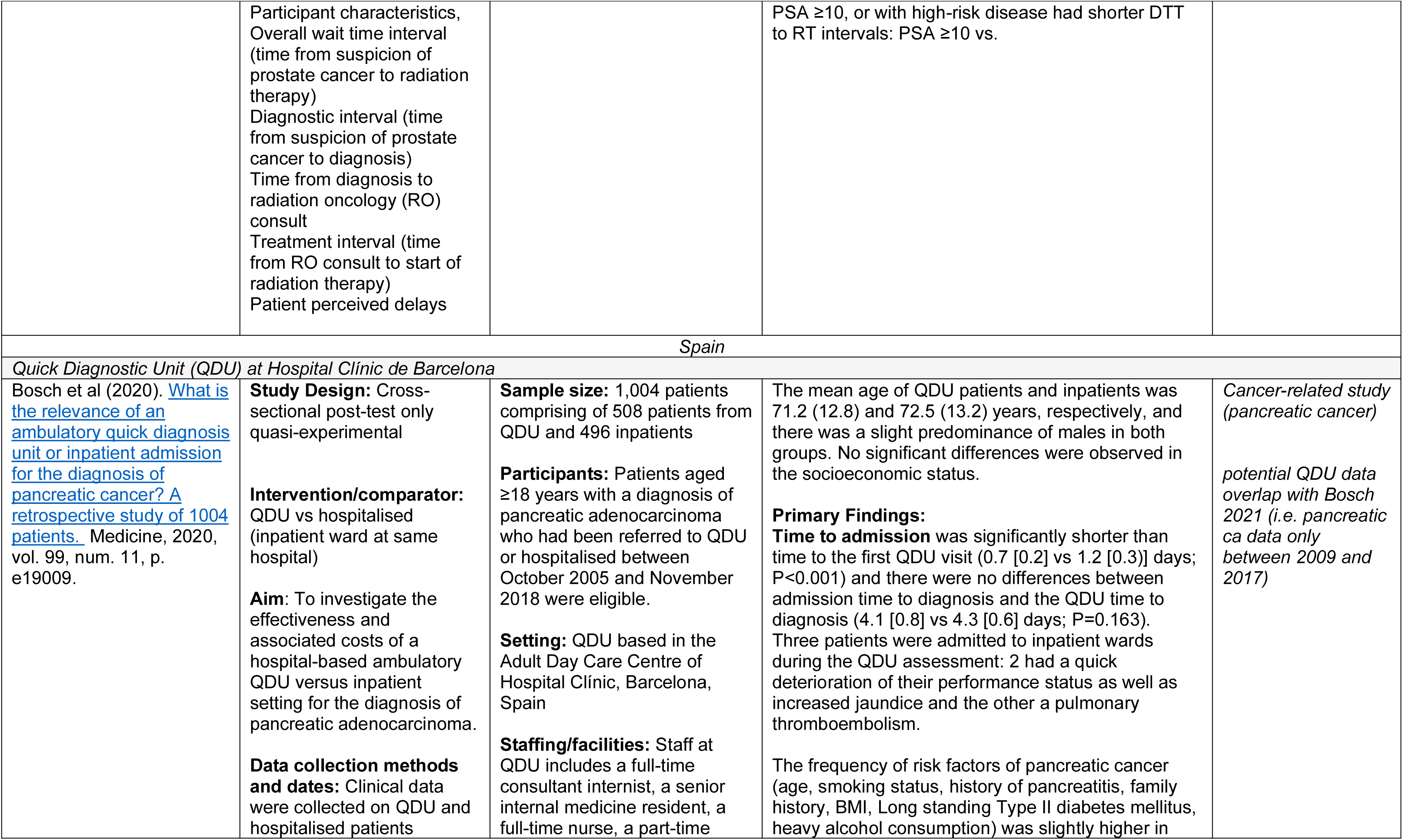
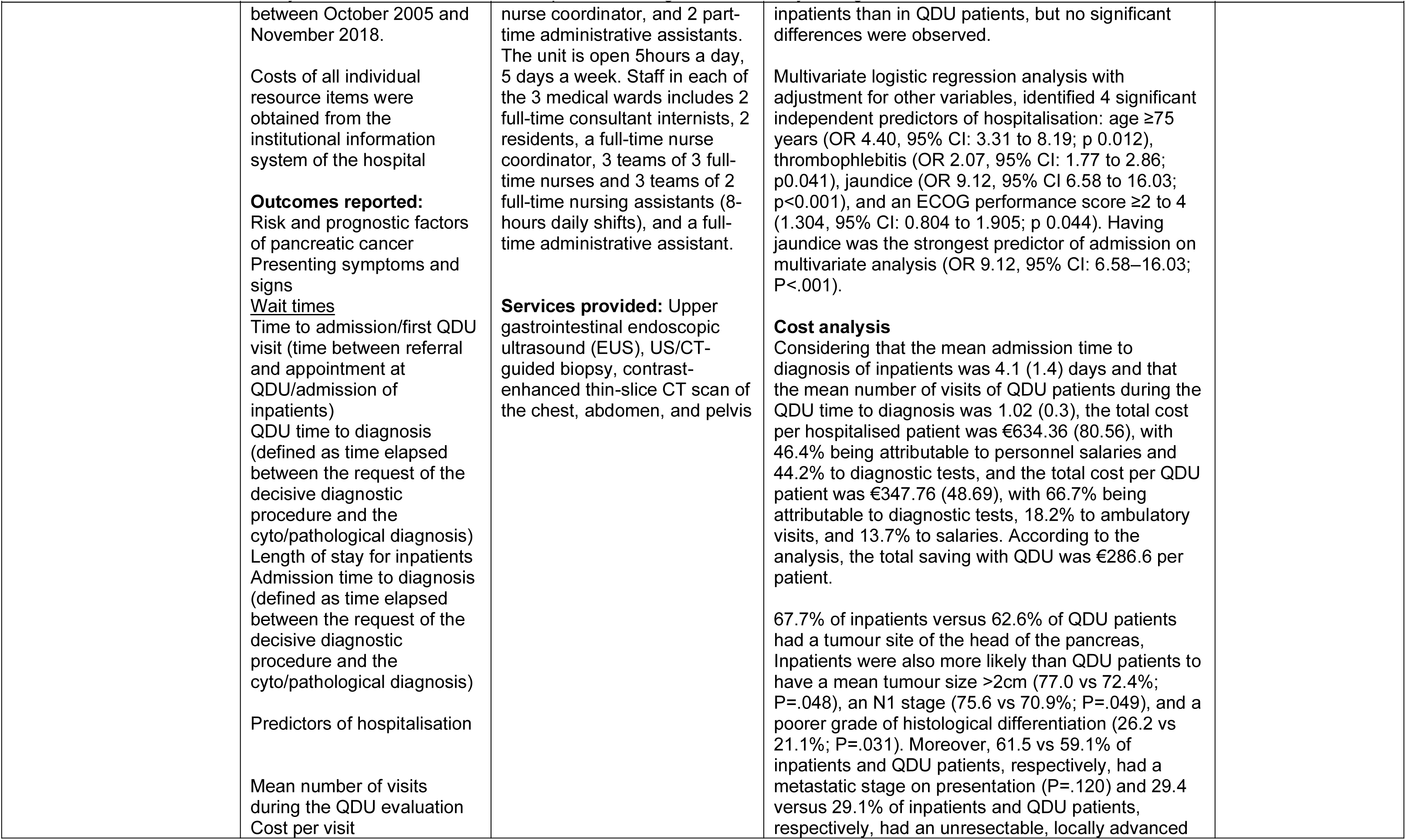
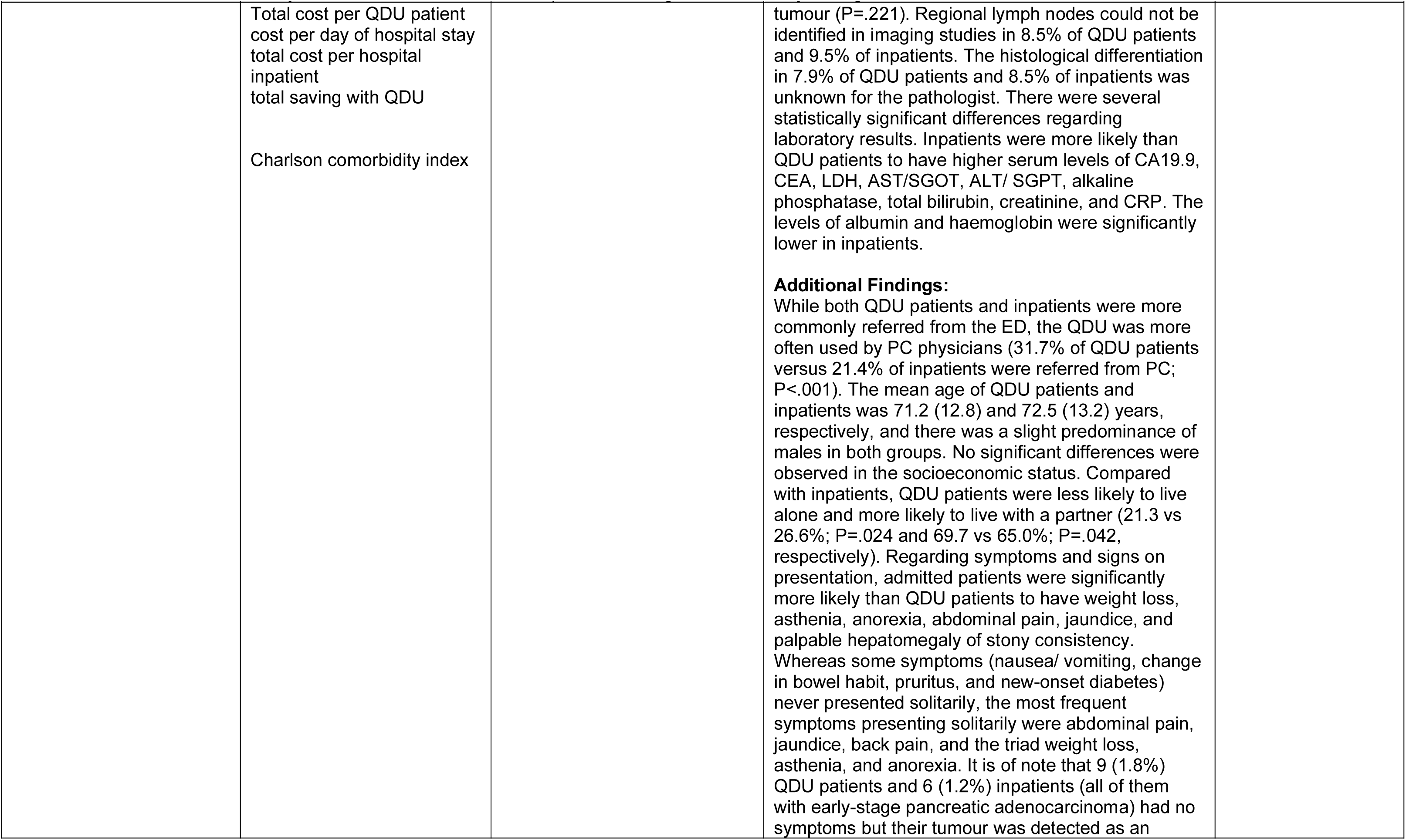
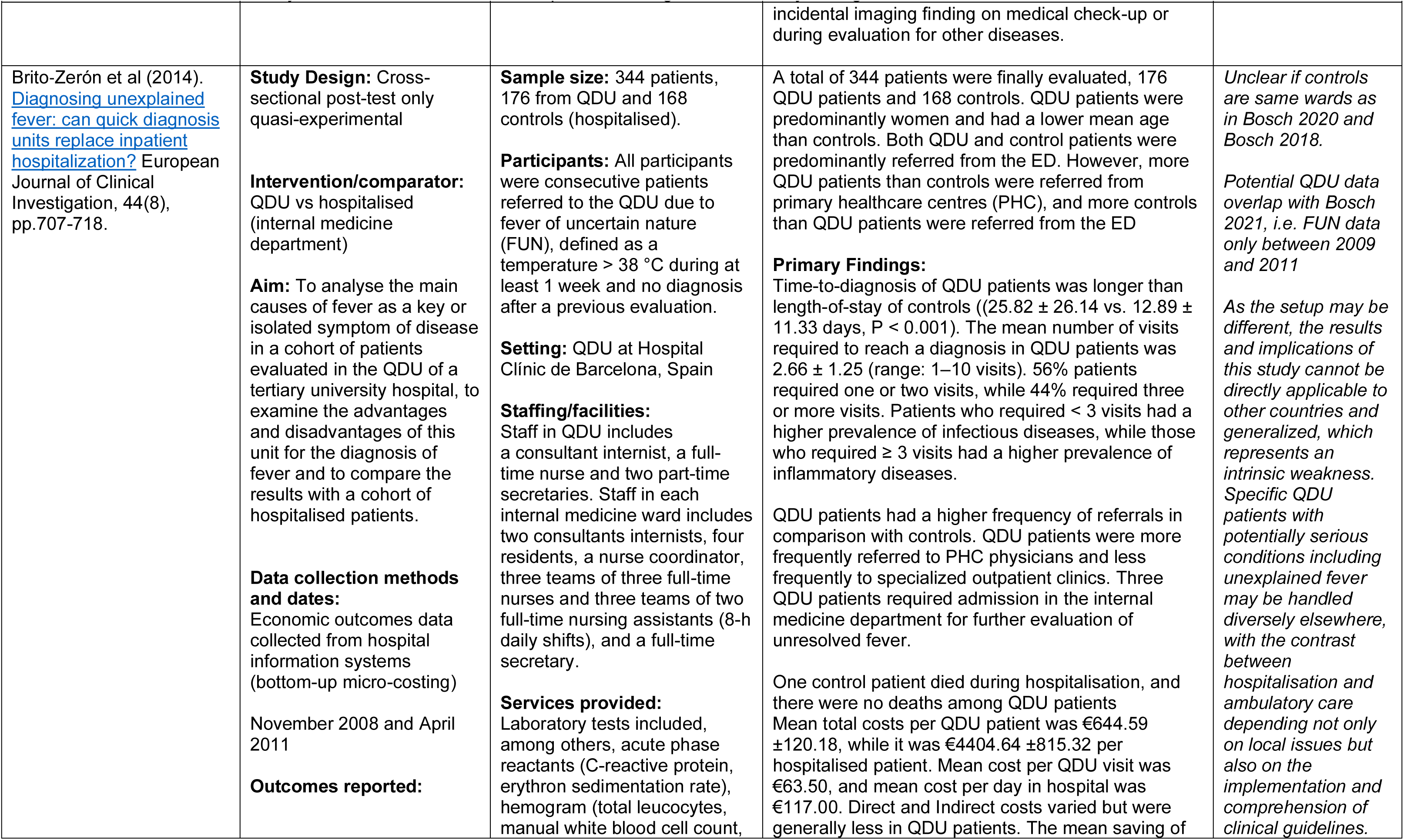
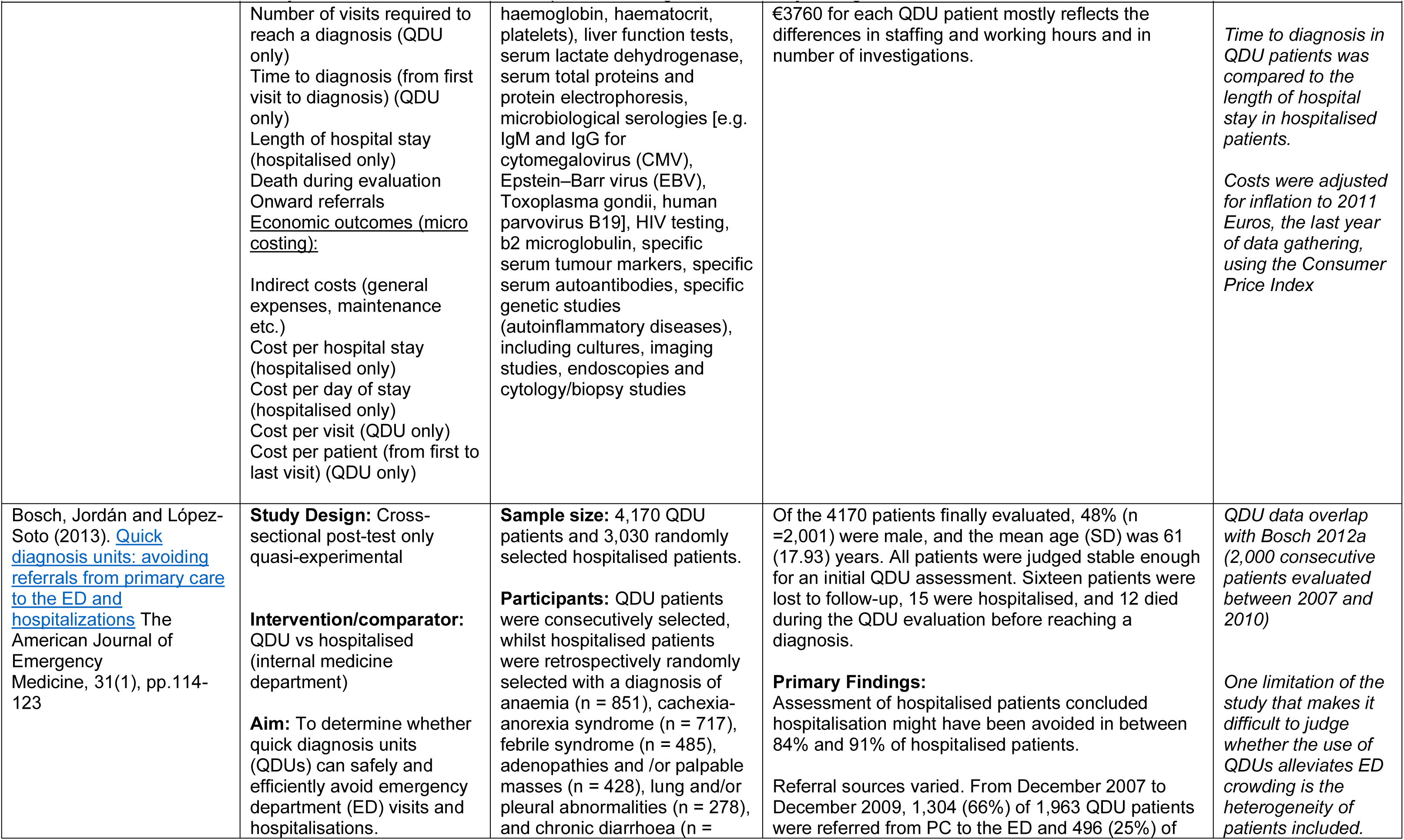
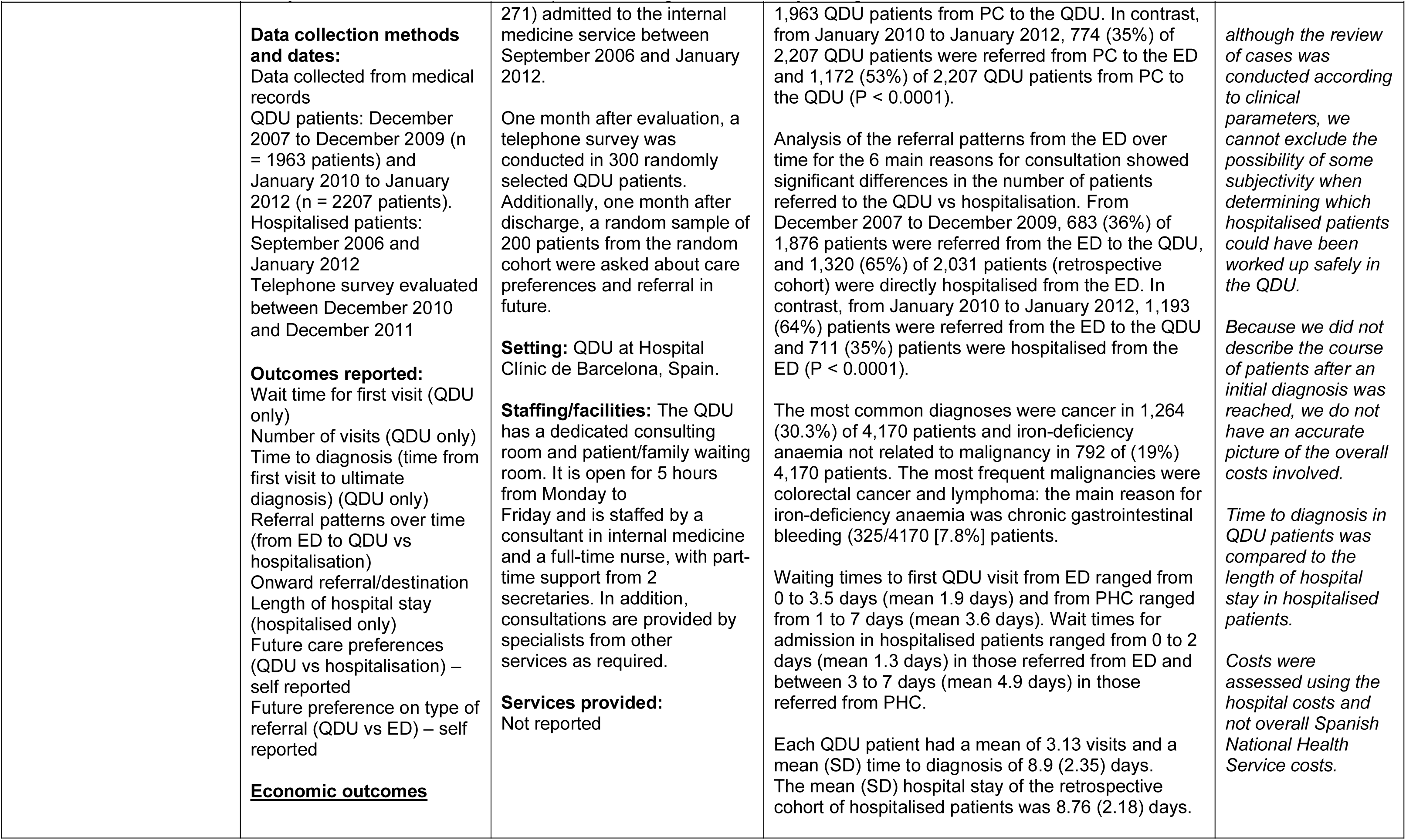
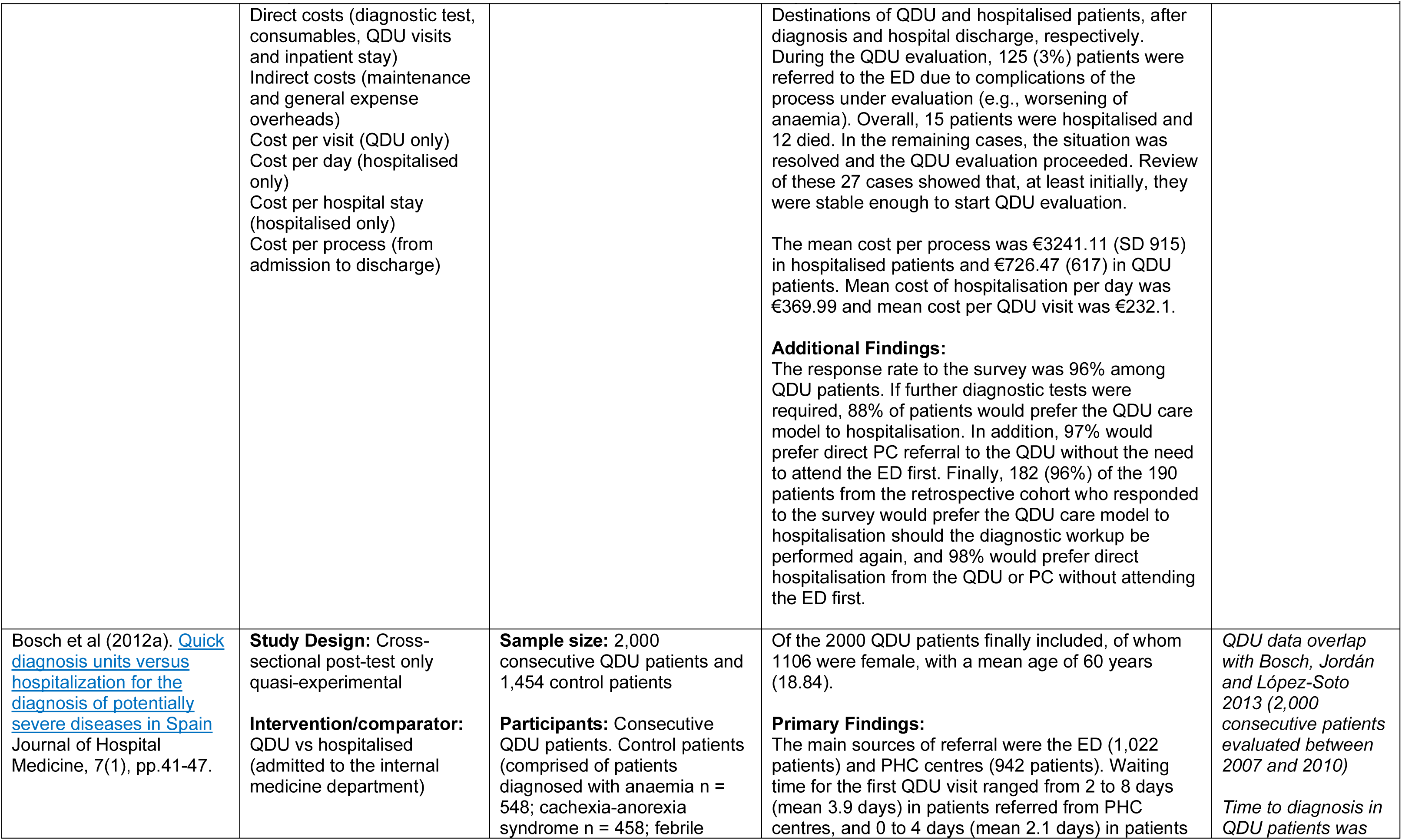
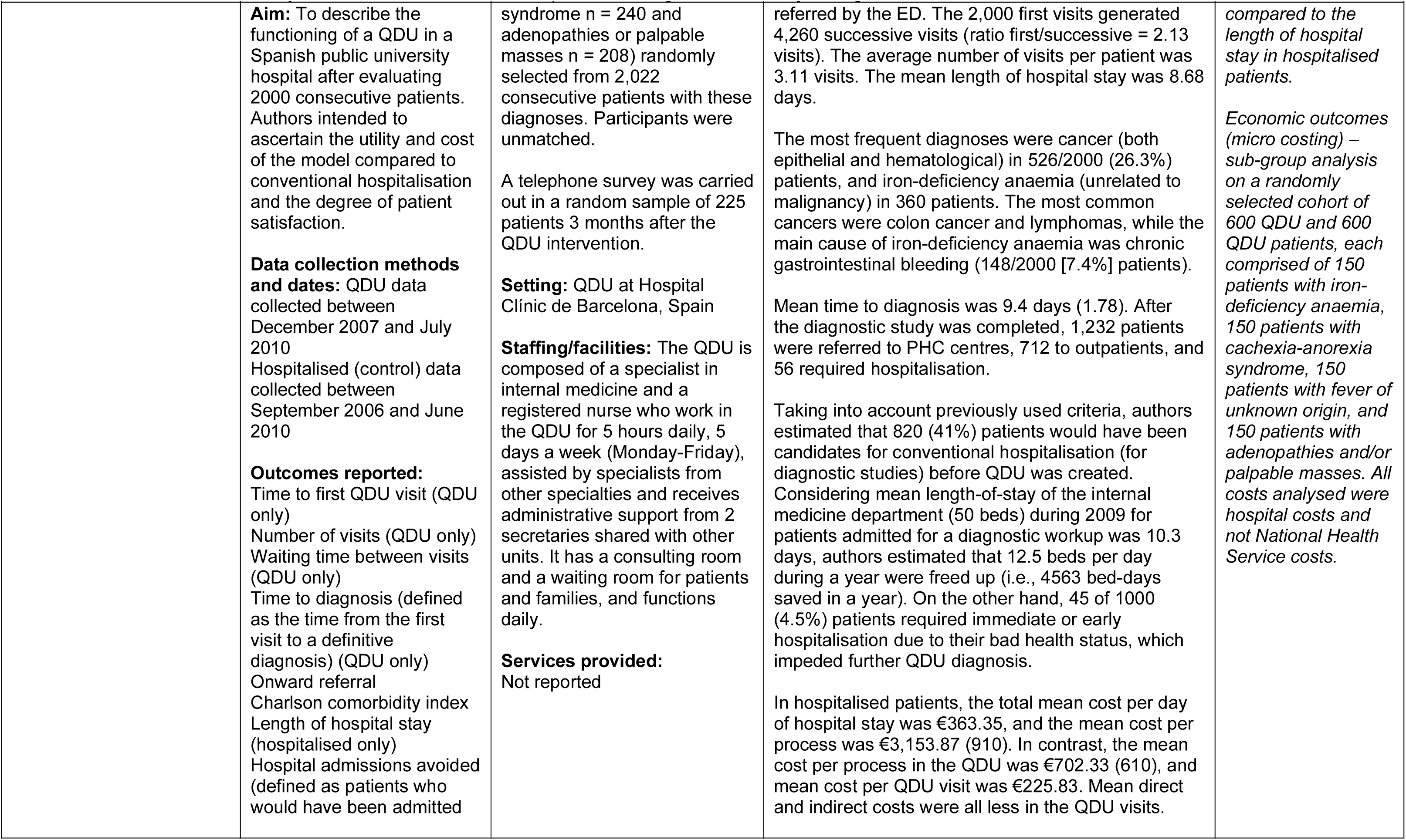
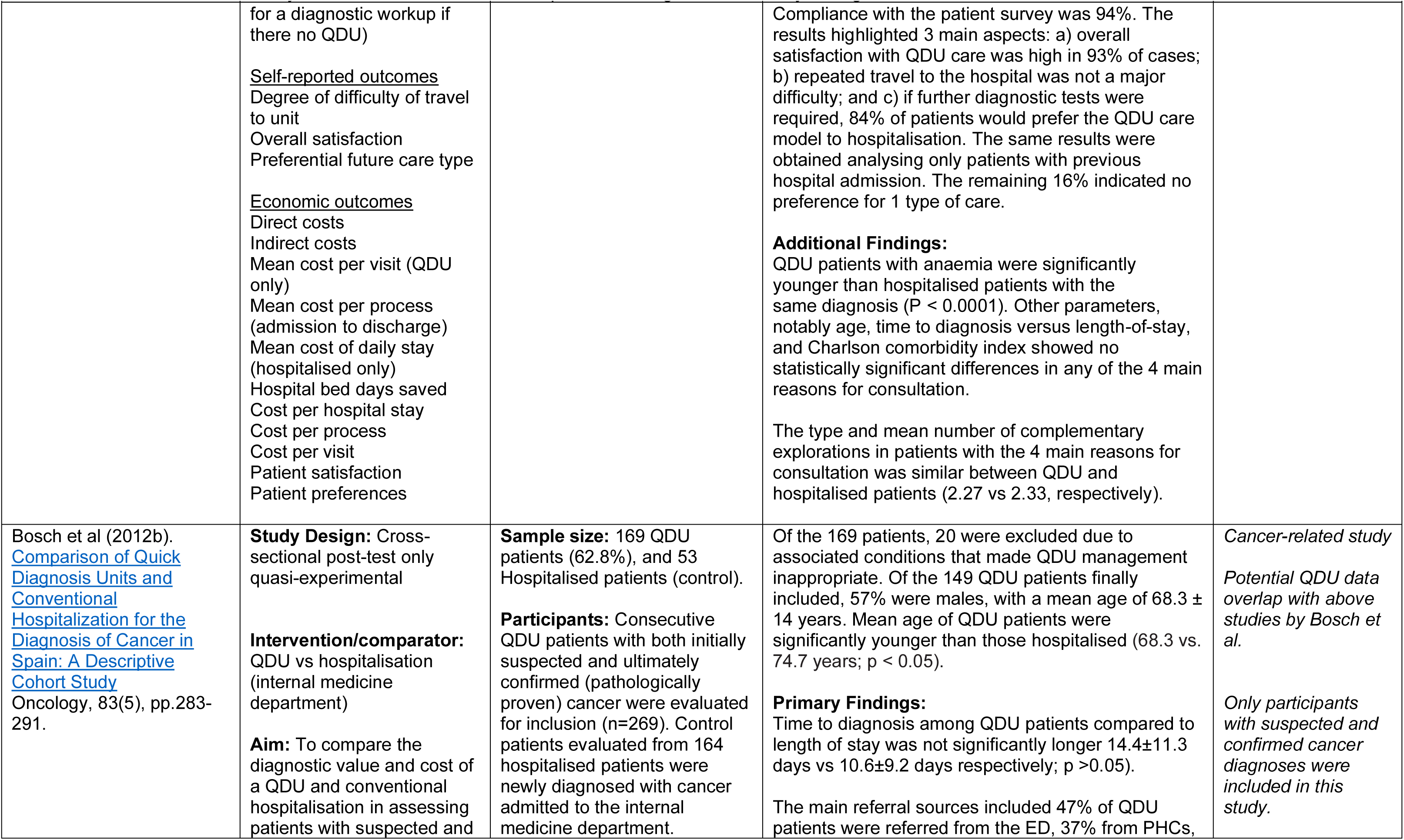
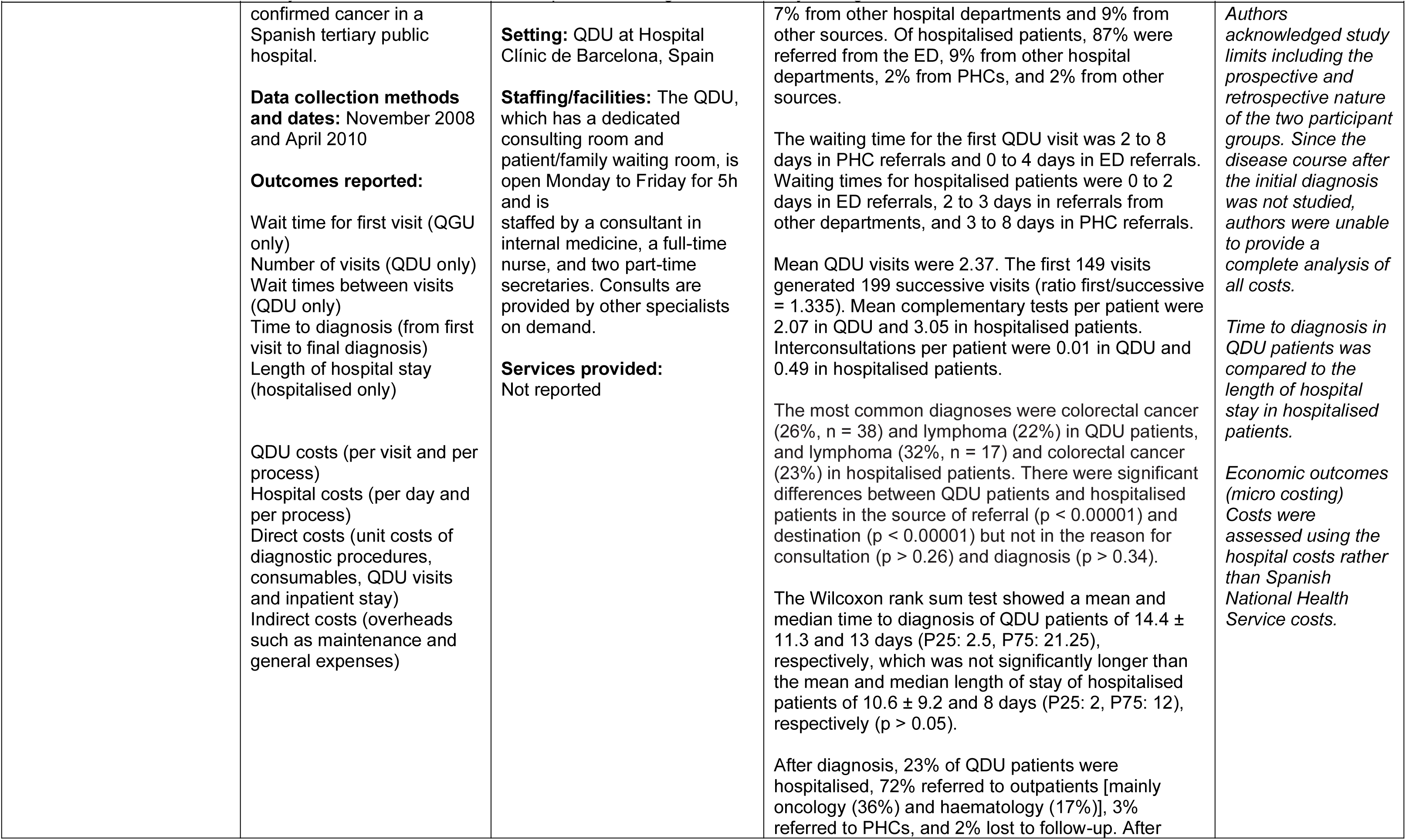
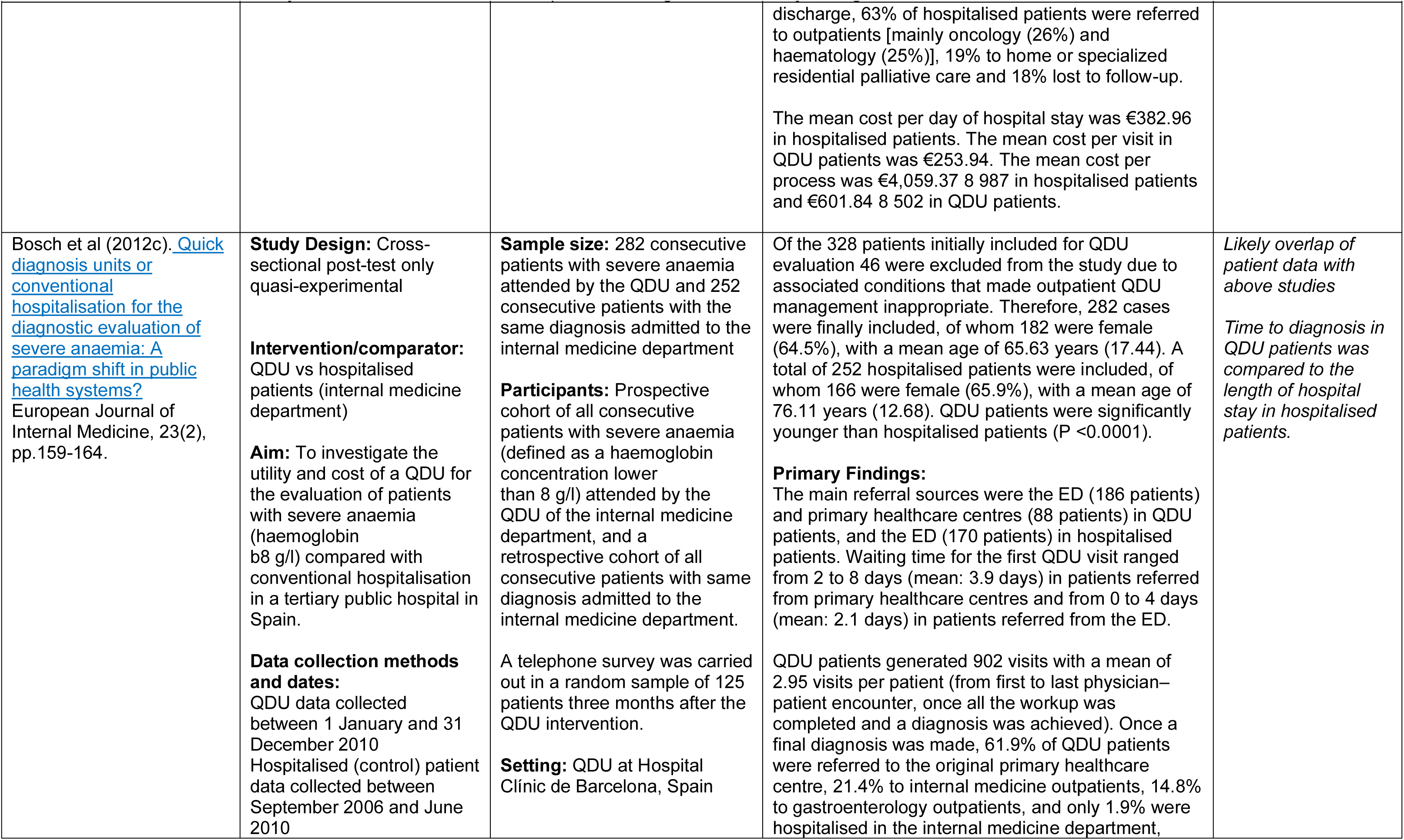
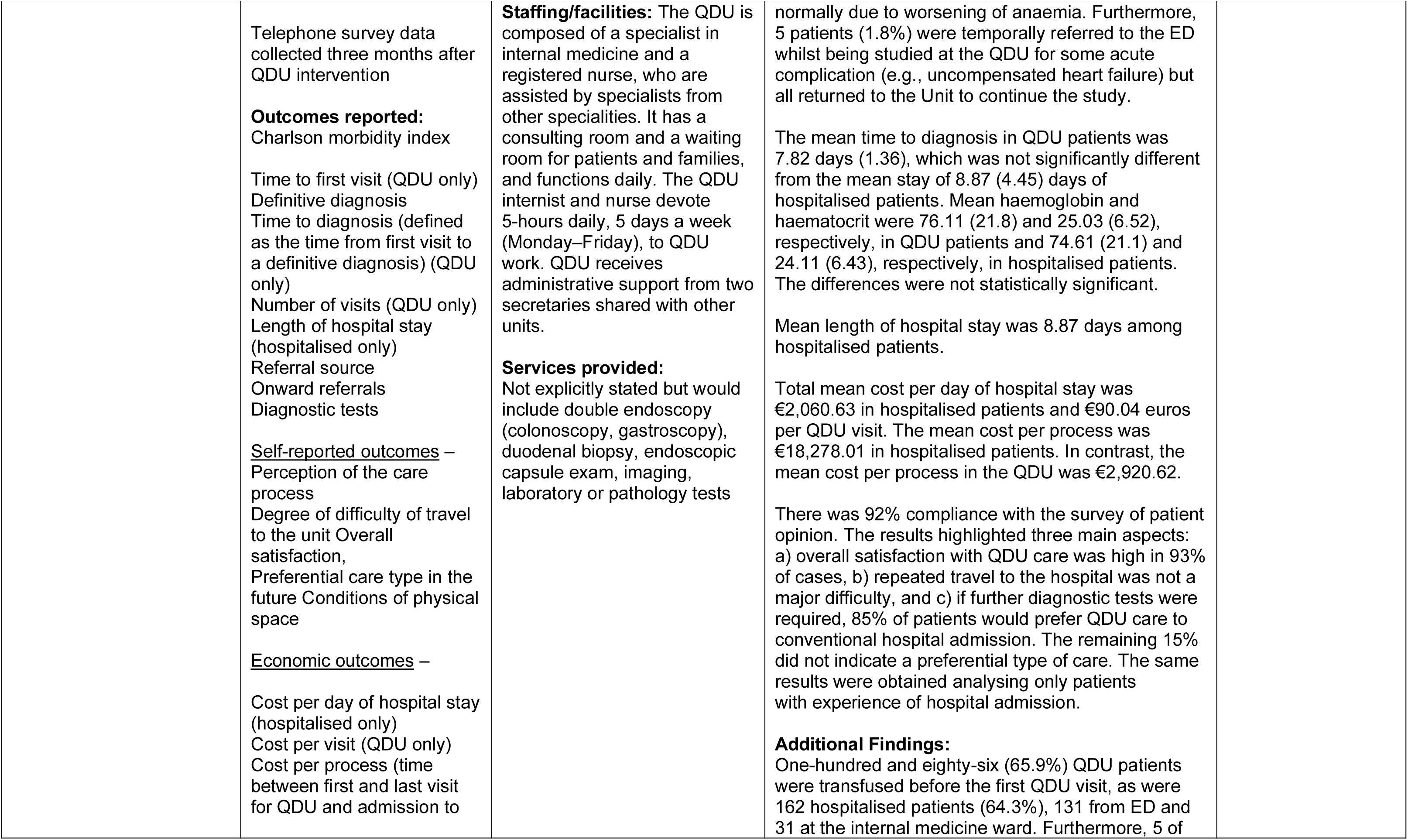
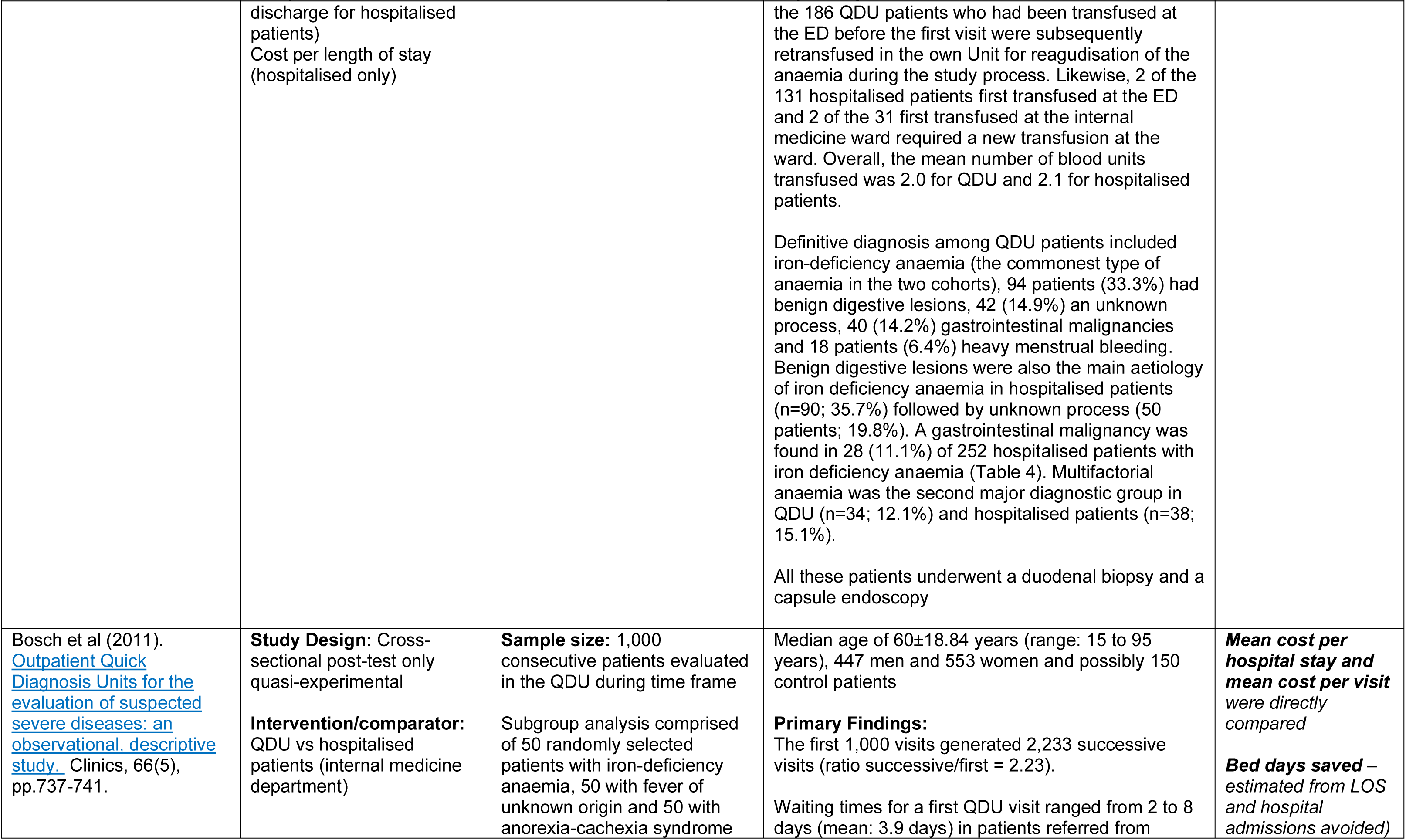
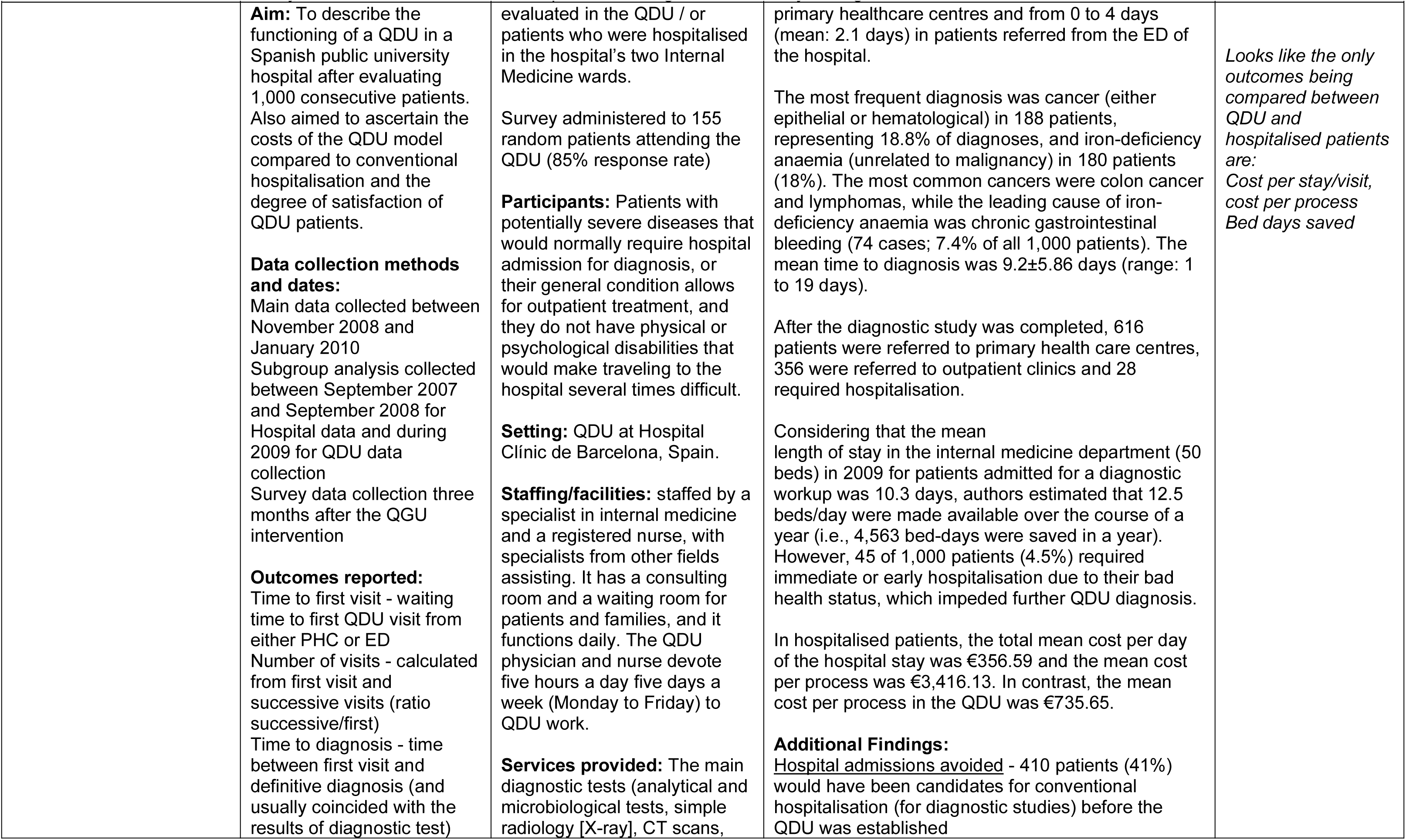
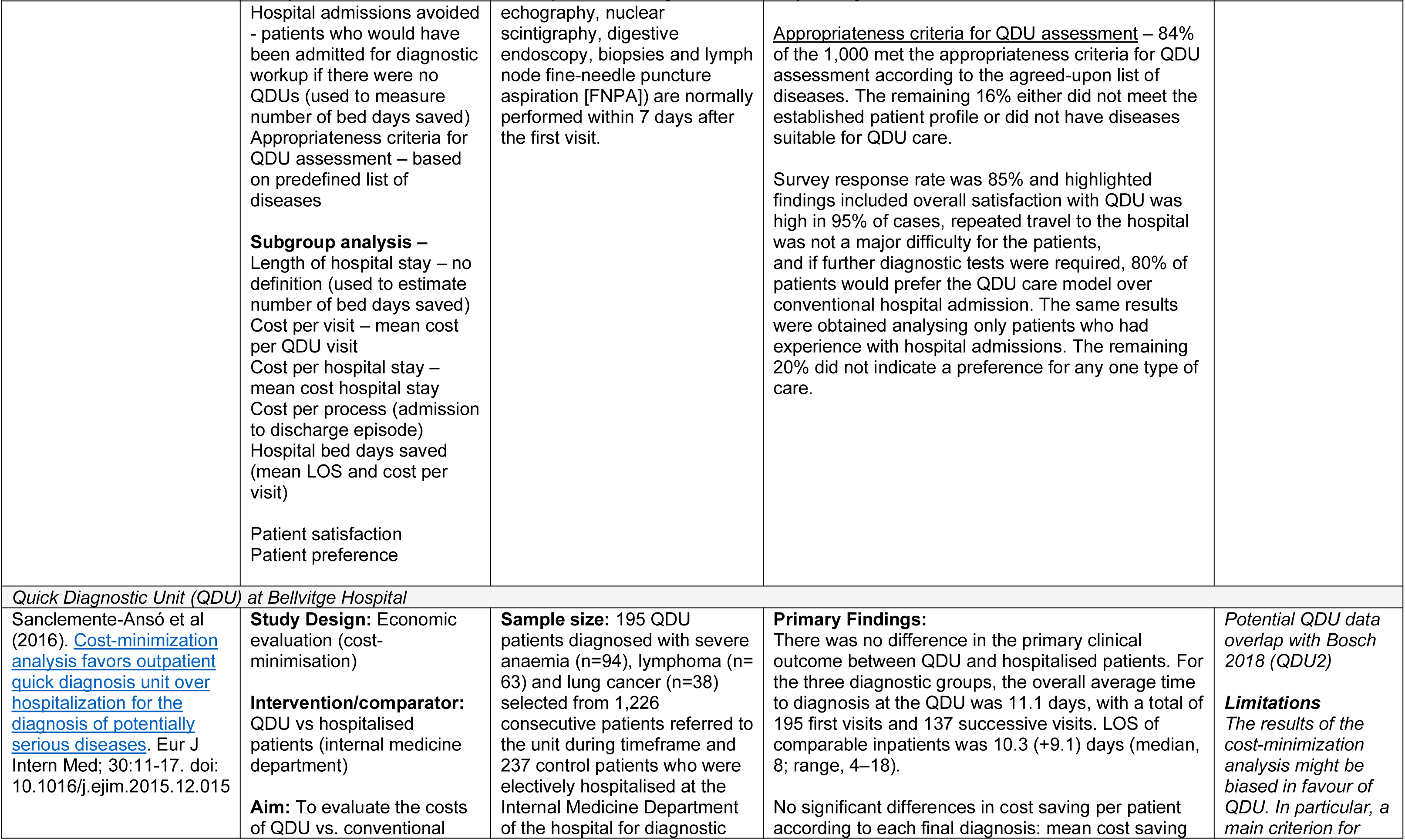
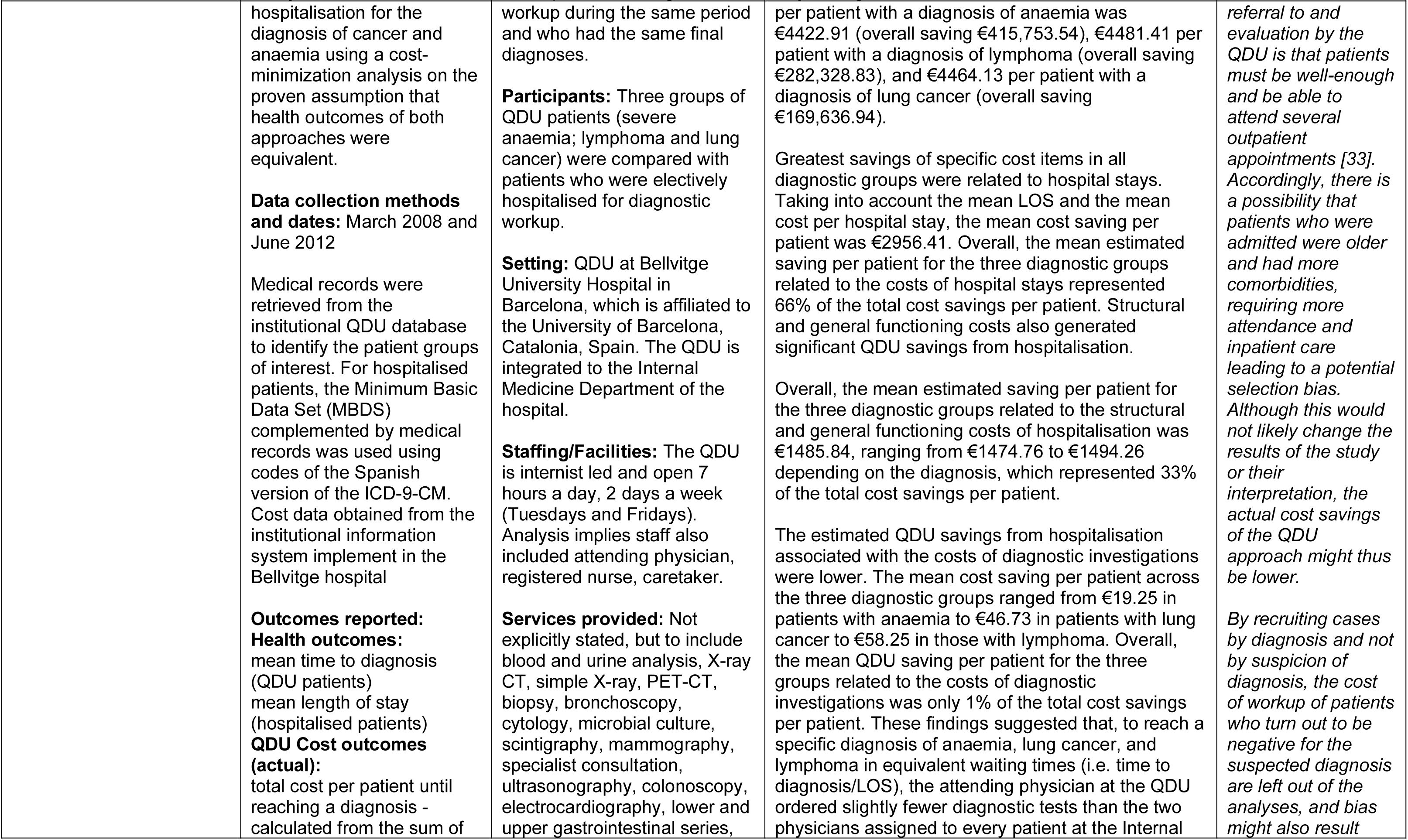
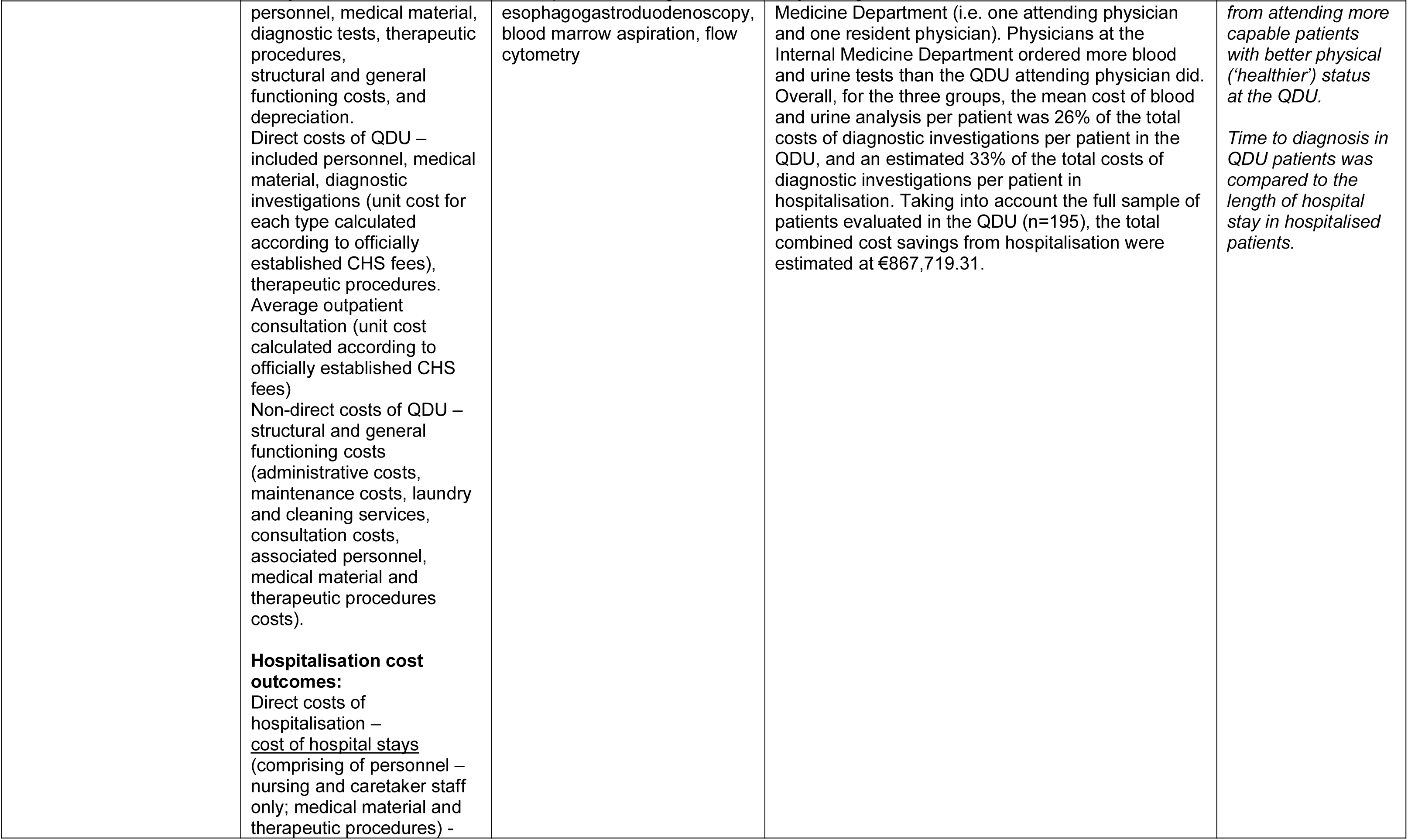
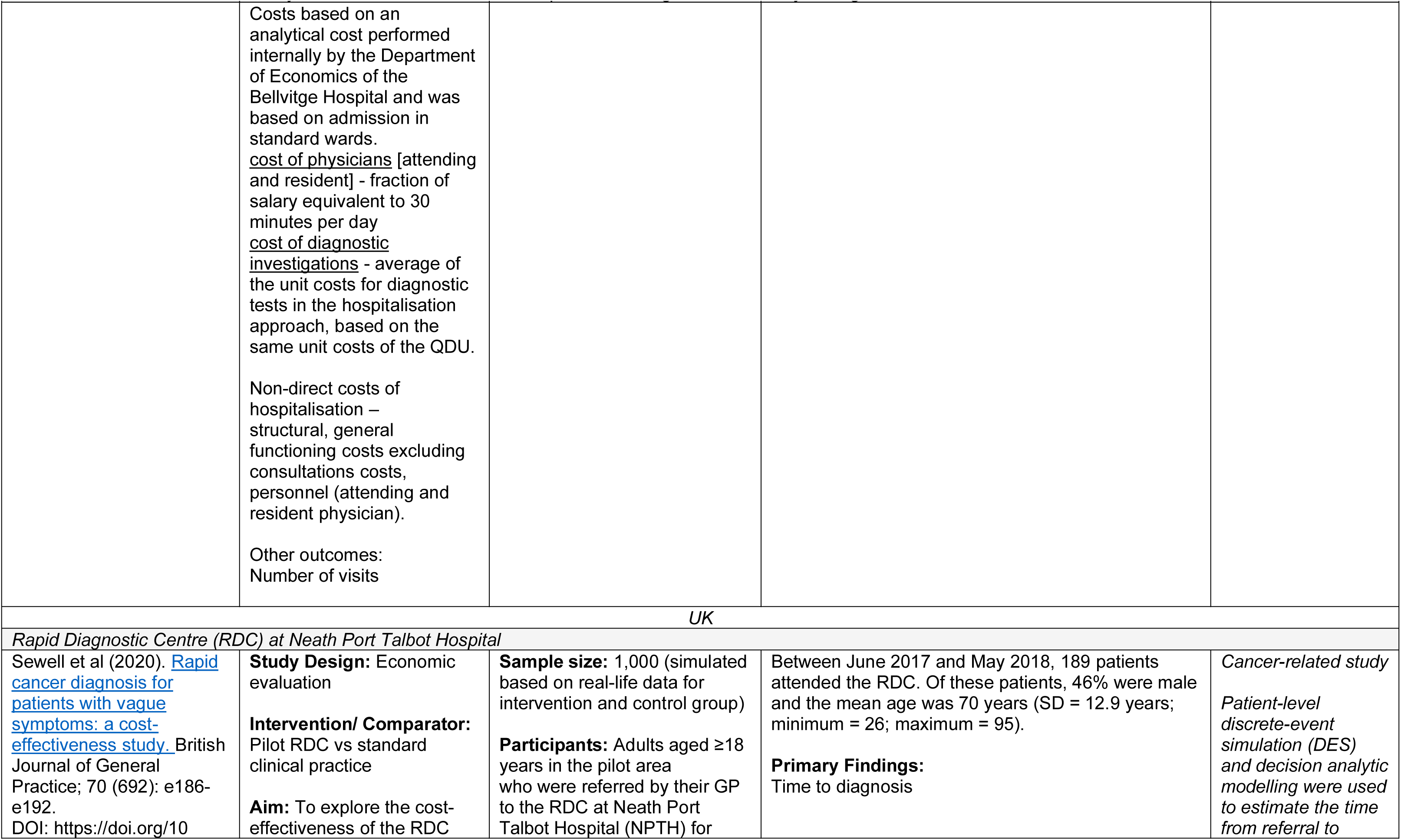
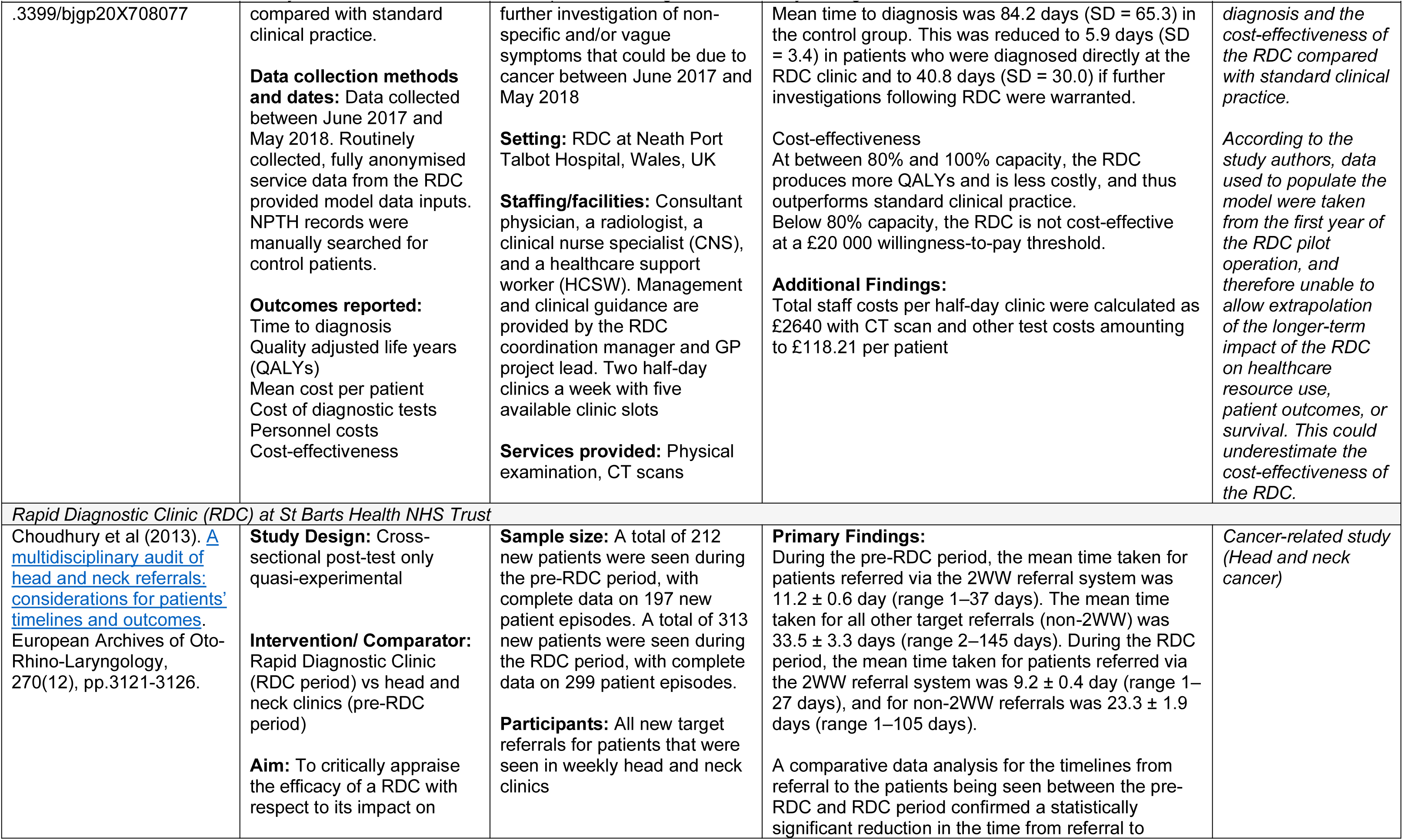
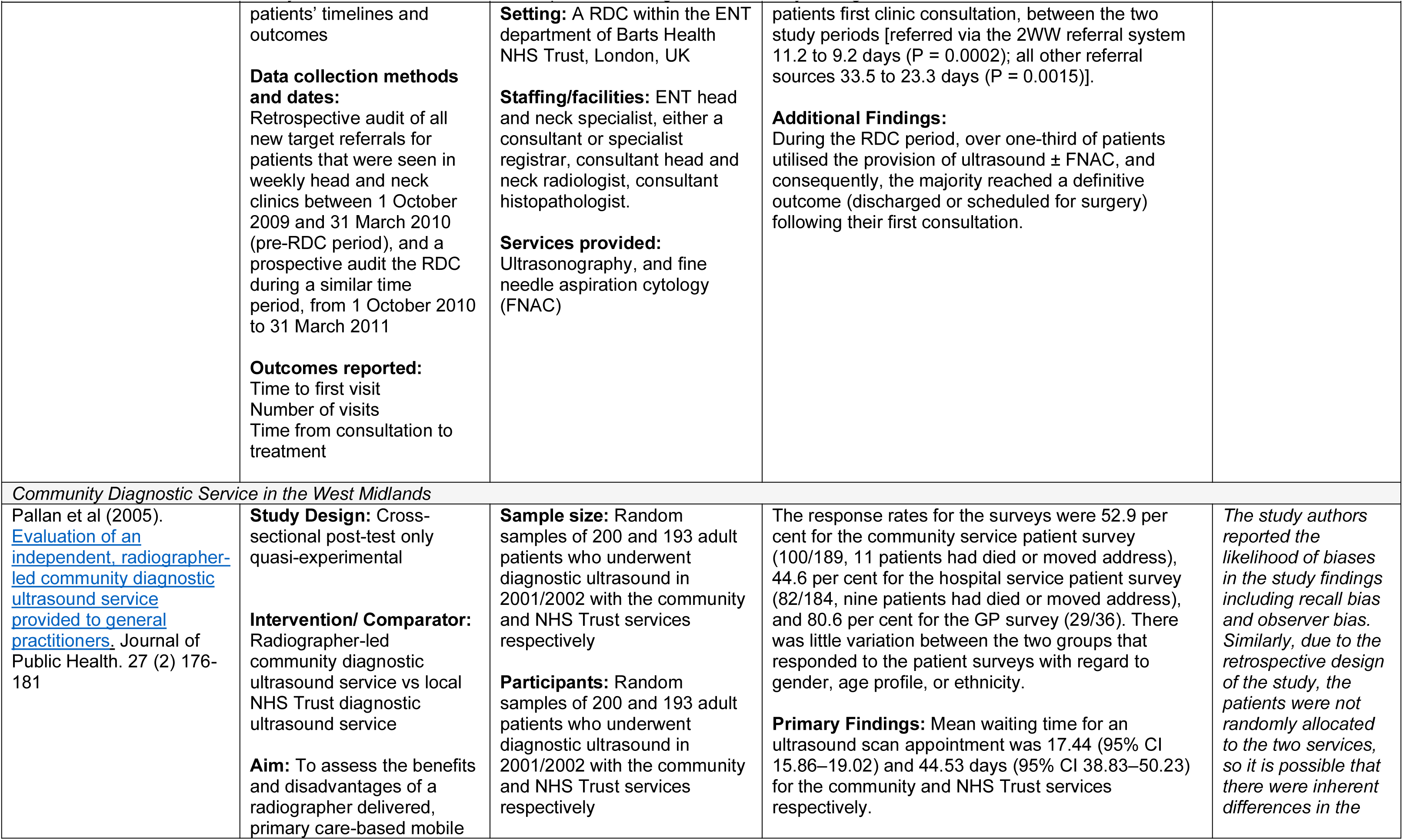
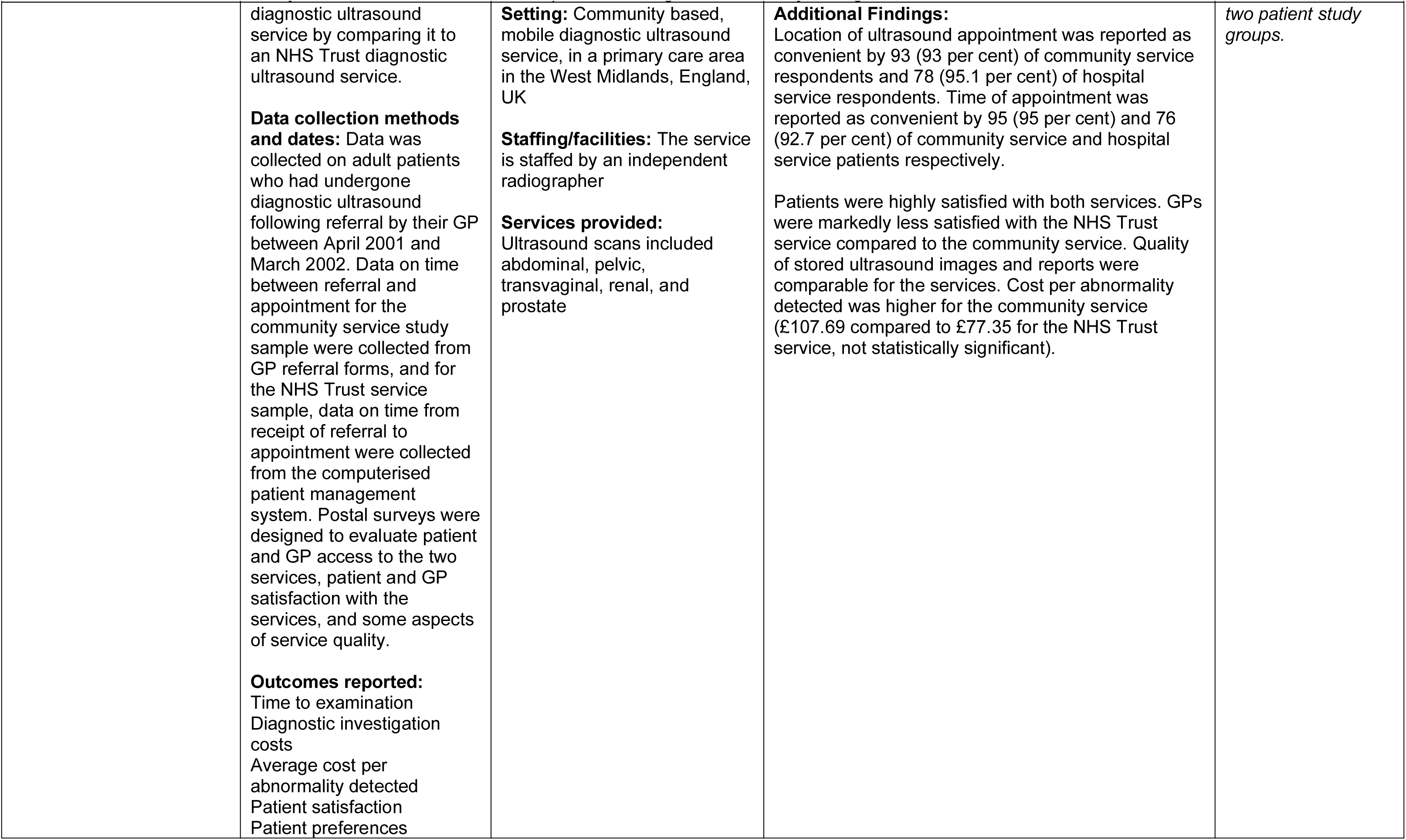
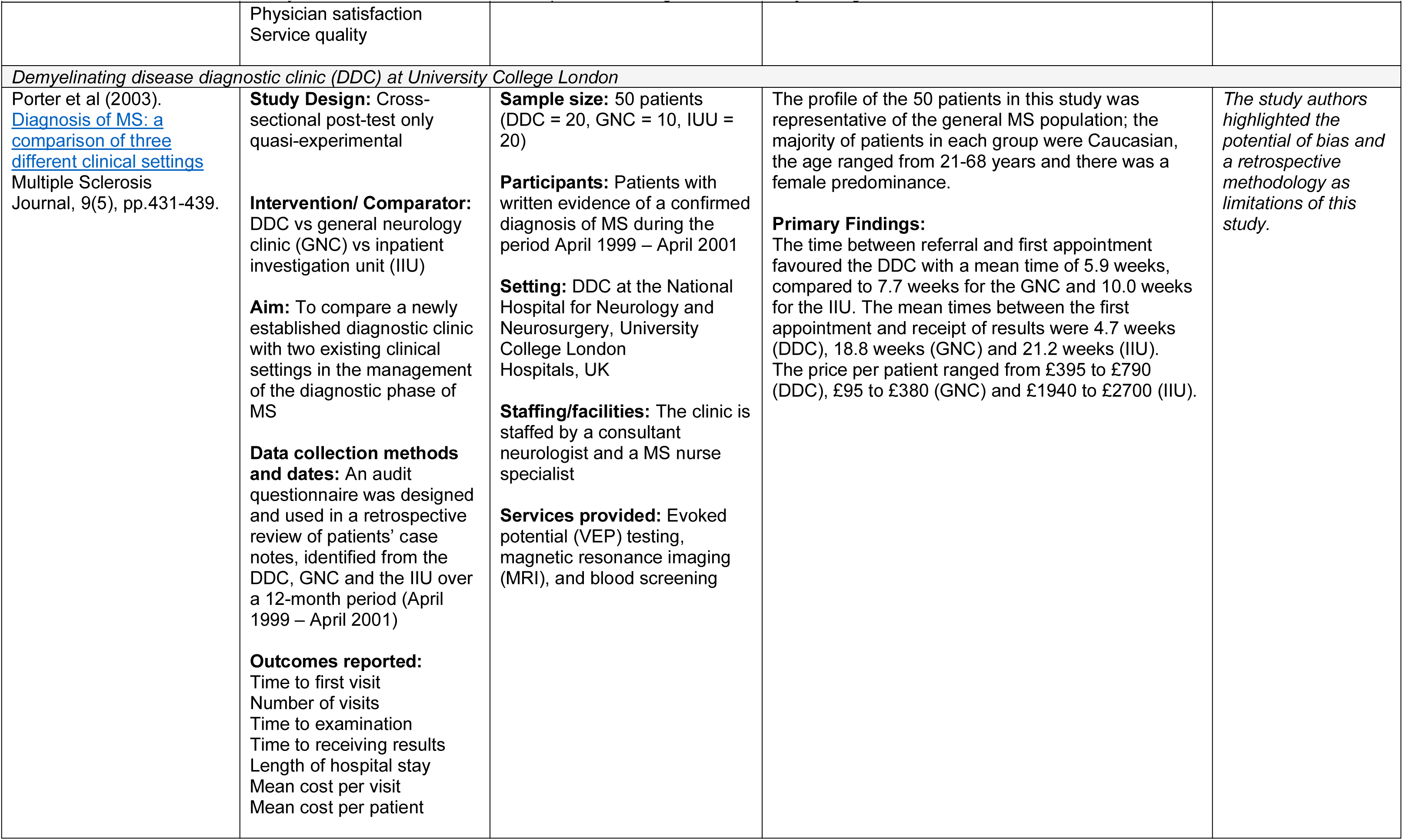
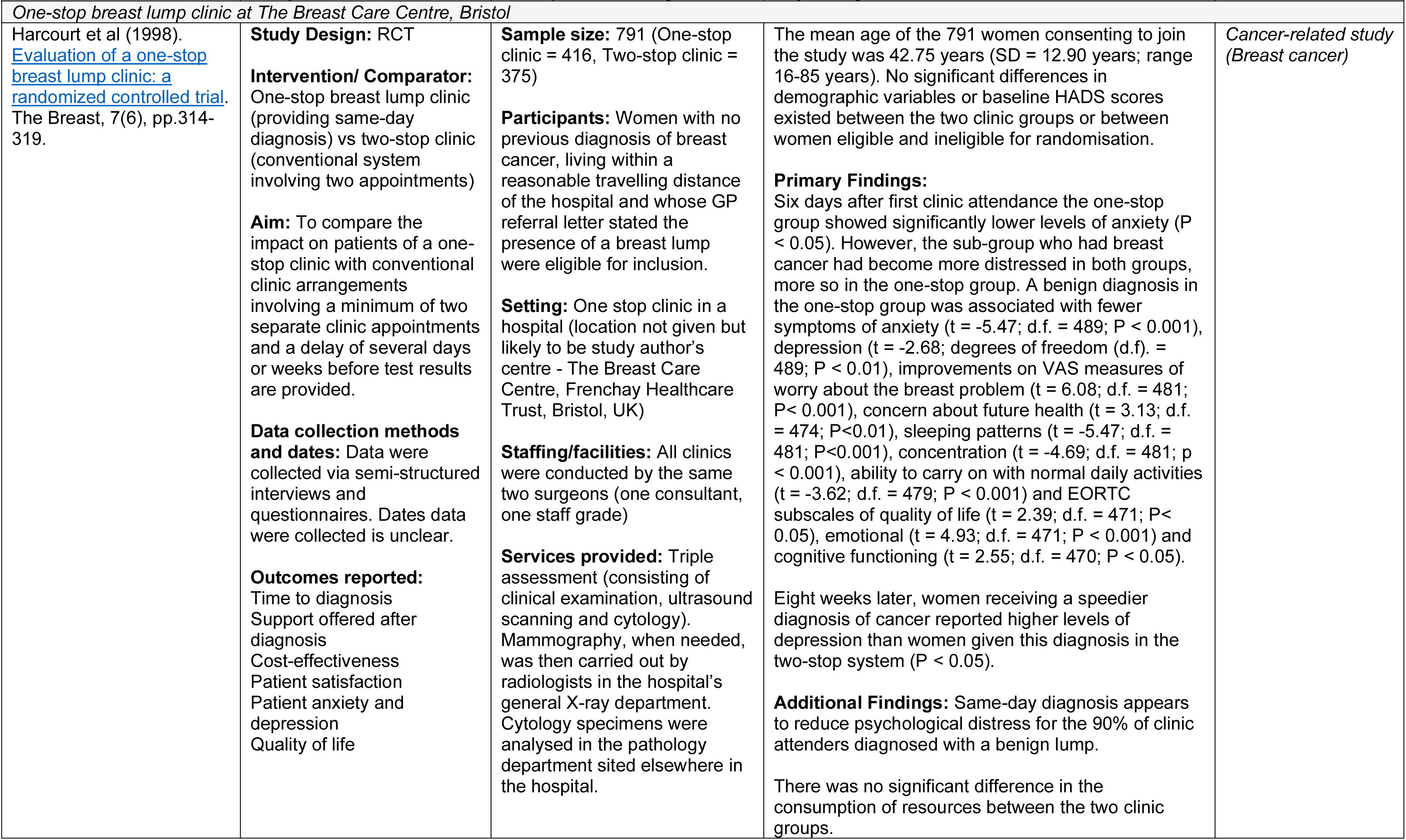
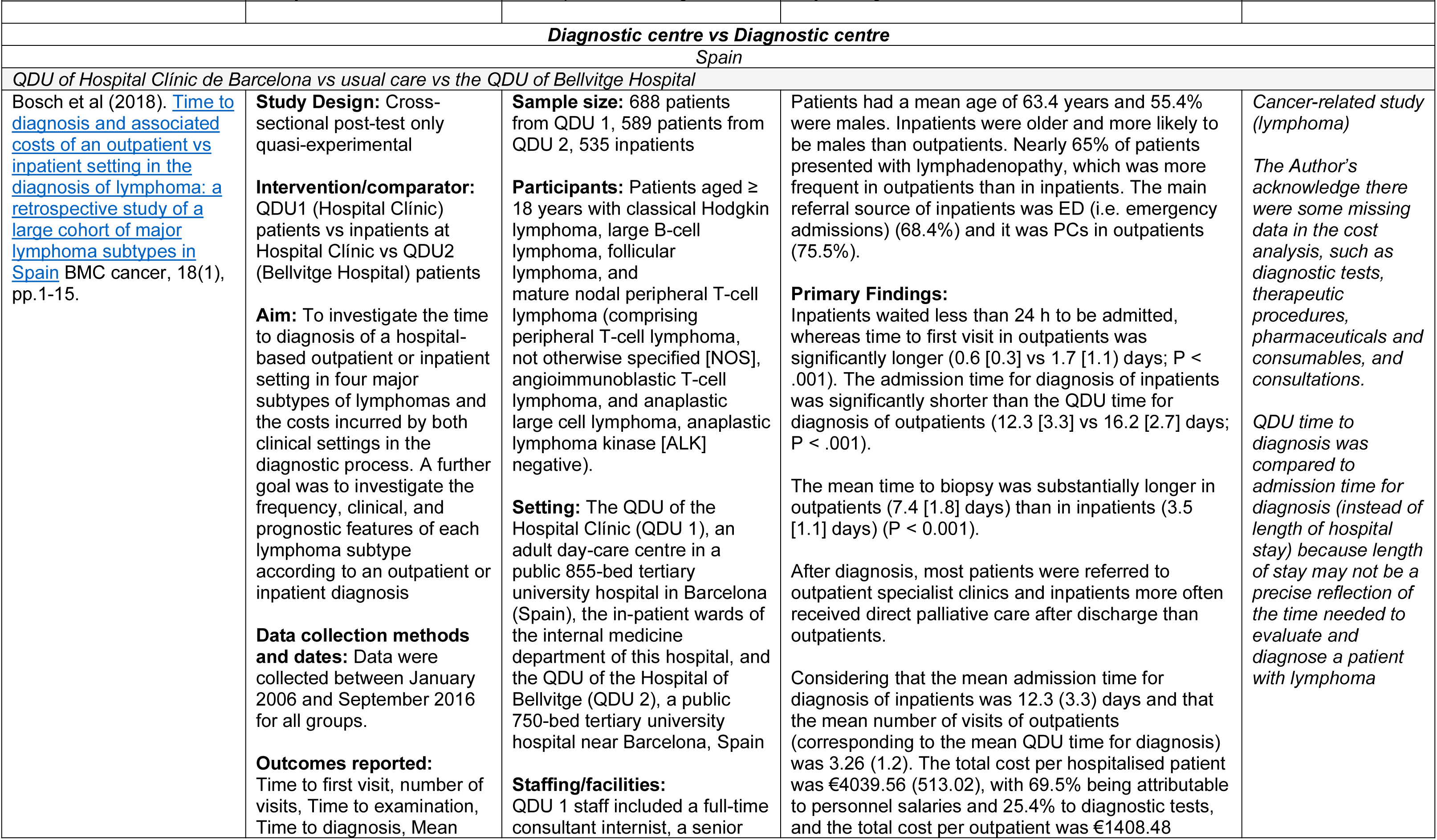
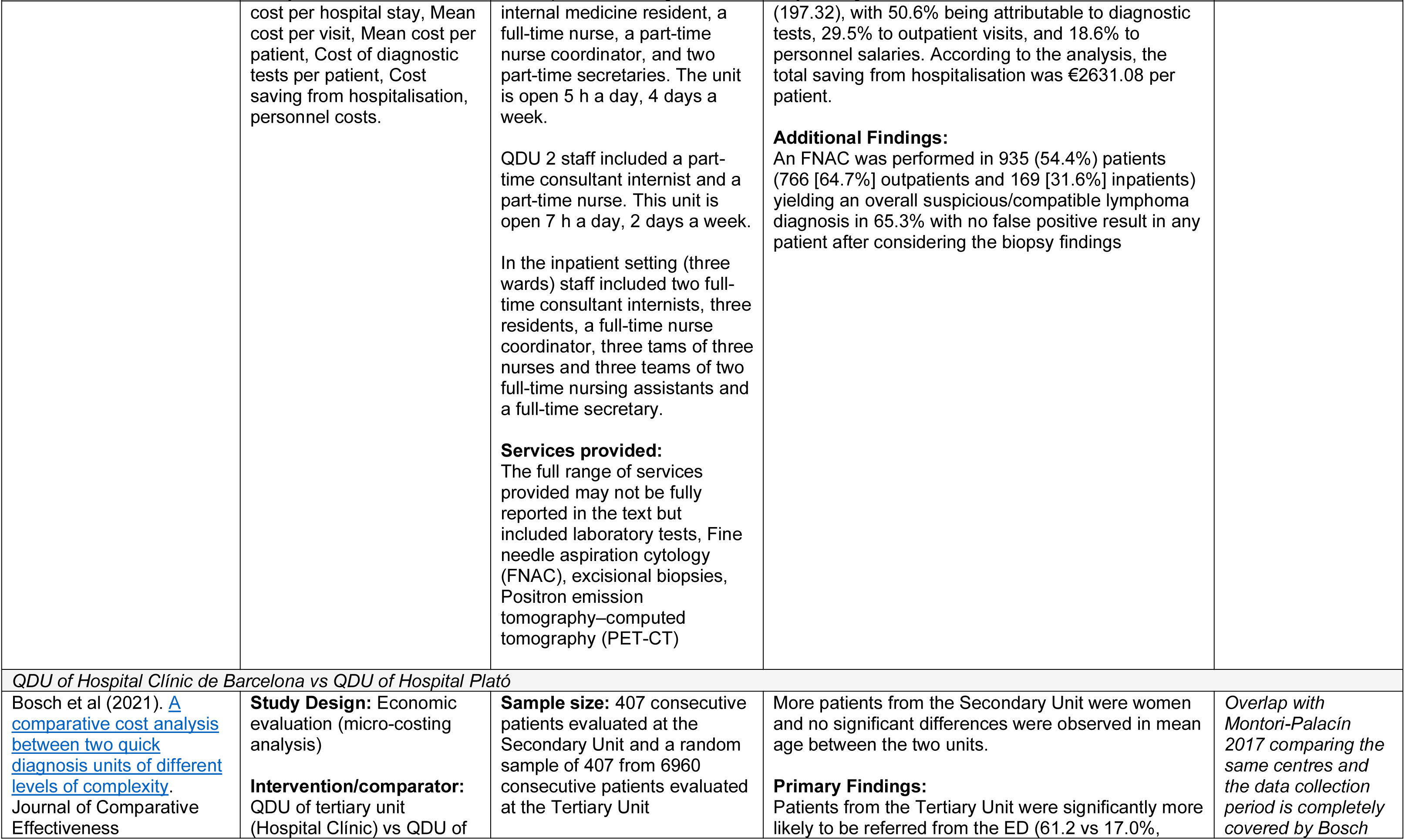
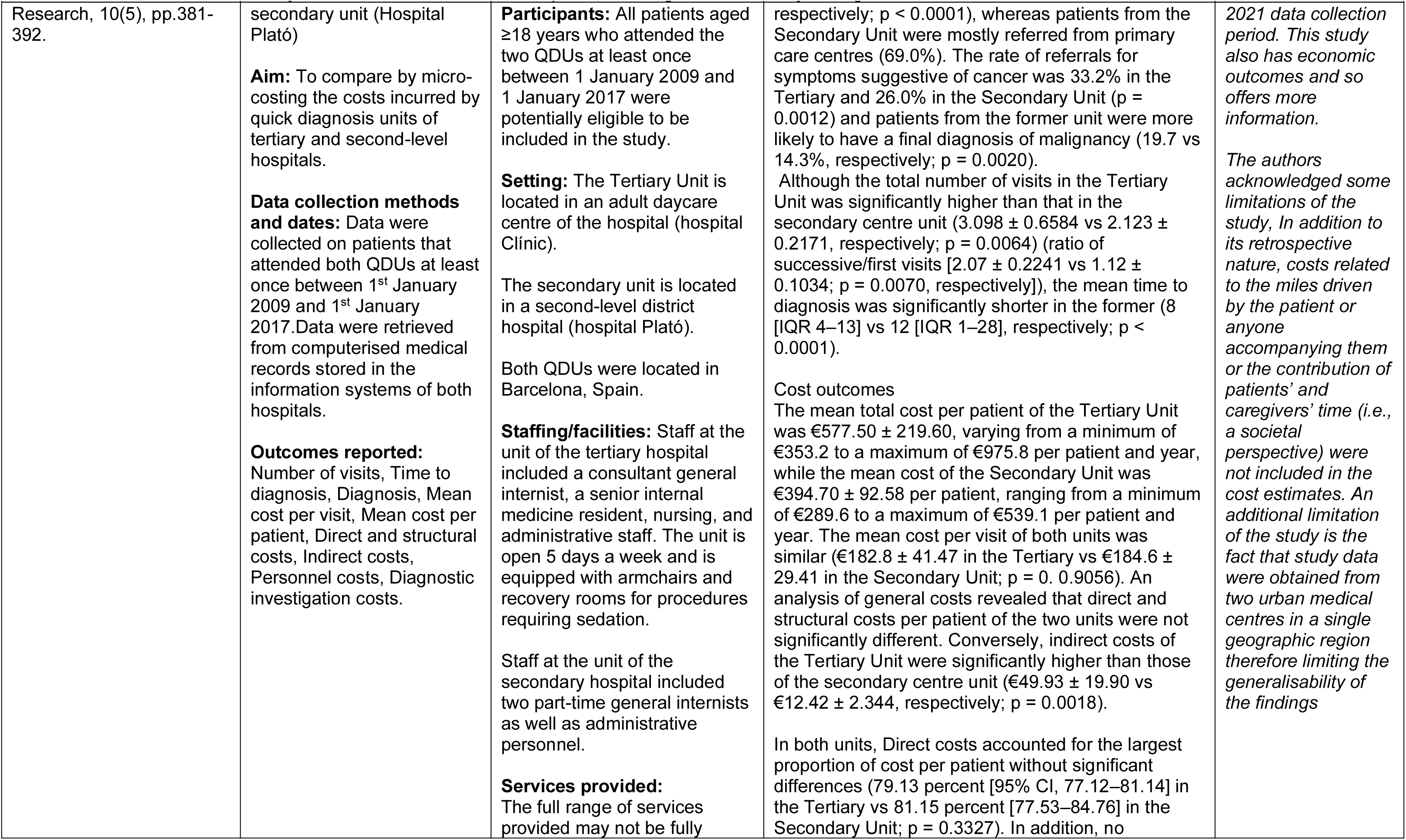
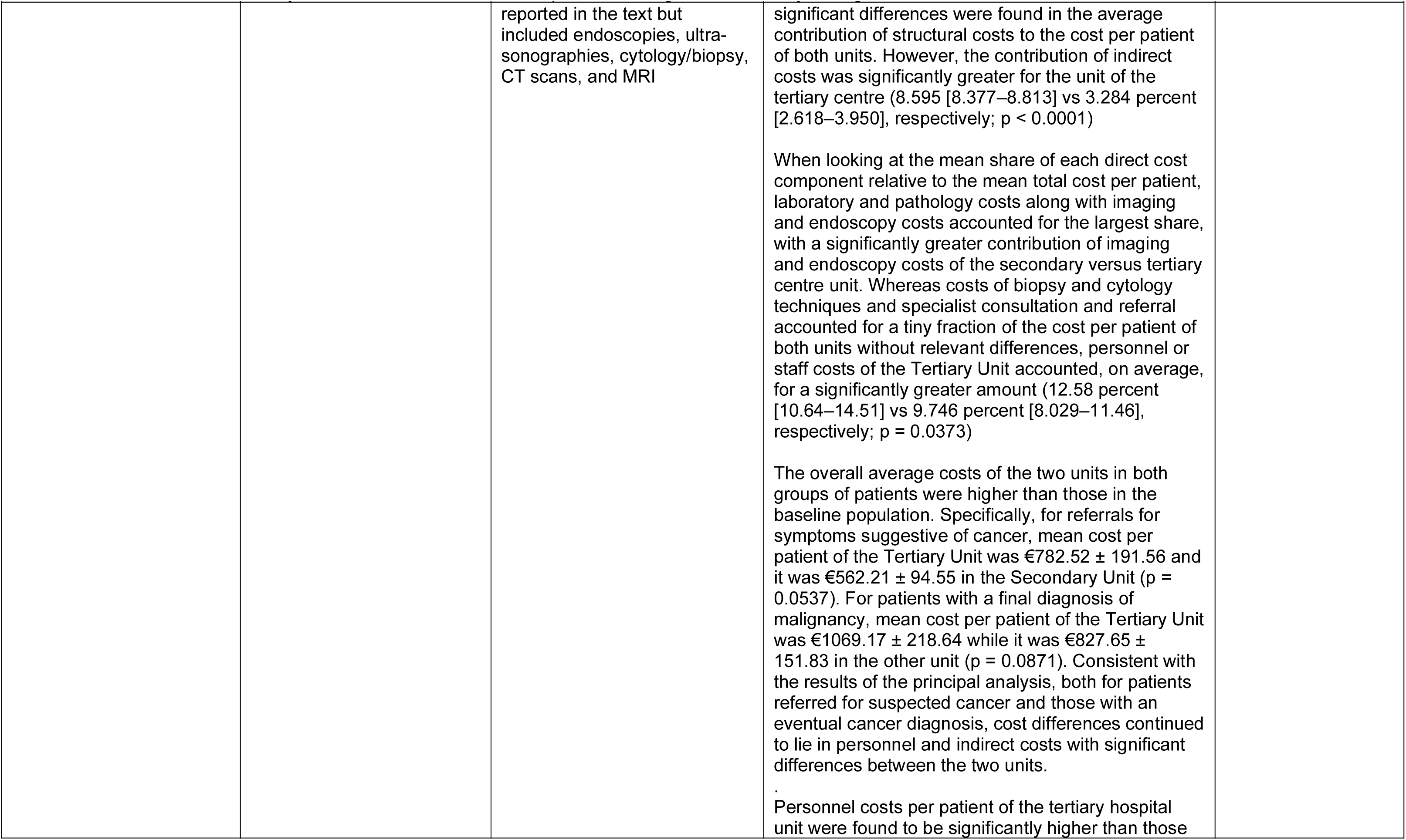
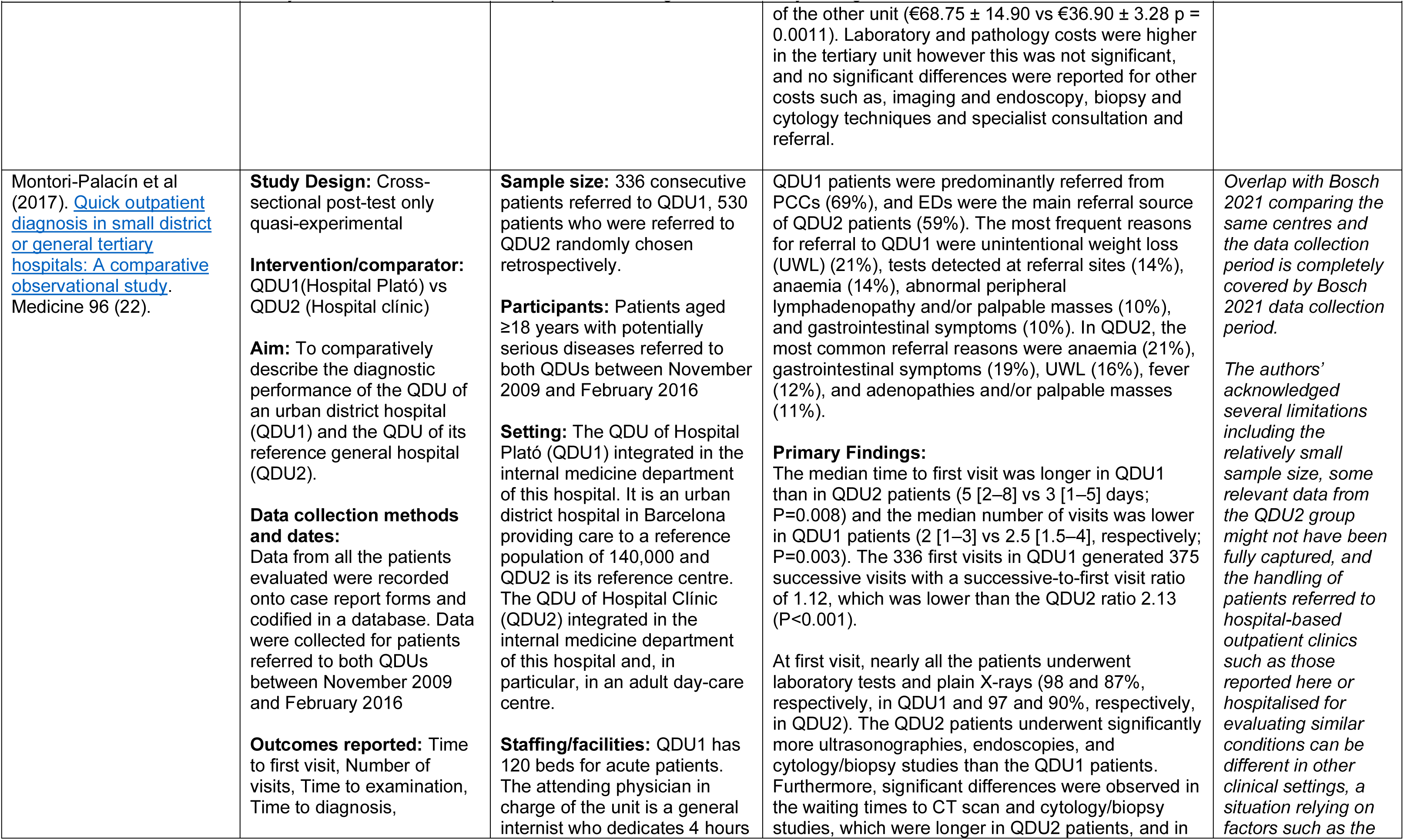
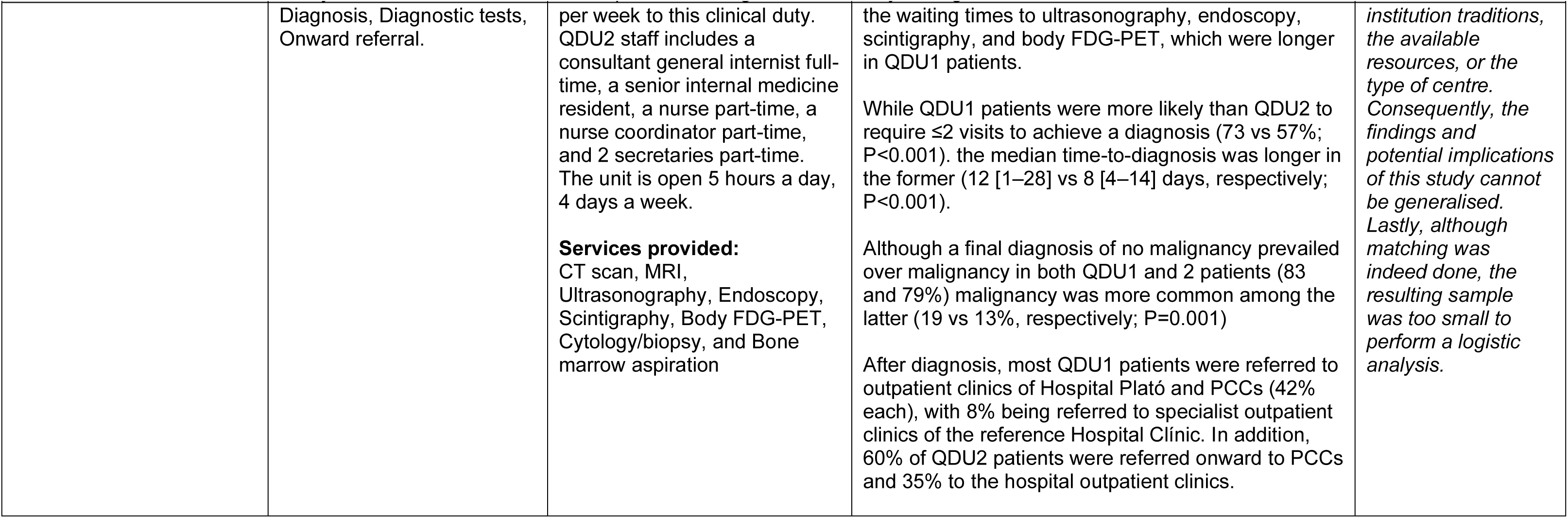
Summary of included studies

#### Twelve individual diagnostic centres were assessed across the 20 included studies

Details on the characteristics of each diagnostic centre can be found in Appendix 1. The majority of studies reported on diagnostic centres located within hospital settings (n=19). Only one study (Pallan et al 2005) reported findings from a diagnostic service located within the community setting. As outlined in our eligibility criteria (Table 2 in Section 5), all diagnostic centres accepted referrals from primary care as a minimum. The diagnostic centres in five studies accepted referrals solely from primary care while the remaining 15 studies accepted referrals from primary care and other referral sources such as emergency departments (ED) and hospital outpatient clinics.

The methodological quality of the included studies was assessed using the appropriate Joanna Briggs Institute (JBI) critical appraisal tool (for quasi-experimental studies, RCTs and economic evaluations) (see Tables 3, 4, 5). The JBI tool contains a set of signalling questions for particular domains of bias, including bias in selection of participants, bias in measurement of outcomes, and bias in selection of the reported results. Critical appraisal of the economic evaluation studies showed that all three studies measured outcomes and costs accurately. However, appraisal determined that none reported results that were generalisable to the setting of interest in our review. The single RCT measured all outcomes in a reliable way across groups but had issues with blinding of participants and outcome assessors. In the quasi-experimental studies, participants received similar treatment/care however, there appeared to be differences between participants included in compared groups. While all studies had some methodological issues, none were excluded from the review after quality appraisal.

### 2.2 Impact of diagnostic centres on waiting times

Nineteen included studies reported a range of outcomes relevant to waiting times. These included outcomes such as time to first visit, time to examination and time to diagnosis. In addition, some outcome measures relating to waiting times were specific to cancer studies, such as time from cancer suspicion to diagnosis, and time to surgical/specialist consultation. Some outcome measures relating to waiting times may have been measured at the same time intervals across the diagnostic pathway, but as they were often poorly defined across the studies, we have reported them separately.

#### Time to first visit

Time to first visit (the interval between referral and first visit to the diagnostic centre) was reported in 10 studies covering five diagnostic centres (three from Spain and two from the UK). The findings from the most recent studies are reported below for each diagnostic centre. Two studies covered the same diagnostic centre (Montori-Palacín et al 2017; Bosch et al 2020). However, one of these compared outcomes between patients referred to the diagnostic centre and patients hospitalised in inpatient wards (Bosch et al 2020), while the other compared the diagnostic performance of two diagnostic centres of different levels of complexity (secondary vs tertiary) (Montori-Palacín et al 2017). The findings of both studies are reported below. Study findings were generally mixed.

- A reduction in time to first visit was reported in two studies (Choudhury et al 2013, Porter et al 2003). Choudhury et al (2013) assessed the efficacy of a newly established rapid diagnostic clinic (RDC) for patients with suspected head and neck cancer by conducting an audit of new referrals made to a head and neck clinic during a six-month period before the new clinic was established (pre-RDC) and compared this with findings from the RDC period. The study reported a **statistically significant reduction in the time from referral to the patients being seen between pre-RDC period and RDC period** for both patients referred via the two-week wait (2WW) source (11.2 vs 9.2 days; p = 0.0002) and for patients referred via all other referral sources (33.5 vs 23.3 days; p = 0.0015).
- Porter et al (2003) compared a newly established demyelinating disease diagnostic clinic (DDC) with two existing clinical settings in the management of MS and reported that **the mean waiting time to first visit was shorter for the patients attending the DDC** (5.9 weeks) compared to the general neurology clinic (7.7 weeks) and the inpatient investigation unit (10.0 weeks) operating within the same hospital.
- Bosch et al (2020) compared outcomes between patients referred to a quick diagnostic unit (QDU) and patients hospitalised in inpatient wards and reported that the **time to first QDU visit (outpatients) was found to be significantly longer than the time to admission in hospitalised patients** (1.2 vs 0.7 days; p<0.001).
- Bosch et al (2018) compared outcomes between patients referred to two QDUs and patients hospitalised in inpatient wards and reported that the **time to first QDU visit (outpatients) was found to be significantly longer than the time to admission in hospitalised patients** (1.7 vs 0.6 days; p<0.001). It should be noted that two QDUs were combined and analysed as a single outpatient unit in this study.
- Montori-Palacín et al (2017) compared the diagnostic performance of two QDUs of different levels of complexity (secondary vs tertiary) and found that **the median time to first visit was longer in the QDU of a secondary hospital than that of the QDU of its reference general tertiary hospital** (5 vs 3 days; p=0.008). The study authors suggested that this difference could be due to direct appointments to the tertiary hospital QDU from the ED and a lack of administrative support of the secondary hospital QDU. This study, however, did not provide any non-QDU comparator data.

#### Time to diagnostic examination

Time to examination (the interval between diagnostic centre physician’s order and the examination being actually performed) was reported in four studies covering four different diagnostic centres (three from Spain and one from the UK). Two studies reported findings from the same diagnostic centre (Bosch et al 2012c; Bosch et al 2018), however one of these studies compared outcomes between patients referred to the diagnostic centre and patients hospitalised in inpatient wards (Bosch et al 2012c), while the other study combined two diagnostic centres into a single unit and compared outcomes with patients hospitalised in inpatient wards (Bosch et al 2018). The findings of both studies are therefore reported below. Similarly, Montori-Palacín et al (2017) and Bosch et al (2018) reported findings from the same diagnostic centre. However, unlike the latter, Montori-Palacín et al (2017) compared the diagnostic performance of the diagnostic centre with that of another diagnostic centre of a different level of complexity. Study findings were generally mixed.

- Pallan et al (2005) assessed the effectiveness of a primary care-based mobile diagnostic ultrasound service provided by an independent radiographer compared to an NHS Trust diagnostic ultrasound service and found that **the mean waiting time for an ultrasound scan appointment was shorter for the community service than for the hospital service** (17.44 days vs 44.53 days).
- Bosch et al (2018) compared waiting times of patients attending two outpatient QDUs versus conventional hospitalisation for the diagnosis of lymphoma and found that **the mean time to biopsy was significantly longer in the outpatient QDUs than in inpatient settings** (7.4 vs 3.5 days; p< 0.001). Two QDUs were combined and analysed as a single outpatient unit in this study.
- Bosch et al (2012c) compared waiting times of patients attending a QDU versus conventional hospitalisation for those with severe anaemia and found that the mean waiting time to gastroscopy was shorter in the QDU than in inpatient settings (3.23 vs 3.47 days) while the mean waiting time to colonoscopy was longer in the QDU than in inpatient settings (4.45 vs 4.24 days). However, these differences were not statistically significant.
- Montori-Palacín et al (2017) compared waiting times of two QDUs of different levels of complexity. Significant differences were observed in the waiting times to computed tomography (CT) scan (2 vs 3 days; p=0.03) and cytology/biopsy studies (3 vs 4 days; p=0.03), which were longer in patients attending the QDU of the tertiary hospital, and in the waiting times to ultrasonography (3 vs 2 days; p=0.04), endoscopy (12 vs 5 days; p<0.001), scintigraphy (7 vs 2 days; p<0.001), and body fluorodeoxyglucose-positron emission tomography (FDG-PET) (6 vs 3 days; p<0.001), compared to patients attending the QDU of the secondary hospital. However, this study did not provide non-QDU comparator data.

#### Time to diagnosis

Time to diagnosis (the time elapsed between the request of the decisive diagnostic procedure and the cyto/pathological diagnosis) was reported in 14 studies covering six diagnostic centres (three from Spain, two from Canada, and one from the UK). The findings from the most recent studies for each diagnostic centre are reported below. Two studies reported findings from the same diagnostic centre (Bosch et al 2020; Bosch et al 2021), however the former compared outcomes between patients referred to the diagnostic centre and patients hospitalised in inpatient wards, while the latter compared the diagnostic performance of two diagnostic centres of different levels of complexity. Both studies are reported below. Findings were generally mixed.

- Nixon et al (2020) examined the impact of a nurse practitioner–led lymphoma rapid diagnosis clinic (LRDC) by comparing findings from the initial 30-month experience of the LRDC with those prior to its implementation. **The time from initial assessment to lymphoma diagnosis was found to be significantly shorter for the patients attending the LRDC** compared to historical controls (16 vs 28 days; p<0.001).
- Sethukavalan et al (2013) compared wait time intervals for patients with prostate cancer diagnosed at a rapid diagnostic unit (RDU) versus the usual community process. The results showed **that time from urologist visit to diagnosis was significantly shorter in the RDU patients** compared to community patients (29 vs 100 days; p=0.0094).
- Sewell et al (2020) found **that patients diagnosed directly at a RDC had a shorter time to diagnosis than patients receiving usual care** (5.9 days vs 84.2 days). However, this study was conducted using patient-level discrete-event simulation (DES) and decision analytic modelling.
- Bosch et al (2020) found **no significant differences** between admission time to diagnosis (hospitalised patients) and the QDU time to diagnosis (4.1 vs 4.3 days; p=0.163).
- Bosch et al (2018) found that the **time to diagnosis of QDU outpatients was significantly longer than the admission time to diagnosis among inpatients** (hospitalised) (16.2 vs 12.3 days; p<0.001). Two QDUs were combined and analysed as a single outpatient unit in this study.
- Bosch et al (2021) compared the diagnostic performance of two QDUs of different levels of complexity (secondary vs tertiary) and found that the **time to diagnosis was significantly longer in the QDU of a secondary hospital than in QDU of its reference general tertiary hospital** (12 vs 8 days; p<0.0001). This study, however, does not provide any non-QDU comparator data.

#### Wait time from abnormal imaging to biopsy and from biopsy to pathology verification

One study conducted in Canada reported on the wait time from abnormal imaging to biopsy and from biopsy to pathology verification.

- Arnaout et al (2013) assessed the impact of a Rapid Diagnosis and Support (RADS) programme on diagnostic and treatment wait times for patients with a high probability of breast cancer compared to patients who had BI-RADS 5 diagnostic imaging in the year before the programme (PRE-RADS). **Significant reductions were found in the mean wait time from abnormal imaging to biopsy** (7.1 days vs 3 days; p<0.01) **and from biopsy to pathology verification** (3.9 days vs 3.4 days; p<0.01) **for patients attending the RADS programme** compared to PRE-RADS.

#### Time to surgical consultation

Time to surgical consultation (the time from presentation to seeing the surgeon/ the interval between pathology verification to surgical consultation) was reported by two Canadian studies investigating two different rapid diagnostic services. It should be noted that these studies use different start points to measure this outcome, as described above. Both studies identified a statistically significant reduction in wait times compared to their comparators.

- McKevitt et al (2017) reported that **patients seen at a Rapid Access Breast Clinic (RABC) had a significantly decreased time from presentation to surgical consultation** (33 vs 86 days; p<0.0001) for both malignant (36 vs 59 days; p=0.0007) and benign diagnoses (31 vs 95 days; p<0.0001) compared to patients diagnosed through the traditional system (TS).
- Arnaout et al (2013) reported that **the time from pathology verification to surgical consultation had been reduced significantly** from 16.1 to 5.9 days (p<0.01) in patients who took part in **a RADS programme** compared to patients who had BI-RADS 5 diagnostic imaging in the year before the programme (PRE-RADS).

#### Time from cancer suspicion to treatment

One study conducted in Canada reported the time from cancer suspicion to treatment (the time from suspicion of cancer to radiotherapy).

- Sethukavalan et al (2013) compared wait time intervals for patients with prostate cancer diagnosed at a multidisciplinary RDU versus the usual community referral process. The results showed a **statistically significant difference in the time interval from cancer suspicion to treatment** (158 days vs 218; p = 0.046) **in favour of the patients attending the RDU.**

#### Time from consultation to treatment

Time from consultation to therapy (the time interval from consultation to treatment/surgery) was reported in five studies covering five different diagnostic centres (four from Canada and one from UK). All studies identified a reduction in time from consultation to treatment compared to their comparators, but only two identified a statistically significant difference.

- Nixon et al (2020) found that **the time from initial assessment to treatment of aggressive lymphomas and Hodgkin’s lymphoma was significantly shorter for patients attending a LRDC** compared to historical controls (29 days vs 48 days; p<0.001).
- Arnaout et al (2013) reported that **time from surgical consultation to treatment was reduced significantly** from 31.5 to 24.1 days (p=0.04) **for patients who took part in a RADS programme** compared to patients who had BI-RADS 5 diagnostic imaging in the year before the programme (PRE-RADS).
- McKevitt et al (2017) reported that patients seen at a RABC had a **decreased time from surgical consultation to surgery** compared to patients diagnosed through the TS (31 days vs 33 days). However, this difference was not statistically significant (p=0.78).
- Choudhury et al (2013) reported that patients seen during the RDC period had a **decreased time from initial consultation to date of surgery** compared to patients seen during the pre-RDC period (32.5 vs 38.9 days) however this difference was not statistically significant (p = 0.307).
- Sethukavalan et al (2013) found **the time from consultation to treatment to be shorter for patients attending a RDU compared** to the usual community referral process (60 vs 62 days), however this difference was not statistically significant (p = 0.52).

#### Time from diagnosis to specialist consultation

One study conducted in Canada reported the outcome time from diagnosis to specialist consultation (the time from diagnosis to radiation oncology consult).

- Sethukavalan et al (2013) found the **time from diagnosis to specialist consultation was significantly shorter for patients attending a RDU** compared to the usual community referral process (27 days vs 49 days; p=0.0019).

#### Time from diagnosis to treatment

One study conducted in Canada reported the outcome time from diagnosis to treatment.

- Sethukavalan et al (2013) found the **time from diagnosis to treatment was significantly shorter for patients attending a RDU** compared to the usual community referral process (mean 91 days vs 120; p=0.016).

#### 2.2.1 Bottom line results for the impact of diagnostic centres on waiting times

Evidence relating to the impact of diagnostic centres on waiting times appears to be mixed. There is evidence to suggest that the utilisation of diagnostic centres can reduce various waiting times, including time to surgical consultation and time from consultation to treatment. However, the evidence was mixed for other wait time outcomes including the time to first visit, time to diagnostic examination and time to diagnosis.

Reductions in waiting times were also reported for the time to biopsy, from cancer suspicion to treatment, from diagnosis to specialist consultation and from diagnosis to treatment, for patients attending diagnostic centres. However, these outcomes were reported by individual studies and as such firm conclusions cannot be made. Furthermore, the methodological limitations across included studies and variations in healthcare systems of the countries of origin of included studies, could limit the applicability of all findings.

### 2.3 Impact of diagnostic centres on capacity and pressure on secondary care

Thirteen studies reported a range of outcomes relevant to the impact of diagnostic centres on capacity and pressure on secondary care. These included outcomes such as the number of visits required to obtain a definite diagnosis, number of biopsies required to arrive at a diagnosis, onward referrals and referral patterns over time.

#### Number of visits required to receive a diagnosis

The number of visits to the diagnostic centre required to obtain a definite diagnosis was reported by four studies covering three diagnostic centres (two from Spain and one from the UK). The findings from the most recent studies are reported below for each diagnostic centre. Findings were unclear.

- Bosch et al (2021) investigated the costs incurred by two QDUs of tertiary and secondary hospitals and found that **significantly fewer visits were required to achieve a diagnosis at the secondary unit compared to the tertiary unit** (2.123 vs 3.098 visits; p = 0.0064). However, this study does not provide any non-QDU comparator data.
- Porter et al (2003) compared a newly established DDC to two existing clinical settings in the management of MS. They found **that patients attending the DDC required two visits** (one initial and one follow-up appointment) before receiving a definite diagnosis compared to one to four visits, and two to five visits respectively, for the other clinical settings.

#### Number of biopsies to arrive at a diagnosis

One study conducted in Canada reported the number of biopsies required to arrive at a definitive diagnosis.

- Nixon et al (2020) examining the effectiveness of a nurse-led LRDC, found **that significantly fewer patients required two or more biopsies to arrive at a diagnosis of lymphoma after institution of a LRDC compared with patients diagnosed prior to the implementation of the clinic** (40% vs 12%; p<0.001).

#### Referral patterns over time and onward referrals

One study conducted in Spain reported referral patterns over time.

- Bosch, Jordán and López-Soto (2013) conducted a study to determine whether QDUs could be used to safely and efficiently avoid ED visits and hospitalisations. This study compared the referral trends over time of patients with suspected serious disease, from primary care and EDs to the QDU. **They found statistically significantly more direct referrals to the QDU (from 36% to 64%) and less to the ED (from 65% to 35%) respectively during the 25 month study period** (p<0.0001). In addition, at least 84% of hospitalised patients were found to be stable and their hospitalisations might have been avoided.

Onward referrals after attending a diagnostic centre were reported by nine studies covering four diagnostic centres (three from Spain and one from UK). The findings from the most recent studies are reported below for each diagnostic centre. Two studies reported findings from the same diagnostic centre (Montori-Palacín et al 2017; Bosch et al 2018). However, Montori-Palacín et al (2017) compared the diagnostic performance of the diagnostic centre with that of another diagnostic centre of a different level of complexity, while Bosch et al (2018) combined two diagnostic centres into a single unit and compared outcomes with patients hospitalised in inpatient wards. Both are reported below.

- Bosch et al (2018) reported onward referrals for patients attending one of two QDUs and compared this to hospitalised patients. The study found the majority of QDU patients were referred to outpatient specialist clinics (95.1% and 96% respectively, compared to 92% of controls). Patients were also referred to primary care (3.1% and 3% respectively compared to 2.1% of controls) and to palliative care (1.9% and 1% compared to 6% of controls). **The patients attending the QDUs were significantly more likely to be referred to outpatient specialist clinics (p=0.046) and were significantly less likely to be referred to palliative care** (p<0.001). However, authors reported this significant difference is likely related to the fact that the inpatient cohort were generally older and likely to have more aggressive lymphoma subtypes than those attending the diagnostic centres.
- Montori-Palacín et al (2017) assessed the diagnostic performance of a QDU of a secondary hospital by assessing patients with potentially serious disorders and compared it with a tertiary hospital QDU. The study found that after diagnosis**, most secondary hospital QDU patients were referred to the outpatient clinics of the secondary hospital and primary care centres (42% each), and 8% referred to specialist outpatient clinics at the tertiary hospital. In contrast, 60% of the tertiary hospital QDU patients were referred onward to primary care centres and 35% to the hospital outpatient clinics.** However, this study does not provide any non-QDU comparator data.
- Choudhury et al (2013) assessed the efficacy of a newly established RDC by conducting an audit of new referrals to a head and neck clinic during a six-month pre-RDC period and compared this with findings from the RDC period. The study reported an increase in the number of patients in whom a definitive outcome was reached (discharged or being listed for surgery) from the RDC. **In the pre-RDC period, one-third (33%) of patients reached a clear management plan including one of these two definitive outcomes, compared to almost one half (48 %) of all patients who attended the RDC, who were either discharged or scheduled for surgery.** Similarly, the number of patients that needed to be referred for an investigation fell by more than half, from 37 % in the pre-RDC period, to 15 % from the RDC period.

#### 2.3.1 Bottom line results for the impact of diagnostic centres on capacity and pressure on secondary care

Evidence relating to the impact of diagnostic centres on capacity and pressure in secondary care appears to be unclear. There is evidence to suggest that diagnostic centres could reduce the number of visits needed to receive a definite diagnosis. The evidence also suggest diagnostic centres could reduce the number of biopsies needed to arrive at a diagnosis and reduce the number of stable patients being referred for hospitalisation overtime however, these outcomes were only reported by individual studies and as such firm conclusions cannot be made.

Onward referrals were made to a range of settings including primary care centres, palliative care, outpatient clinics and referrals for surgery. The evidence suggests onward referrals differed in diagnostic centres when compared with hospitalisation (however this is likely to be due to the differences of the patients who are able to attend a diagnostic centre and those who require hospitalisation). The evidence suggests that diagnostic centres can increase the number of patients reaching a clear management plan (discharged or scheduled for surgery) and reduce the need to be referred for further investigations. However, these findings were only reported by individual studies and as such firm conclusions cannot be made. Onward referrals appeared to differ between diagnostic centre patients and inpatients, possibly due to the fact that inpatients were generally older and more unwell than those attending diagnostic centres. The methodological limitations across included studies and variations in healthcare systems of the countries of origin of included studies could further limit the applicability of these findings.

### 2.4 Economic impact of diagnostic centres and other economic outcomes

Fourteen studies covering seven diagnostic centres (three from UK, three from Spain, and one from Canada) reported economic outcomes. Of these, three studies were economic evaluations: one cost-minimisation analysis (Sanclemente-Ansó et al 2016), one cost-effectiveness study (Sewell et al 2020), and one comparative cost analysis (Bosch et al 2021), while the other studies (11 quasi-experimental studies) reported more generic economic outcomes.

#### 2.4.1 Economic evaluations

Bosch et al (2021) conducted a comparative cost analysis using micro-costing, to compare the costs incurred by two QDUs of different levels of complexity (tertiary vs secondary). The results showed that the mean total cost per patient of the tertiary unit was €577.50 ± 219.60, while the mean cost of the secondary unit was €394.70 ± 92.58 per patient (p value for difference between the two units = 0.0559). The mean cost per visit of both units was similar (€182.8 ± 41.47 in the tertiary vs €184.6 ± 29.41 in the secondary unit; p = 0.9056). An analysis of general costs revealed that direct and structural costs per patient of the two units were not significantly different. Conversely, indirect costs of the tertiary unit were significantly higher than those of the secondary unit (€49.93 ± 19.90 vs €12.42 ± 2.344, respectively; p = 0.0018). **The main driver of the cost differences between the two QDUs was found to be the total number of visits and successive/first visits ratio.** However, this study does not provide any non-QDU comparator data.

Sewell et al (2020) used patient-level DES and decision analytic modelling to estimate the cost-effectiveness of a pilot RDC in its first year of operation compared with standard clinical practice. The results showed that during the start-up phase of the service, the RDC was seeing a mean number of 2.78 patients per clinic and was more costly and more effective compared to standard clinical practice. However, **when run at near or full capacity (80% or higher, seeing a mean number of 4.65 patients/clinic), the RDC was found to outperform usual care, i.e. being less costly and more effective.**

Sanclemente-Ansó et al (2016) conducted a cost-minimisation analysis to assess the costs of the QDU approach compared with the costs of conventional hospitalisation for the diagnosis of cancer and severe anaemia. For this analysis, three groups of QDU patients (with a final diagnosis of severe anaemia, lymphoma, and lung cancer) were compared with hospitalised patients with the same final diagnoses. **The results showed a significant cost saving of care delivered by a QDU service compared with traditional inpatient care**. The QDU savings from hospitalisation for the three diagnostic groups were related to fixed direct costs of hospital stays (66% of total savings). Savings related to fixed non-direct costs of structural and general functioning were 33% of total savings. Savings related to variable direct costs of diagnostic investigations were 1% of total savings. Overall savings from hospitalisation of all patients were €867,719.31.

#### 2.4.2 Generic economic outcomes derived from quasi-experimental studies

Eleven quasi-experimental studies reported a range of generic economic outcomes including patient costs (such as the mean cost per visit to the diagnostic centre, cost of diagnostic test per patients and the mean costs per patient), costs associated with running a diagnostic centre (such as the average cost per process, staffing costs and direct, indirect and structural costs) and cost savings.

##### Patient costs

###### Cost per visit to the diagnostic centre

Nine studies covering three different diagnostic centres (two from Spain and one from the UK) reported the mean cost per diagnostic centre visit. The findings from the most recent studies are reported below for each diagnostic centre. Two studies reported findings from the same diagnostic centre (Bosch et al 2020; Bosch et al 2018), however Bosch et al 2018 combined the cohort of patients in this centre with another diagnostic centre to form a single unit, and as such both studies are reported below. The findings were generally unclear.

- Bosch et al (2020) compared the costs of outpatients (QDU patients) and inpatients and found **the cost per hospital stay was less expensive than cost per visit to the QDU at the same hospital** (€154.72 vs €340.90).
- Bosch et al (2018) compared the associated costs of outpatient (QDU) and inpatient (hospitalised) settings and found **the total cost per one day stay as an inpatient was less expensive than the cost per visit to outpatients** (€328.42 vs €432.05). However, it is worth noting that two QDUs were combined and analysed as a single unit in this study.
- Porter et al (2003) compared a newly established DDC with two existing clinical settings in the management of MS. They found that **the cost per appointment to the DDC was £395, compared to £95 per visit to another outpatient clinic. For inpatients, the length of stay ranged from one to five days and patients in this group also attended outpatient appointments on two to five occasions before receiving a definite diagnosis. The collective price of admission and outpatient visits ranged from £1940 to £2700.**

###### Cost of diagnostic test per patient

Three studies covering three diagnostic centres (two from Spain and one from the UK) reported the cost of diagnostic tests per patient. Two studies reported findings from the same diagnostic centre (Bosch et al 2020; Bosch et al 2018), however the latter combined the cohort of patients from this centre with another diagnostic centre to form a single unit, and as such both studies are reported below. The findings indicate costs of diagnostic tests may be cheaper in diagnostic centres situated within hospital grounds. Findings for mobile-based diagnostic services are unclear.

- Bosch et al (2020) compared the costs incurred by outpatient (QDU) and inpatient (hospitalised) settings and reported **the total cost of diagnostic examinations per patient to be significantly cheaper for patients attending the QDU** compared to hospitalised patients (€231.88 vs €280.60; p<0.001).
- Bosch et al (2018), compared the associated costs of outpatient (QDU) and inpatient (hospitalised) settings and found **the total cost of diagnostic examinations per patient was significantly cheaper for patients attending the QDUs** compared to hospitalised patients (€713.19 vs €1,026.80; p<0.001). Two QDUs were combined and analysed as a single unit in this study.
- Pallan et al (2005) compared a primary care-based mobile diagnostic ultrasound service to a NHS Trust diagnostic ultrasound service and found **the cost of diagnostic tests was more expensive per patient attending the mobile diagnostic ultrasound service in the community compared to the NHS Trust hospital service** (cost per ultrasound £30 vs £20.62 - £27.51 respectively).

###### Total cost per patient

Three studies conducted in Spain reported on the total cost per patient for two diagnostic centres. The findings from the most recent studies are reported below for each diagnostic centre. Two studies reported findings from the same diagnostic centre (Bosch et al 2020; Bosch et al 2018), however the latter combined the cohort of patients from this centre with another diagnostic centre to form a single unit, and as such both studies are reported below. Both studies identified statistically significant reductions in cost per patient attending a QDU compared to hospitalisation.

- Bosch et al (2020), reported that **the total cost per patient at a QDU was significantly less than the total cost per hospitalised patient** (€347.76 [SD 48.69] vs €634.36 [SD 80.56]; p<0.001). Total cost per QDU patient included 66.7% being attributable to diagnostic tests, 18.2% to ambulatory visits, and 13.7% to salaries, and total cost per hospitalised patient included 46.4% being attributable to personnel salaries and 44.2% to diagnostic tests.
- Bosch et al (2018) reported that **the total cost per patient was significantly less for outpatients (QDU patients) than the cost per hospitalised patient** (€1,408.48 [197.32] vs €4,039.56 [513.02]; p<0.001). Two QDUs were combined and analysed as a single unit in this study.

##### Costs of running a diagnostic centre

###### Average cost per process (from admission to discharge)

Five studies covering one diagnostic centre reported the average cost per process. All five studies were conducted in Spain. The findings of the most recent study is reported below.

- Bosch, Jordán and López-Soto (2013) compared patients attending a QDU with hospitalised patients and found **the average cost per process was more expensive for hospitalised patients compared to patients attending the QDU** (€3,241.11 [standard deviation (SD), 915] vs €726.47 [SD, 617] respectively).

###### Direct, indirect and structural costs

Two studies conducted in Spain (Brito-Zerón et al 2014; Bosch et al 2020) reported on the direct, indirect and structural costs of one diagnostic centre. Therefore, only the findings from the most recent study is reported below.

- Bosch et al (2020) described mean **non-direct costs per patient** in QDU and hospitalised patients. These costs mainly corresponded to structural and general functioning costs such as costs related to maintenance (€0.05 [SD, 0.01] vs €0.66 [SD, 0.07]), laundry (€0.26 [SD, 0.02] vs €5.17 [SD, 0.36]), cleaning services (€0.73 [SD, 0.05] vs €8.36 [SD, 0.67]), administrative costs (€0.02 [SD, 0.01] vs €0.37 [SD, 0.03]), as well as depreciation of fixed costs (€3.10 [SD, 0.61] vs €4.43 [SD, 0.89]). All were found to **cost significantly less for patients attending the QDU** compared to hospitalised patients (p<0.001).

###### Staffing costs

Two studies conducted in Spain covering two diagnostic centres, reported on staffing costs. The two studies covered the same diagnostic centre (Bosch 2020; Bosch 2018) however one study combined the findings for this centre with another diagnostic centre and as such, both studies are reported below. Both studies identified a statistically significant reduction in staff costs compared to controls.

- Bosch et al (2020) reported that **the mean staff wages per patient attending the QDU was significantly lower than the staff wages for hospitalised patients** (€47.71 [SD, 4.15] vs €294.50 [SD, 18.26] respectively; p<0.001).
- Bosch et al (2018) reported that **the mean staff wages per patient attending the QDU was significantly lower than the staff wages for hospitalised patients** (€262.50 [SD,22.92] vs €2,806.49 [SD,174]; p<0.001). Two QDUs were combined and analysed as a single unit in this study.

###### Cost saving from hospitalisation

One study conducted in Spain reported on the cost saving from hospitalisation.

- Bosch et al (2018) compared associated costs of outpatient (QDU) and inpatient (hospitalised) settings and found that **the total cost saving from hospitalisation** was €2,631.08 per patient. However, two QDUs were combined and analysed as a single unit in this study.

#### 2.4.3 Bottom line results for the economic effectiveness of diagnostic centres

There is evidence to suggest that diagnostic centres are cost-saving and a more cost-effective resource than traditional inpatient care. However, it appears that overall cost-effectiveness may be dependent on whether or not the diagnostic centre is running at full capacity. Factors that could determine the costs incurred by a diagnostic centre include the diagnostic and clinical complexity of the patients managed at the unit, as well as the characteristics of the unit including the number of staff and contribution of staff time. Additionally, there is evidence to suggest that utilisation of diagnostic centres can reduce staffing costs, costs incurred per patient, and the costs of diagnostic tests. However, the methodological limitations across included studies and variations in healthcare systems of the countries of origin of included studies could limit the applicability of these findings.

## 3. DISCUSSION

### 3.1 Summary of the findings

Our rapid review sought to identify community diagnostic centres, however our search only identified one diagnostic service located within the community, while the remaining studies covered diagnostic centres located in hospitals with direct access from primary care. There is evidence to suggest that diagnostic centres can be an effective alternative model of care, capable of reducing waiting times, and reducing pressure on hospitals by avoiding hospitalisations, reducing the number of visits required to receive a definite diagnosis, and increasing the number of patients in whom a definite outcome is reached. However, the costs incurred by a diagnostic centre can be impacted by the diagnostic and clinical complexity of the patients managed at the unit, as well as the characteristics of the unit including the number of staff and contribution of staff time. Overall cost-effectiveness of diagnostic centres may be dependent on whether or not the centre is running at full capacity.

However, much of the evidence was derived from quasi-experimental studies, with only three economic evaluations. We identified one RCT, however this study did not contribute to the evidence base used in this review because it was old and primarily focussed on patient anxiety. While it did report some economic outcomes these were superseded by more recent studies. Considerable methodological limitations across included studies, as well as structural differences in healthcare systems across international studies, could limit the applicability of these findings. However, we did identify five studies that were conducted in the UK, three of which were specific to cancer diagnosis, one specific to the diagnosis of MS, and one assessed the effectiveness of a mobile diagnostic ultrasound service.

### 3.2 Limitations of the available evidence

This RR has highlighted several evidence gaps including the paucity of robust study designs in this area of research. The majority of study designs included in this review utilised weak methodologies that may not be appropriate for inferring effectiveness. This RR did not identify any studies exploring equity of access to diagnostic centres and only three economic evaluations were identified, one of which was conducted in the UK.

The quality of reporting in the included studies was oftentimes poor. Key details pertaining to outcome measures or information about diagnostic centres, were often lacking or poorly described. In addition, key statistical parameters, such as confidence intervals, were not reported in some study results, making it difficult to determine the magnitude of effect of some diagnostic centres.

Most of the evidence identified in this review were derived from diagnostic centres located within hospital sites. Only one study included in this review examined a diagnostic service located within the community (a mobile ultrasound service). Whilst siting a diagnostic centre within the hospital setting is likely to provide greater availability to already established and functioning diagnostic equipment and services, it may not be accessible to certain groups of patients, and further worsen health inequalities.

Only five UK studies were identified, with the remaining studies conducted in a variety of other countries. A large number of these studies were conducted in Spain, and these measured many of the relevant outcomes included in this RR. As a result, the generalisability of our findings to Wales could be limited due to differences in healthcare systems and healthcare provision between both countries.

Furthermore, many of the studies conducted in Spain reported data from the same set of diagnostic centres - some with similar data collection periods, thereby creating the potential for double counting (see Appendix 2 for details about the potential for data overlap). To reduce the likelihood of double counting, where multiple studies reported on the same diagnostic centre, we only reported findings from the most recent study in the narrative synthesis (if a comparison to usual care was provided) and highlighted whether any of the other studies identified any other relevant outcomes not reported in the more recent publication. Although this is not a usual approach to take, we believe this was a pragmatic approach, given the timescale.

The majority of included studies compared patients attending diagnostic centres with a range of comparators including hospitalised patients and historical controls. These comparisons may not be appropriate considering the fact that hospitalised patients are generally more acutely unwell and require more clinical input and longer care than those eligible to attend a diagnostic centre. In addition, unlike hospital wards, diagnostic centres are generally not open 24 hours a day, and the infrastructure and operational functioning of these centres were inconsistently reported.

### 3.3 Implications for policy and practice

This RR has highlighted possible benefits of diagnostic centres, particularly with regards to their impact on waiting times and pressure on secondary care. Although inferences around the effectiveness of community diagnostic centres cannot be made due to the paucity of evidence from diagnostic centres located outside of hospital settings, the information extracted from these studies provide valuable information into the potential benefits of establishing these centres within Wales.

In light of the paucity of robust evidence, further well-designed, higher quality research from the UK and similar countries is needed to better understand the effectiveness of community diagnostic centres within Wales. Research around diagnostic centres sited outside of hospital locations is particularly needed to investigate the impact on equity of access. In addition, further research is required to evaluate the effectiveness of diagnostic centres for conditions other than cancer, and full economic evaluations of these centres are also needed to better understand how diagnostic centres can be efficiently utilised.

Comparative impact evaluations should be incorporated into service development plans from the onset, to assess the effectiveness of newly opened diagnostic centres in the UK over time.

### 3.4 Strengths and limitations of this rapid review

The studies included in this RR were identified through an extensive search of electronic databases, trial registries, grey literature, as well as consultation of content experts in the field. Despite making every effort to capture all relevant publications and reduce the risk of bias, it is possible that additional eligible publications may have been missed or we may have introduced some biases to this RR through our inclusion criteria. Efforts were made to reduce this risk of introducing bias and have highlighted this where possible, for example in the investigation into the potential risk of multiple studies reporting the same data.

We identified a large number of studies during the initial stages of this review (prior Rapid Evidence Map), many of which were descriptive in nature. To overcome this potential limitation, we made the decision to include only studies that had some sort of comparator and utilised a study design algorithm developed by Leatherdale (2019), to assign an accurate description of the methods employed.

We made the pragmatic decision to report the findings from the most recent studies when multiple studies were reporting on the same diagnostic centre and outcome. However, this may have introduced some bias in the reporting as there is a potential that the studies could have had different aims and objectives and may also have been focussed on different health conditions. Therefore, the findings may have differed between studies, specifically for the economic evaluations.

As no date or country of study limits were set, and the data collection dates of included studies are wide ranging, it is possible that the diagnostic centres we have included here may not be the same as the proposed diagnostic centres within Wales. It is also possible that as many of the diagnostic centres included were from other countries where the healthcare system is different to that of the UK, the results may not be generalisable to the UK.

## Data Availability

All data produced in the present study are available upon reasonable request to the authors

## Abbreviations

2WW: Two-Week Wait
COVID-19: Coronavirus Disease 2019
CMV: Cytomegalovirus
CNS: Clinical Nurse Specialist
CT: Computed Tomography
DDC: Demyelinating Disease Diagnostic Clinic
DES: Discrete-Event Simulation
D.F.: Degrees Of Freedom
EBV: Epstein-Barr Virus
ED: Emergency Department
EUS: Upper Gastrointestinal Endoscopic Ultrasound
FDG-PET: Fluorodeoxyglucose-Positron Emission Tomography
FNA: Fine-Needle Aspiration
FUN: Fever of Uncertain Nature
HCSW: Healthcare Support Worker
JBI: Joanna Briggs Institute
LRDC: Lymphoma Rapid Diagnosis Clinic
MS: Multiple Sclerosis
PHC: Primary Healthcare Centres
RABC: Rapid Access Breast Clinic
RADS: Rapid Diagnosis And Support
RCT: Randomised Controlled Trial
RDC: Rapid Diagnostic Centre/clinic
RDU: Rapid Diagnostic Unit
REM: Rapid Evidence Map
RO: Radiation Oncology
RR: Rapid Review
RT: Radiotherapy Treatment
SD: Standard Deviation
TAC: Technical Advisory Cell
TS: Traditional System
QALYs: Quality Adjusted Life Years
QDU: Quick Diagnostic Unit
UHN: University Health Network
UK: United Kingdom
WCEC: Wales COVID-19 Evidence Centre

## 5. RAPID REVIEW METHODS

### 5.1 Eligibility criteria

We searched for primary sources to answer the review question “What is the effectiveness of community diagnostic centres?”

The following eligibility criteria were used to identify studies for inclusion in the RR:

**Table 2:** Eligibility criteria

|  | **Inclusion criteria** | **Exclusion criteria** |
| --- | --- | --- |
| **Participants** | Symptomatic patients, all conditions being referred to diagnostic centres via primary care settings |  |
| **Settings** | Diagnostic centres in any setting | Exclude screening programmes, or where there is treatment undertaken but no diagnostics. |
| **Intervention / exposure** | Diagnostic centres/units/hubs and clinics accepting referrals from primary care (as a minimum) | Community diagnostic centres accepting referrals exclusively from other routes |
| **Comparison (if applicable)** | Usual care/other diagnostic centres |  |
| **Outcomes** | All outcomes, with a focus on:<br>- capacity<br>- pressure on secondary care<br>- waiting times<br>- equity of access<br>- and all economic outcomes |  |
| **Study design** | Any design that contains a comparison that can infer effectiveness |  |
| **Countries** | All countries |  |
| **Language of publication** | Studies published in English | Any study not published in English |
| **Publication date** | No date limits set |  |
| **Publication type** | Published and preprint primary literature | All publication types other than primary literature |

### 5.2 Literature search

The studies included in this RR were identified through the literature search conducted in our preliminary work. COVID-19 specific and general repositories of evidence reviews noted in our resource list were searched on 6_th_ July 2022 by three reviewers and an updated search was conducted on the 3_rd_ of August 2022 for the REM. An audit trail of the search process is provided within the resource list (Appendix 3). Searches were limited to English-language publications and included searches for primary studies. References of secondary sources identified during preliminary work were scanned for relevant primary studies and forward and backward citation tracking was also conducted on the secondary sources.

Search concepts and keywords around diagnostic units, centres, hubs, and clinics were utilised. The searches combined free text words and descriptors when available. We deliberately kept our search strategy broad to capture as much evidence on diagnostic centres as possible. Resources searched are outlined in Appendix 3 and the search strategy used to search Medline is available in Appendix 4.

### 5.3 Study selection process

Our prior Rapid Evidence Map included 50 primary studies, all of which were screened for inclusion in the Rapid Review using the updated eligibility criteria in 5.1 by two independent reviewers.

### 5.4 Data extraction

One researcher performed the data extraction and a second researcher carried out consistency checks. Information extracted includes:

- Reference (author, year, country)
- Study design
- Intervention / comparator
- Aim
- Data collection methods (and dates)
- Outcome(s) measured
- Study participants (e.g., sample size, age range, sex, any other specifics)
- Setting
- Staffing/facilities
- Services provided (e.g. MRI, ultrasound, etc.)
- Key findings
- Additional notes/comments

An observations/notes column was added to report key information that was not captured above and to record any limitations of the included sources (see Table 1).

### 5.5 Study design categorisation

Studies were categorised by research design and additional analytic techniques if applicable, using the study design classification system developed by Leatherdale (2019) by a single reviewer, with verification of all judgements by a second reviewer (see Tables 3, 4 and 5). The Leatherdale tool includes a series of questions on the methodological characteristics of a natural experimental study to identify an accurate description of the research design, in particular the characterisation of designs by the frequency and points at which data is collected.

### 5.6 Quality appraisal

A range of JBI quality appraisal checklists (which were selected based on the study design used) were used to assess the methodological quality of each included study. Quality assessment was undertaken by a single reviewer, with verification of all judgements by a second reviewer. Any discrepancies were discussed and resolved amongst the review team. The results of the quality appraisals can be seen in Tables 3, 4, and 5.

### 5.7 Synthesis

The effectiveness of diagnostic centres was compared to a comparator (most often usual care) on a range of outcomes including, pressure on secondary care, capacity, wait times and cost-effectiveness. Following on from the REM, a narrative synthesis was conducted reporting the results of selected studies that included diagnostic centres accepting referrals from primary care and that had a comparative element. Stakeholders highlighted the importance of finding out if diagnostic centres can impact waiting times and current pressures on secondary care, as well as the economic impact. With this in mind, we categorised the outcomes identified into ‘impact on waiting times’, ‘impact on pressure’ and ‘economic outcomes’ and reported findings using these categories. In an attempt to highlight the more robust methodological studies (economic evaluations), we reported these findings first. Where multiple studies reported outcomes on the same diagnostic centre, we chose to report only the findings form the most recent study to avoid the risk of double counting.

## 6. EVIDENCE

### 6.1 Study selection

Due to the large number of studies identified it was decided, in conjunction with stakeholders, to include only studies that included a diagnostic centre accepting referrals from primary care as a minimum and included a comparator group. Of the initial 50 studies screened 42 included diagnostic centres that accepted referrals from primary care and of these, 21 were comparative and 20 reported on our outcomes of interest. The study selection process is outlined below.

### 6.2 Study selection flow chart

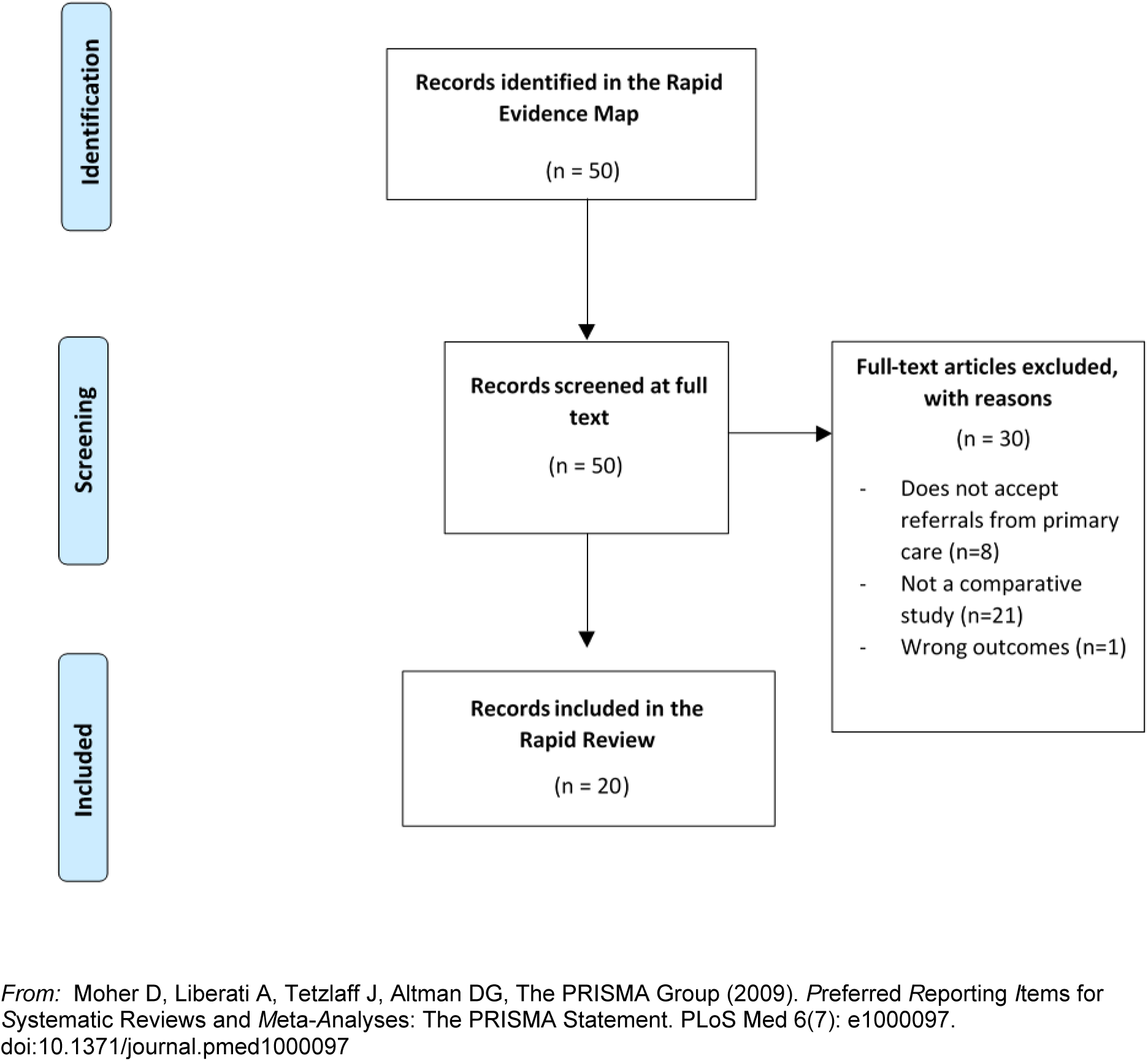

### 6.3 Quality appraisal tables

**Table 3.** Quality appraisal results for economic evaluations

| Study | JBI Appraisal Items – Economic evaluations |  |  |  |  |  |  |  |  |  |  |
| --- | --- | --- | --- | --- | --- | --- | --- | --- | --- | --- | --- |
|  | Q1 | Q2 | Q3 | Q4 | Q5 | Q6 | Q7 | Q8 | Q9 | Q10 | Q11 |
| Bosch et al 2021 | Y | Y | Y | U | Y | Y | Y | U | N | Y | N |
| Sanclemente-Ansó et al 2016 | Y | Y | Y | Y | Y | Y | Y | U | N | Y | N |
| Sewell et al 2020 | Y | Y | Y | Y | Y | Y | N | N | Y | Y | N |

Key: Y=Yes, N=No, U=Unclear, N/A=not applicable
Q1. Is there a well-defined question?
Q2. Is there comprehensive description of alternatives?
Q3. Are all important and relevant costs and outcomes for each alternative identified?
Q4. Has clinical effectiveness been established?
Q5. Are costs and outcomes measured accurately?
Q6. Are costs and outcome valued credibly
Q7. Are costs and outcomes adjusted for differential timing?
Q8. Is there an incremental analysis of costs and consequences?

**Table 4.**
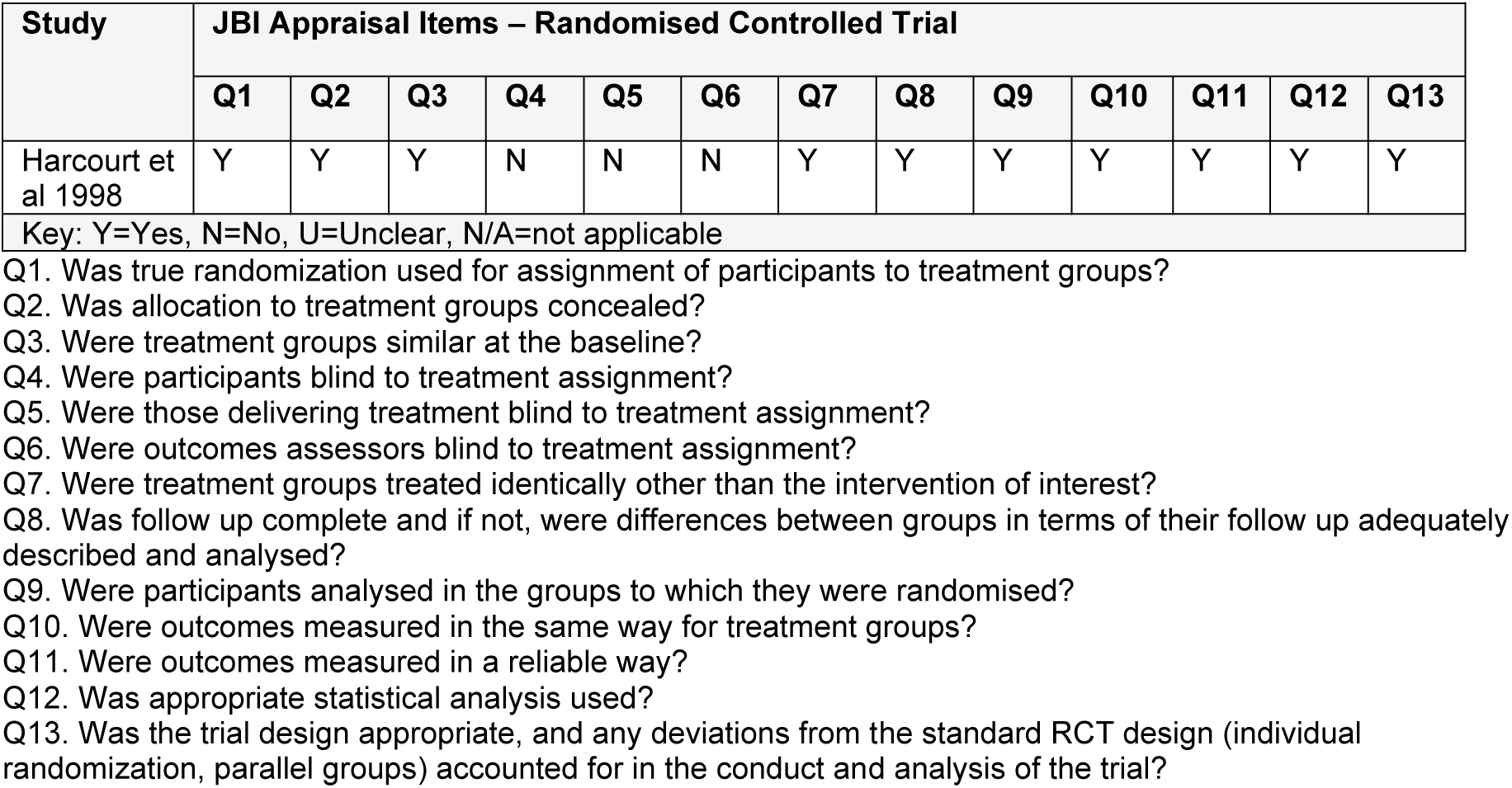
Quality appraisal results for randomised controlled trials

**Table 5.** Quality appraisal results for quasi-experimental studies

| Study | JBI Appraisal Items – Quasi-experimental studies |  |  |  |  |  |  |  |  |
| --- | --- | --- | --- | --- | --- | --- | --- | --- | --- |
|  | Q1. | Q2. | Q3. | Q4. | Q5. | Q6. | Q7. | Q8. | Q9. |
| Arnaout et al 2013 | Y | U | Y | Y | N | N/A | Y | Y | Y |
| Bosch et al 2011 | Y | U | Y | Y | N | N/A | Y | Y | N/A |
| Bosch et al 2012a | Y | N | Y | Y | N | N/A | N | Y | Y |
| Bosch et al 2012b | Y | N | Y | Y | N | N/A | Y | Y | Y |
| Bosch et al 2012c | Y | N | Y | Y | N | N/A | N | Y | Y |
| Bosch, Jordán and López-Soto 2013 | Y | N | Y | Y | N | N/A | N | Y | Y |
| Bosch et al 2018 | Y | N | Y | Y | N | N/A | Y | Y | Y |
| Bosch et al 2020 | Y | N | Y | Y | N | N/A | Y | Y | Y |
| Brito-Zerón et al 2014 | Y | N | Y | Y | N | N/A | N | Y | Y |
| Choudhury et al 2013 | Y | U | Y | Y | N | N/A | Y | Y | Y |
| McKevitt et al 2017 | Y | U | Y | Y | N | N/A | Y | Y | Y |
| Montori-Palacín et al 2017 | Y | N | Y | Y | N | N/A | Y | Y | Y |
| Nixon et al 2019 | Y | U | Y | Y | N | N/A | Y | Y | Y |
| Pallan et al 2005 | Y | U | Y | Y | N | N/A | Y | U | U |
| Porter et al 2003 | Y | N | Y | Y | N | N/A | Y | U | N/A |
| Sethukavalan et al 2013 | Y | Y | Y | Y | N | N/A | Y | Y | Y |
| Key: Y=Yes, N=No, U=Unclear, N/A=not applicable |  |  |  |  |  |  |  |  |  |

1. Is it clear what is the cause and what is the effect? 2. Were the participants included in any comparisons similar? 3. Were the participants included in any comparisons receiving similar treatment/care, other than the exposure or intervention of interest? 4. Was there a control group? 5. Were there multiple measurements of the outcome both pre and post the intervention/ exposure? 6. Was follow up complete and if not, were differences between groups in terms of their follow up adequately described and analysed? 7. Were the outcomes of participants included in any comparisons measured in the same way? 8. Were outcomes measured in a reliable way? 9. Was appropriate statistical analysis used?

## 7. ADDITIONAL INFORMATION

### 7.2 Information available on request

The protocol for this RR is available on request from Hannah Shaw, Public Health Wales, **E-mail:**.

### 7.3 Acknowledgements

The authors would like to thank Brendan Collins, Delia Ripley, Jennifer Morgan, Joanna Charles, Leon Wong, Rob Orford and Sally Anstey for their contributions during stakeholder meetings in guiding the focus of the review and interpretation of findings.

## 8. ABOUT THE WALES COVID-19 EVIDENCE CENTRE (WCEC)

The WCEC integrates with worldwide efforts to synthesise and mobilise knowledge from research.

We operate with a core team as part of Health and Care Research Wales, are hosted in the [Wales Centre for Primary and Emergency Care Research (PRIME)](http://www.primecentre.wales/), and are led by [Professor Adrian Edwards of Cardiff University](https://www.cardiff.ac.uk/people/view/123022-edwards-adrian).

The core team of the centre works closely with collaborating partners in [Health Technology Wales](https://healthtechnology.wales/), Wales Centre for Evidence-Based Care, [Specialist Unit for Review Evidence centre](https://www.cardiff.ac.uk/specialist-unit-for-review-evidence), [SAIL Databank](https://healthandcareresearchwales.org/about-research-community/secure-anonymised-information-linkage-sail-databank), Bangor Institute for Health & Medical Research/ Health and Care Economics Cymru, and the [Public Health Wales Observatory](https://phw.nhs.wales/services-and-teams/observatory/).

Together we aim to provide around 50 reviews per year, answering the priority questions for policy and practice in Wales as we meet the demands of the pandemic and its impacts.

### Director

Professor Adrian Edwards

### Website

https://healthandcareresearchwales.org/about-research-community/wales-covid-19-evidence-centre

## 9. APPENDIX

## APPENDIX 1: Characteristics of included diagnostic centres

| Reference | Location & setting | Aim of the centre | Staff & facilities | Investigative procedures/services conducted | Diagnosis of interest | Referral | Referral criteria | Key underpinning elements of diagnostic centre |
| --- | --- | --- | --- | --- | --- | --- | --- | --- |
| Canada |  |  |  |  |  |  |  |  |
| *Lymphoma Rapid Diagnosis Clinic (LRDC) at Princess Margaret Cancer Centre, Toronto, Ontario* |  |  |  |  |  |  |  |  |
| Nixon et al (2020). | Lymphoma **Rapid diagnosis** clinic (LRDC) **based in a tertiary cancer centre** (Princess Margaret Cancer Centre), part of University Health Network (UHN), Toronto, Ontario, Canada. | The goal of the centre was to provide **specialised comprehensive assessment and timely and appropriate investigations** to reduce wait times to a **definitive biopsy** to establish histologic diagnosis and initiation of treatment. | The clinic was **led by a Nurse practitioner**; however, it appears weekly dedicated operating rooms for lymphoma biopsies were used by head and neck, thoracic, and general surgeons, and biopsy material from outside facilities were reviewed by hemapathologists when available. | Laboratory tests, peripheral blood flow cytometry, tuberculosis skin testing, abdominal ultrasound, computed tomography scans, bone marrow biopsy, or FNA. Referral to surgical services for consideration of excisional lymph node biopsy or radiology for image guided core biopsy was requested based on location and size of lymphadenopathy. | Lymphadenopathy and suspected lymphoma | Primary care or medicine, ER, or community specialists | Lymphadenopathy on the basis of **clinical assessment or imaging, biopsy results suspicious** for lymphoma, or peripheral blood abnormalities.<br>Patient symptoms included: symptoms of viral infection, "B" symptoms, new pain, pruritus, palpable lymph nodes, lymphocytosis (ALC > 4.0 X 10<sup>9</sup>/L) | The clinic was Nurse practitioner-led<br>The optimal approach to lymphoma diagnosis is multidisciplinary, and our data suggest that even a modest additional investment in personnel and resources would be expected to significantly improve delivery of care for patients with suspected lymphoma. |
| *The Gale and Graham Wright Prostate Centre, North York General Hospital Toronto, Ontario* |  |  |  |  |  |  |  |  |
| Sethukavalan et al (2013). | **Rapid diagnostic** unit (RDU) called The Gale and Graham Wright Prostate Centre, set up at the North | The Wright Prostate Centre is a diagnostic assessment programme and **multidisciplinary clinic where both radiation oncology and urology specialists see** | Staff include **radiation oncology and urology specialists** | Unclear | Prostate cancer | Urologists or family physicians (primary care) | Patients with **suspicion of cancer** either (based on a **prostate-specific antigen (PSA) test**), because of **symptoms**, or an abnormal **digital rectal exam (DRE)**, which warranted further testing or |  |
|  | York General Hospital (NYGH), Branson Site, Toronto, Ontario, Canada. | **every patient on the same day** after obtaining their **biopsy results** |  |  |  |  | referral to a specialist to definitively diagnose or rule out cancer |  |
| *Rapid Access Diagnostic and Support (RADS) at The Women's Breast Health Center, Ottawa Hospital, Ottawa, Ontario* |  |  |  |  |  |  |  |  |
| Arnaout et al (2013). | The Women's Breast Health Center of the Ottawa Hospital in Ottawa, Ontario, Canada (a university-affiliated tertiary care centre) | The **Rapid Diagnosis** and Support (RADS) Program initiative was a pilot programme which aimed to accommodate the **increasing patient volume, reduce wait times, decrease fragmentation of care**, and enhance a patient's overall experience. | A **multidisciplinary team** of breast cancer specialists (five radiologists, five surgeons, two pathologists, one nurse manager, two nurse navigators, and a diagnostic imaging manager) | Routine screening mammography, initial diagnostic imaging workup (mammogram and/or breast ultrasound) for a breast problem (e.g., palpable mass, breast pain, nipple discharge), or additional diagnostic imaging and biopsy workup following an abnormal mammogram performed at another institution | Breast cancer | Primary care | Patients are routinely referred to the breast centre for either:<br>1. Routine **screening** mammogram<br>2. **Breast symptoms** requiring diagnostic imaging workup<br>3. **Abnormal outside imaging** needing additional diagnostic workup/biopsy | Coordination of diagnostic imaging workup and nursing support were provided by **a nurse navigator**. The nurse navigator was assigned to track all patients within the programme to help expedite and inform them of additional diagnostic imaging workup appointments, provide psychosocial support to the patients during their diagnostic care, and provide a telephone hotline service for which patients can call if they had any questions or concerns.<br>RADS program patients were labelled as such on imaging and pathology requisitions to help avoid delays. Interventions, such as coordinating same day tests and changing biopsy schedules and calling pathologists to accommodate faster access to pathology, were implemented as much as possible.<br>Through this prospective pilot project, we have learned that we can efficiently triage patients at highest risk of having breast cancer and therefore initiate strategies to reduce diagnostic wait times for these patients and prevent fragmentation of care. Most of these interventions were aimed **at tracking the diagnostic journey of these patients so that their workup can be appropriately expedited and unnecessary gaps in scheduling can be avoided.** |
| *Rapid Access Breast Clinic (RABC) at Mount St Joseph Hospital, Vancouver* |  |  |  |  |  |  |  |  |
| McKevitt et al (2017). | **Rapid Access** Breast Clinic (RABC) at Mt St Joseph Hospital, British Columbia, Canada | The RABC was established **to offer a single site for coordinated clinical and radiological assessment** of breast problems | Clerical staff, radiologists, surgeons, nurse navigator and clinic family physician (FP) | Offers on-site mammography, breast ultrasound, ultrasound-guided biopsy, and mammographic and ultrasound-guided fine wire localisation. Patients requiring **stereotactic core biopsy, MRI or MRI-guided biopsy have those investigations coordinated by the RABC at regional imaging sites** offering those investigations. | Breast cancer | Family physician (primary care) | Abnormal **screening** mammogram or presenting with a **new breast problem**. | The Rapid Access Breast Clinic (RABC) was established following the guidelines for breast centres outlined by the European Society of Mastology (EUSOMA).<br>The clinic **provided triple evaluation** of patients with close collaboration between clinicians and radiologists, facilitated by clinical pathways and nurse navigation. The development of the RABC in conjunction with the radiology department at the centre created a unique situation in which the breast surgeons saw patients managed by two separate diagnostic pathways. |
| Spain |  |  |  |  |  |  |  |  |
| *Quick Diagnosis Unit (QDU), Hospital Clínic, Barcelona* |  |  |  |  |  |  |  |  |
| Bosch et al (2020). Brito-Zerón et al (2014). Bosch, Jordán and López-Soto (2013). Bosch et al (2012a). Bosch et al (2012b). Bosch et al (2012c). Bosch et al (2011). | **Quick Diagnosis** Unit (QDU) based in the Adult Day Care Centre which is situated within the General Internal Medicine Department of a public tertiary university hospital (Hospital Clínic), Barcelona, Spain. | The QDUs implemented in Spain are mainly **led by general internists** and aim to provide patients with serious disease with the **prompt and effective diagnosis and treatment** they deserve and require | Staff at QDU includes a full-time consultant internist, a senior internal medicine resident, a full-time nurse, a part-time nurse coordinator, and 2 part-time administrative assistants. The unit is open 5 hours a day, 5 days a week. In addition, consultants from other services are provided as required.<br>It has a consulting room and a waiting room for patients | Upper gastrointestinal endoscopic ultrasound (EUS), US/CT-guided biopsy, contrast-enhanced thin-slice CT scan of the chest, abdomen, and pelvis<br>Laboratory tests included, among others, acute phase reactants (C-reactive protein, erythrocyte sedimentation rate), hemogram (total leucocytes, manual white blood cell count, haemoglobin, haematocrit, platelets), liver function tests, serum lactate dehydrogenase, serum total proteins and protein electrophoresis, microbiological serologies [e.g. IgM and IgG for cytomegalovirus (CMV), | Most studies were non-specific but including potentially severe diseases such as **cancer and severe anaemia** (some studies had a specific focus including severe anaemia, pancreatic cancer, unexplained fever). | Primary care and emergency department | The QDU evaluates stable **patients with suspected serious disease who require expeditious workup** and who, in many instances, would be admitted to hospital for diagnostic testing. Thus, patients must be physically and mentally capable of attending several outpatient appointments.<br>Symptoms may include: Anaemia, cachexia-anorexia syndrome, febrile syndrome, adenopathies and/or | The operation of the QDU is based on an urgent first visit, followed by the preferential scheduling and coordination of complementary tests and subsequent visits until a diagnosis is made. |
|  |  |  | and families, and functions daily. | Epstein–Barr virus (EBV), Toxoplasma gondii, human parvovirus B19], HIV testing, b2 microglobulin, specific serum tumour markers, specific serum autoantibodies, specific genetic studies (autoinflammatory diseases), including cultures, imaging studies, endoscopies and cytology/biopsy studies<br>The main diagnostic tests (analytical and microbiological tests, simple radiology [X-ray], computed tomography [CT], echography, nuclear scintigraphy, digestive endoscopy, biopsies and lymph node fine-needle puncture aspiration [FNPA]) are normally performed within 7 days after the first visit. |  |  | palpable masses, unexplained severe abdominal pain, chronic diarrhoea, recent severe constipation, rectorrhagia, jaundice, lung and/or pleural abnormalities, unexplained dyspnea, dysphagia, ascites, anasarca, bone pain with suspicion of malignancy, arthritis, hemogram abnormalities suggestive of primary hematologic disorder, splenomegaly and/or hepatomegaly, monoclonal paraprotein band with or without suspicion of multiple myeloma, neurologic disorders (central, spinal, and peripheral nervous system) |  |
| *Quick Diagnostic Unit (QDU) at Bellvitge Hospital, Barcelona* |  |  |  |  |  |  |  |  |
| Sanclémente-Ansó et al (2016). | QDU at Bellvitge University Hospital in Barcelona, which is affiliated to the University of Barcelona, Catalonia, | Not stated | The QDU is **internist led** and open 7 hours a day, 2 days a week (Tuesdays and Fridays). Analysis implies staff also included attending physician, | Not explicitly stated, but to include blood and urine analysis, X-ray CT, simple X-ray, PET-CT, biopsy, bronchoscopy, cytology, microbial culture, scintigraphy, mammography, specialist consultation, ultrasonography, colonoscopy, | Non-specific but including **potentially serious diseases** | Primary care and the emergency department | Anaemia, cachexia-anorexia syndrome, febrile syndrome, adenopathies and/or palpable masses, unexplained severe abdominal pain, chronic diarrhoea, recent severe constipation, | Hospital-based QDUs are normally led by internists and are a **distinct model of outpatient care delivery** almost exclusively reported in Spain, most notably in Catalonia. The driving reason explaining the important role of internists leading QDUs is the common presence of **nonspecific symptoms such as weight loss, fatigue, malaise or fever of unknown origin in patients referred to** |
|  | Spain. The QDU is integrated to the Internal Medicine Department of the hospital. |  | registered nurse, caretaker. | electrocardiography, lower and upper gastrointestinal series, esophagogastroduodenoscopy, blood marrow aspiration, flow cytometry |  |  | rectorrhagia, jaundice, lung and/or pleural abnormalities, unexplained dyspnea, dysphagia, ascites, anasarca, bone pain with suspicion of malignancy, arthritis, hemogram abnormalities suggestive of primary hematologic disorder, splenomegaly and/or hepatomegaly, monoclonal paraprotein band with or without suspicion of multiple myeloma, neurologic disorders (central, spinal, and peripheral nervous system) | **these units. The versatility of these physicians for the diagnosis of a wide range of serious disorders together with their integral, global view of the patient** contrast with the more specialized approach of physicians at other units such as the UK one-stop diagnostic clinics.<br>The requirements for evaluation by the QDU at Bellvitge Hospital are: 1) **clear referral criteria based on a pre-established list of suspected serious disorders**; 2) the first visit has to occur as soon as possible after referral ($\leq 15$ days); 3) patients must have preferential access to a wide range of diagnostic tests; and 4) patients should be able to attend several appointments for outpatient visits and diagnostic tests. |
| *Quick Diagnosis Unit (QDU), Hospital Plató* |  |  |  |  |  |  |  |  |
| Bosch et al (2021). Montori-Palacín et al (2017). | QDU of a second-level district hospital (Hospital Plató) with 160 beds and providing healthcare for a population of 140,000 inhabitants. | The unit works as an **ambulatory clinic** evaluating patients with **suspected severe conditions** whose physical performance allows them to travel from home to hospital and back for visits and examinations. | The unit is staffed with two part-time **general internists** each working 4 hours per week, as well as administrative personnel. | Not described but appear to include CT scan, MRI, ultrasonography, endoscopy, scintigraphy, body FDG-PET, cytology/biopsy, bone marrow aspiration. | **Suspected severe conditions** | Primary care, emergency department, outpatient clinics, and inpatient wards. | Not described in detail, but included unintentional weight loss, adenopathies and/or palpable masses, anaemia, fever, gastrointestinal symptoms, test abnormalities, osteoarticular symptoms, respiratory symptoms, unexplained tiredness, neurological disorders, ascites and other reasons (not defined). | The general working protocol of the unit consists of a **rapid first appointment after referral from primary care** centres or the emergency departments of the hospitals (usually within 5 days), followed by **preferential programming of diagnostics tests and subsequent visits until a diagnosis** is made. |
| United Kingdom |  |  |  |  |  |  |  |  |
| *The Breast Care Centre, Bristol* |  |  |  |  |  |  |  |  |
| Harcourt et al (1998). | **One stop clinic** in a hospital (location not given but likely to be study author's centre - The Breast Care Centre, Frenchay Healthcare Trust, Bristol, UK) | Not stated | All clinics were conducted by the same **two surgeons** (one consultant, one staff grade) | **Triple assessment** (consisting of clinical examination, ultrasound scanning and cytology) and mammography, when needed. | Breast cancer | GP (primary care) | Women with no previous diagnosis of breast cancer, living within reasonable travelling distance of the hospital and whose general practitioner (GP) referral letter stated the presence of a breast lump | Triple assessment of each woman was conducted by either member of staff in the initial appointment. Mammography, when needed, was then carried out by radiologists in the hospital's general X-ray department. Cytology specimens were analysed in the pathology department sited elsewhere in the hospital. This system was constant over the course of the study. |
| *Rapid Diagnostic Clinic (RDC) at St Barts Health NHS Trust, London* |  |  |  |  |  |  |  |  |
| Choudhury et al (2013). | A '**Rapid Diagnostic Clinic**' (RDC) within the ENT department (of Barts Health NHS Trust, London) | The RDC provides **multi-modality specialist assessment** for new referrals, with on-site sonography and cytology. | ENT head and neck specialist, either a consultant or specialist registrar, consultant head and neck radiologist, consultant histopathologist. This clinic is run on a weekly basis. | Ultrasonography, and fine needle aspiration cytology (FNAC) | Head and neck cancer | GPs and other specialities within the hospital | Not clearly stated. (Targeted referrals included patients referred via the 2WW scheme and non-2WW referrals) | The RDC in this study was established based on National Institute of Clinical Excellence (NICE) recommendations for improving healthcare services for head and neck cancers. The RDC is a **one-stop diagnostic service** where all new target referrals for patients with suspected malignancy can receive **multi-modality specialist assessment**. |
| *Demyelinating disease diagnostic clinic (DDC) at University College London* |  |  |  |  |  |  |  |  |
| Porter et al (2003). | Demyelinating disease diagnostic clinic (DDC) at the National Hospital for Neurology and Neurosurgery, University | The DDC aims to **minimize the time between referral and completion of tests** by carrying out tests on the **same day** and | The clinic is staffed by a consultant neurologist and a multiple sclerosis nurse specialist | **Evoked potential (VEP) testing, magnetic resonance imaging (MRI), and blood screening** | **Multiple sclerosis** | GP or another neurologist | GPs tend to refer patients with new **neurological symptoms**, lasting days or weeks with **no obvious explanation**. Neurologists refer **difficult diagnostic** | The DDC was modelled on the UK multiple sclerosis Society standards of healthcare set for the diagnostic phase. The structure of the DDC provides an **appropriate setting and experienced professional support to deal with the initial psychological impact of the diagnosis** and offers ongoing support through patient/family |
|  | College London Hospitals | providing **follow-up within four weeks.** |  |  |  |  | **cases** including suspected primary progressive disease, those with an atypical presentation and those with nonorganic symptoms | health promotion and self-management programmes.<br>The clinic is designed to allow a 45-minute new patient appointment with the consultant neurologist and MS nurse, followed by access to visual evoked potential (VEP) testing, magnetic resonance imaging (MRI) and blood screening, as appropriate. Follow-up appointments to all patients are scheduled to allow 30 minutes with the neurologist and MS nurse followed by a further 30 minutes with the MS nurse in a quiet room. All **newly diagnosed are followed up with one to one appointments with their families** and have access to a structured educational programme entitled 'Working together to understand MS'. |
| *Rapid Diagnostic Centre (RDC) at Neath Port Talbot Hospital, Neath, Wales* |  |  |  |  |  |  |  |  |
| Sewell et al (2020). | Rapid diagnostic centre (RDC), Neath Port Talbot Hospital, Wales, UK | Not stated | Consultant physician, a radiologist, a clinical nurse specialist (CNS), and a healthcare support worker (HCSW). Management and clinical guidance are provided by the RDC coordination manager and GP project lead. Two half-day clinics a week with five available clinic slots | It is unclear if the full range of investigative procedures available at the centre was reported but includes physical examination and computed tomography (CT) | Cancer | GP (primary care) | Patients with **vague and/or non-specific symptoms suspicious of cancer, who do not meet criteria for referral under an urgent suspected cancer (USC) pathway** | This pilot rapid diagnosis centre (RDC) allows GPs within targeted clusters to refer adults with vague and/or non-specific symptoms suspicious of cancer, who do not meet criteria for referral under an urgent suspected cancer (USC) pathway, to a multidisciplinary RDC clinic where they are seen within a week. Patients leave the clinic with either a diagnosis and management plan or further investigations booked. |
| *Community Diagnostic Service throughout the West Midlands* |  |  |  |  |  |  |  |  |
| Pallan et al (2005). | Community based, **mobile diagnostic ultrasound service**, in a primary care area in the West Midlands, England, UK | Not stated | The service is staffed by an **independent radiographer** | Ultrasound scans included abdominal, pelvic, transvaginal, renal, and prostate | Not specified | GP (primary care) | Not stated | The mobile diagnostic ultrasound service was radiographer-led and was provided to GPs by an independent radiographer. |

## Appendix 2: Potential overlap of data for the studies conducted in Spain

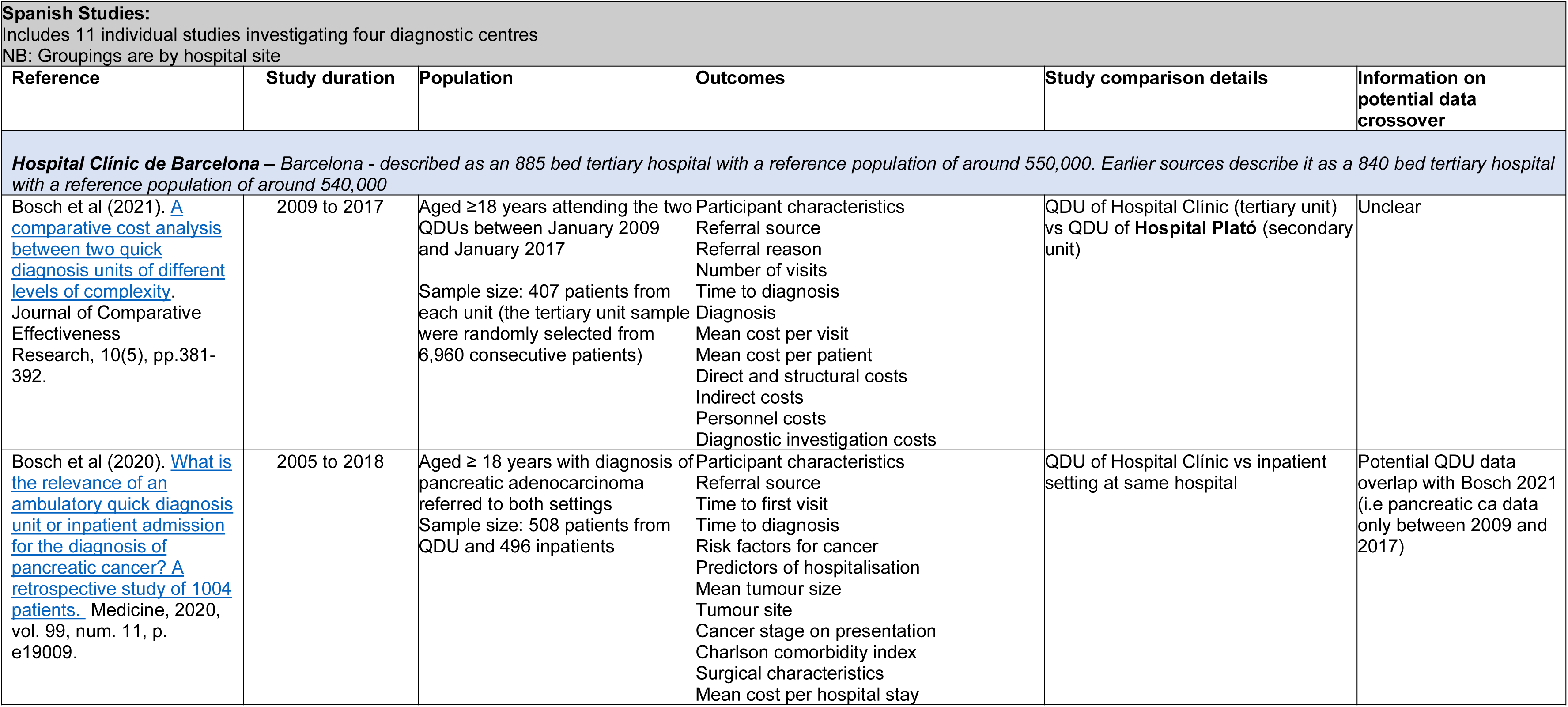
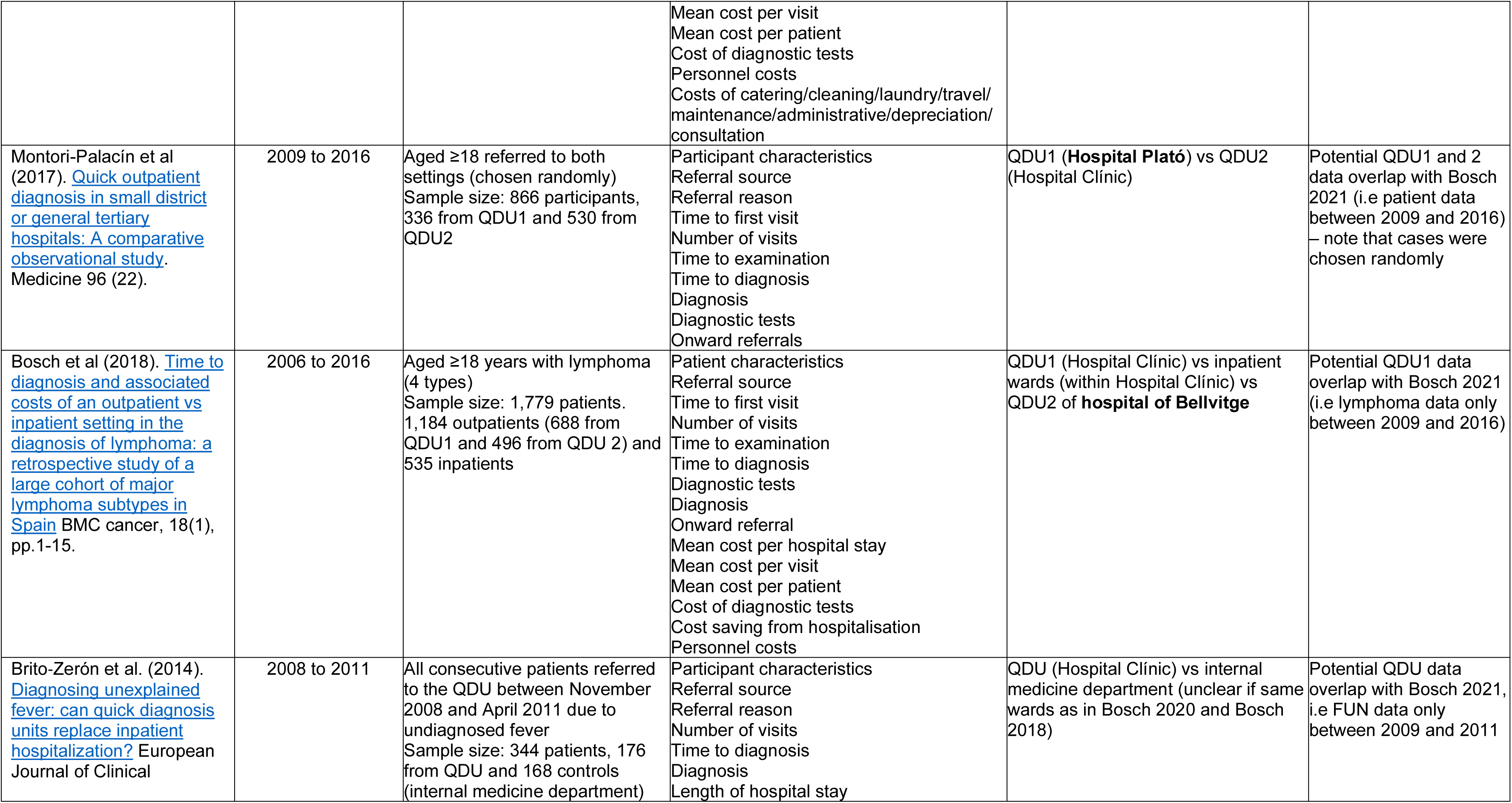
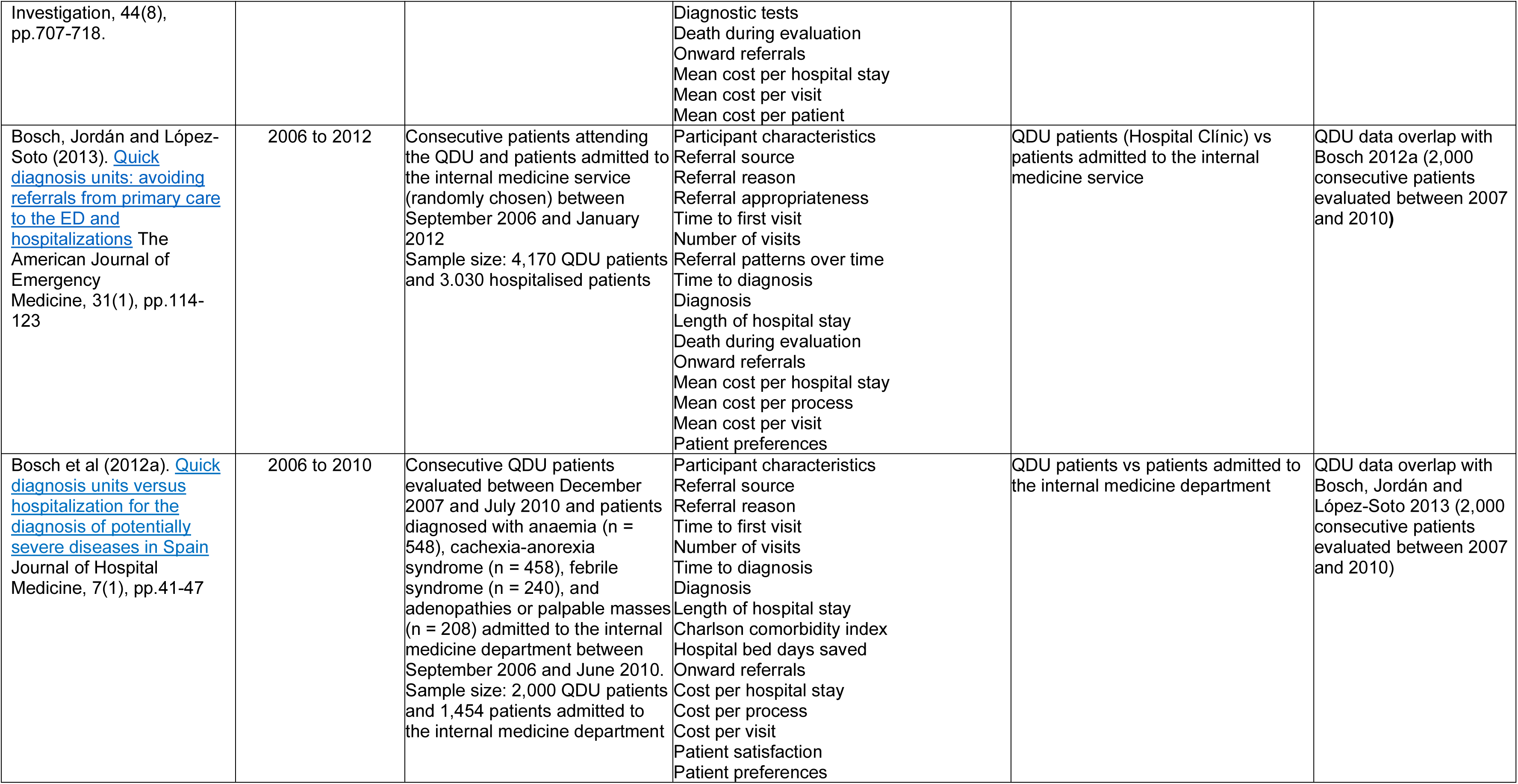
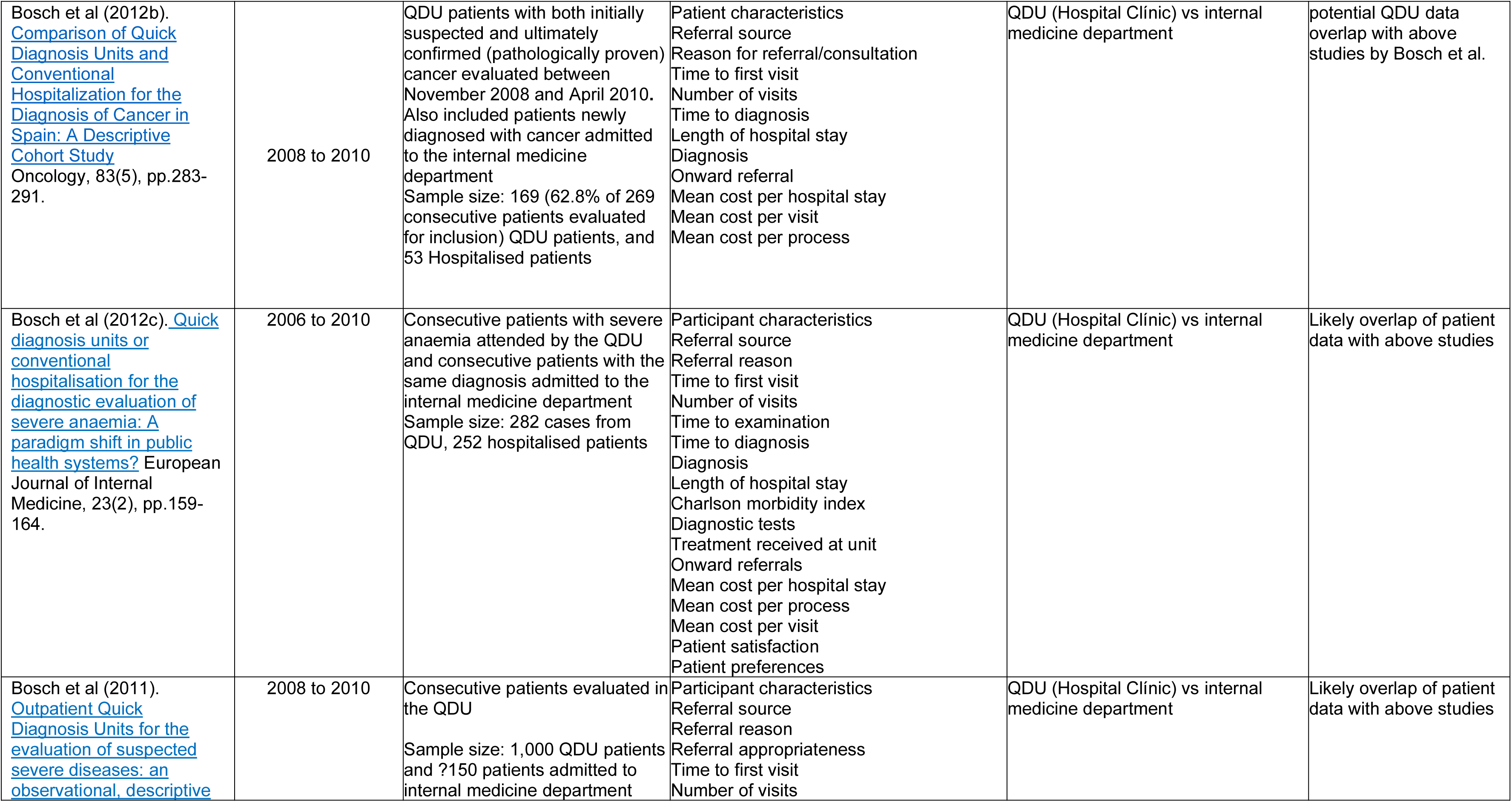
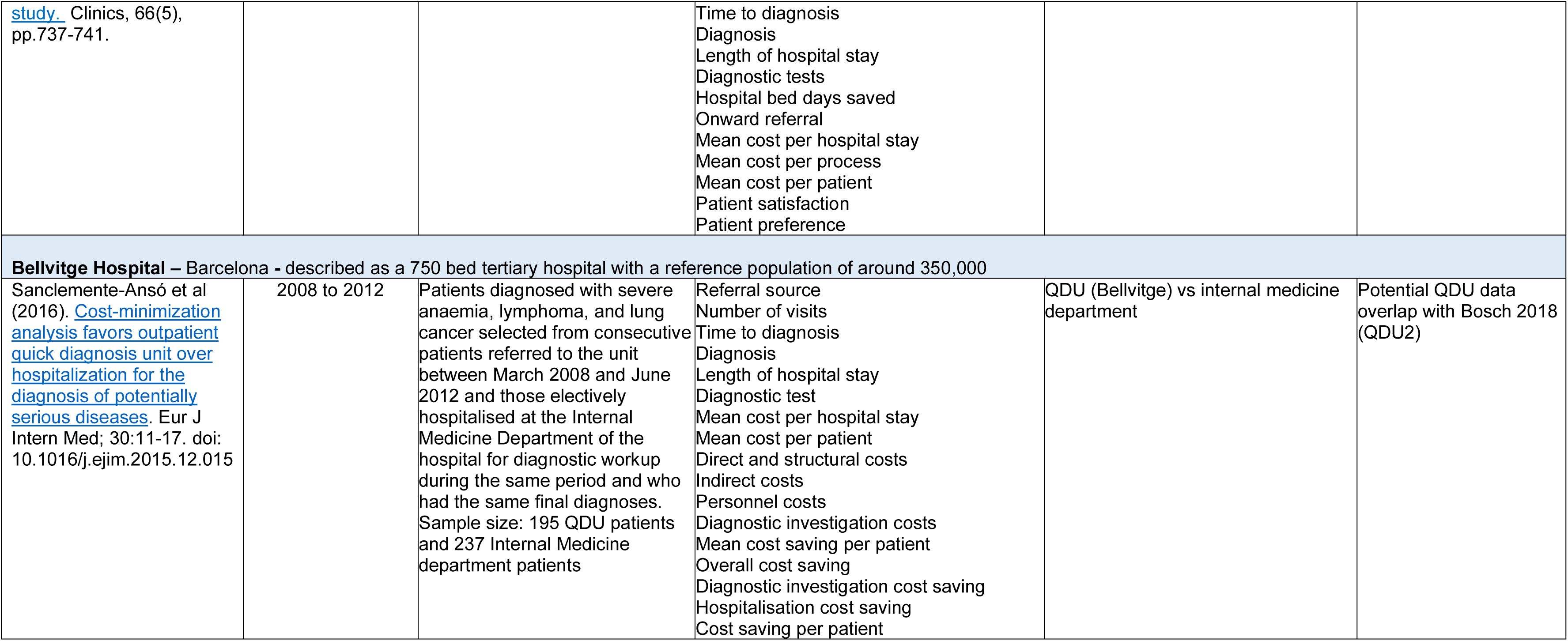

| **Spanish Studies:**<br>Includes 11 individual studies investigating four diagnostic centres<br>NB: Groupings are by hospital site |  |  |  |  |  |
| --- | --- | --- | --- | --- | --- |
| Reference | Study duration | Population | Outcomes | Study comparison details | Information on potential data crossover |
| **Hospital Clínic de Barcelona** – Barcelona - described as an 885 bed tertiary hospital with a reference population of around 550,000. Earlier sources describe it as a 840 bed tertiary hospital with a reference population of around 540,000 |  |  |  |  |  |
| Bosch et al (2021). A comparative cost analysis between two quick diagnosis units of different levels of complexity. Journal of Comparative Effectiveness Research, 10(5), pp.381-392. | 2009 to 2017 | Aged ≥18 years attending the two QDUs between January 2009 and January 2017<br>Sample size: 407 patients from each unit (the tertiary unit sample were randomly selected from 6,960 consecutive patients) | Participant characteristics<br>Referral source<br>Referral reason<br>Number of visits<br>Time to diagnosis<br>Diagnosis<br>Mean cost per visit<br>Mean cost per patient<br>Direct and structural costs<br>Indirect costs<br>Personnel costs<br>Diagnostic investigation costs | QDU of Hospital Clínic (tertiary unit) vs QDU of **Hospital Plató** (secondary unit) | Unclear |
| Bosch et al (2020). What is the relevance of an ambulatory quick diagnosis unit or inpatient admission for the diagnosis of pancreatic cancer? A retrospective study of 1004 patients. Medicine, 2020, vol. 99, num. 11, p. e19009. | 2005 to 2018 | Aged ≥ 18 years with diagnosis of pancreatic adenocarcinoma referred to both settings<br>Sample size: 508 patients from QDU and 496 inpatients | Participant characteristics<br>Referral source<br>Time to first visit<br>Time to diagnosis<br>Risk factors for cancer<br>Predictors of hospitalisation<br>Mean tumour size<br>Tumour site<br>Cancer stage on presentation<br>Charlson comorbidity index<br>Surgical characteristics<br>Mean cost per hospital stay | QDU of Hospital Clínic vs inpatient setting at same hospital | Potential QDU data overlap with Bosch 2021 (i.e pancreatic ca data only between 2009 and 2017) |
|  |  |  | Mean cost per visit<br>Mean cost per patient<br>Cost of diagnostic tests<br>Personnel costs<br>Costs of catering/cleaning/laundry/travel/maintenance/administrative/depreciation/consultation |  |  |
| Montori-Palacín et al (2017). Quick outpatient diagnosis in small district or general tertiary hospitals: A comparative observational study. Medicine 96 (22). | 2009 to 2016 | Aged ≥18 referred to both settings (chosen randomly)<br>Sample size: 866 participants, 336 from QDU1 and 530 from QDU2 | Participant characteristics<br>Referral source<br>Referral reason<br>Time to first visit<br>Number of visits<br>Time to examination<br>Time to diagnosis<br>Diagnosis<br>Diagnostic tests<br>Onward referrals | QDU1 (**Hospital Plató**) vs QDU2 (Hospital Clínic) | Potential QDU1 and 2 data overlap with Bosch 2021 (i.e patient data between 2009 and 2016) – note that cases were chosen randomly |
| Bosch et al (2018). Time to diagnosis and associated costs of an outpatient vs inpatient setting in the diagnosis of lymphoma: a retrospective study of a large cohort of major lymphoma subtypes in Spain BMC cancer, 18(1), pp.1-15. | 2006 to 2016 | Aged ≥18 years with lymphoma (4 types)<br>Sample size: 1,779 patients. 1,184 outpatients (688 from QDU1 and 496 from QDU 2) and 535 inpatients | Patient characteristics<br>Referral source<br>Time to first visit<br>Number of visits<br>Time to examination<br>Time to diagnosis<br>Diagnostic tests<br>Diagnosis<br>Onward referral<br>Mean cost per hospital stay<br>Mean cost per visit<br>Mean cost per patient<br>Cost of diagnostic tests<br>Cost saving from hospitalisation<br>Personnel costs | QDU1 (Hospital Clínic) vs inpatient wards (within Hospital Clínic) vs QDU2 of **hospital of Bellvitge** | Potential QDU1 data overlap with Bosch 2021 (i.e lymphoma data only between 2009 and 2016) |
| Brito-Zerón et al. (2014). Diagnosing unexplained fever: can quick diagnosis units replace inpatient hospitalization? European Journal of Clinical | 2008 to 2011 | All consecutive patients referred to the QDU between November 2008 and April 2011 due to undiagnosed fever<br>Sample size: 344 patients, 176 from QDU and 168 controls (internal medicine department) | Participant characteristics<br>Referral source<br>Referral reason<br>Number of visits<br>Time to diagnosis<br>Diagnosis<br>Length of hospital stay | QDU (Hospital Clínic) vs internal medicine department (unclear if same wards as in Bosch 2020 and Bosch 2018) | Potential QDU data overlap with Bosch 2021, i.e FUN data only between 2009 and 2011 |
| Investigation, 44(8), pp.707-718. |  |  | Diagnostic tests<br>Death during evaluation<br>Onward referrals<br>Mean cost per hospital stay<br>Mean cost per visit<br>Mean cost per patient |  |  |
| Bosch, Jordán and López-Soto (2013). Quick diagnosis units: avoiding referrals from primary care to the ED and hospitalizations The American Journal of Emergency Medicine, 31(1), pp.114-123 | 2006 to 2012 | Consecutive patients attending the QDU and patients admitted to the internal medicine service (randomly chosen) between September 2006 and January 2012<br>Sample size: 4,170 QDU patients and 3.030 hospitalised patients | Participant characteristics<br>Referral source<br>Referral reason<br>Referral appropriateness<br>Time to first visit<br>Number of visits<br>Referral patterns over time<br>Time to diagnosis<br>Diagnosis<br>Length of hospital stay<br>Death during evaluation<br>Onward referrals<br>Mean cost per hospital stay<br>Mean cost per process<br>Mean cost per visit<br>Patient preferences | QDU patients (Hospital Clínic) vs patients admitted to the internal medicine service | QDU data overlap with Bosch 2012a (2,000 consecutive patients evaluated between 2007 and 2010) |
| Bosch et al (2012a). Quick diagnosis units versus hospitalization for the diagnosis of potentially severe diseases in Spain Journal of Hospital Medicine, 7(1), pp.41-47 | 2006 to 2010 | Consecutive QDU patients evaluated between December 2007 and July 2010 and patients diagnosed with anaemia (n = 548), cachexia-anorexia syndrome (n = 458), febrile syndrome (n = 240), and adenopathies or palpable masses (n = 208) admitted to the internal medicine department between September 2006 and June 2010. Sample size: 2,000 QDU patients and 1,454 patients admitted to the internal medicine department | Participant characteristics<br>Referral source<br>Referral reason<br>Time to first visit<br>Number of visits<br>Time to diagnosis<br>Diagnosis<br>Length of hospital stay<br>Charlson comorbidity index<br>Hospital bed days saved<br>Onward referrals<br>Cost per hospital stay<br>Cost per process<br>Cost per visit<br>Patient satisfaction<br>Patient preferences | QDU patients vs patients admitted to the internal medicine department | QDU data overlap with Bosch, Jordán and López-Soto 2013 (2,000 consecutive patients evaluated between 2007 and 2010) |
| Bosch et al (2012b). Comparison of Quick Diagnosis Units and Conventional Hospitalization for the Diagnosis of Cancer in Spain: A Descriptive Cohort Study Oncology, 83(5), pp.283-291. | 2008 to 2010 | QDU patients with both initially suspected and ultimately confirmed (pathologically proven) cancer evaluated between November 2008 and April 2010. Also included patients newly diagnosed with cancer admitted to the internal medicine department<br>Sample size: 169 (62.8% of 269 consecutive patients evaluated for inclusion) QDU patients, and 53 Hospitalised patients | Patient characteristics<br>Referral source<br>Reason for referral/consultation<br>Time to first visit<br>Number of visits<br>Time to diagnosis<br>Length of hospital stay<br>Diagnosis<br>Onward referral<br>Mean cost per hospital stay<br>Mean cost per visit<br>Mean cost per process | QDU (Hospital Clínic) vs internal medicine department | potential QDU data overlap with above studies by Bosch et al. |
| Bosch et al (2012c). Quick diagnosis units or conventional hospitalisation for the diagnostic evaluation of severe anaemia: A paradigm shift in public health systems? European Journal of Internal Medicine, 23(2), pp.159-164. | 2006 to 2010 | Consecutive patients with severe anaemia attended by the QDU and consecutive patients with the same diagnosis admitted to the internal medicine department<br>Sample size: 282 cases from QDU, 252 hospitalised patients | Participant characteristics<br>Referral source<br>Referral reason<br>Time to first visit<br>Number of visits<br>Time to examination<br>Time to diagnosis<br>Diagnosis<br>Length of hospital stay<br>Charlson morbidity index<br>Diagnostic tests<br>Treatment received at unit<br>Onward referrals<br>Mean cost per hospital stay<br>Mean cost per process<br>Mean cost per visit<br>Patient satisfaction<br>Patient preferences | QDU (Hospital Clínic) vs internal medicine department | Likely overlap of patient data with above studies |
| Bosch et al (2011). Outpatient Quick Diagnosis Units for the evaluation of suspected severe diseases: an observational, descriptive | 2008 to 2010 | Consecutive patients evaluated in the QDU<br>Sample size: 1,000 QDU patients and ?150 patients admitted to internal medicine department | Participant characteristics<br>Referral source<br>Referral reason<br>Referral appropriateness<br>Time to first visit<br>Number of visits | QDU (Hospital Clínic) vs internal medicine department | Likely overlap of patient data with above studies |
| study. Clinics, 66(5), pp.737-741. |  |  | Time to diagnosis<br>Diagnosis<br>Length of hospital stay<br>Diagnostic tests<br>Hospital bed days saved<br>Onward referral<br>Mean cost per hospital stay<br>Mean cost per process<br>Mean cost per patient<br>Patient satisfaction<br>Patient preference |  |  |
| **Bellvitge Hospital – Barcelona - described as a 750 bed tertiary hospital with a reference population of around 350,000** |  |  |  |  |  |
| Sanclemente-Ansó et al (2016). Cost-minimization analysis favors outpatient quick diagnosis unit over hospitalization for the diagnosis of potentially serious diseases. Eur J Intern Med; 30:11-17. doi: 10.1016/j.ejim.2015.12.015 | 2008 to 2012 | Patients diagnosed with severe anaemia, lymphoma, and lung cancer selected from consecutive patients referred to the unit between March 2008 and June 2012 and those electively hospitalised at the Internal Medicine Department of the hospital for diagnostic workup during the same period and who had the same final diagnoses. Sample size: 195 QDU patients and 237 Internal Medicine department patients | Referral source<br>Number of visits<br>Time to diagnosis<br>Diagnosis<br>Length of hospital stay<br>Diagnostic test<br>Mean cost per hospital stay<br>Mean cost per patient<br>Direct and structural costs<br>Indirect costs<br>Personnel costs<br>Diagnostic investigation costs<br>Mean cost saving per patient<br>Overall cost saving<br>Diagnostic investigation cost saving<br>Hospitalisation cost saving<br>Cost saving per patient | QDU (Bellvitge) vs internal medicine department | Potential QDU data overlap with Bosch 2018 (QDU2) |

## APPENDIX 3: Resources searched during Rapid Review Searching

**Table 6:**
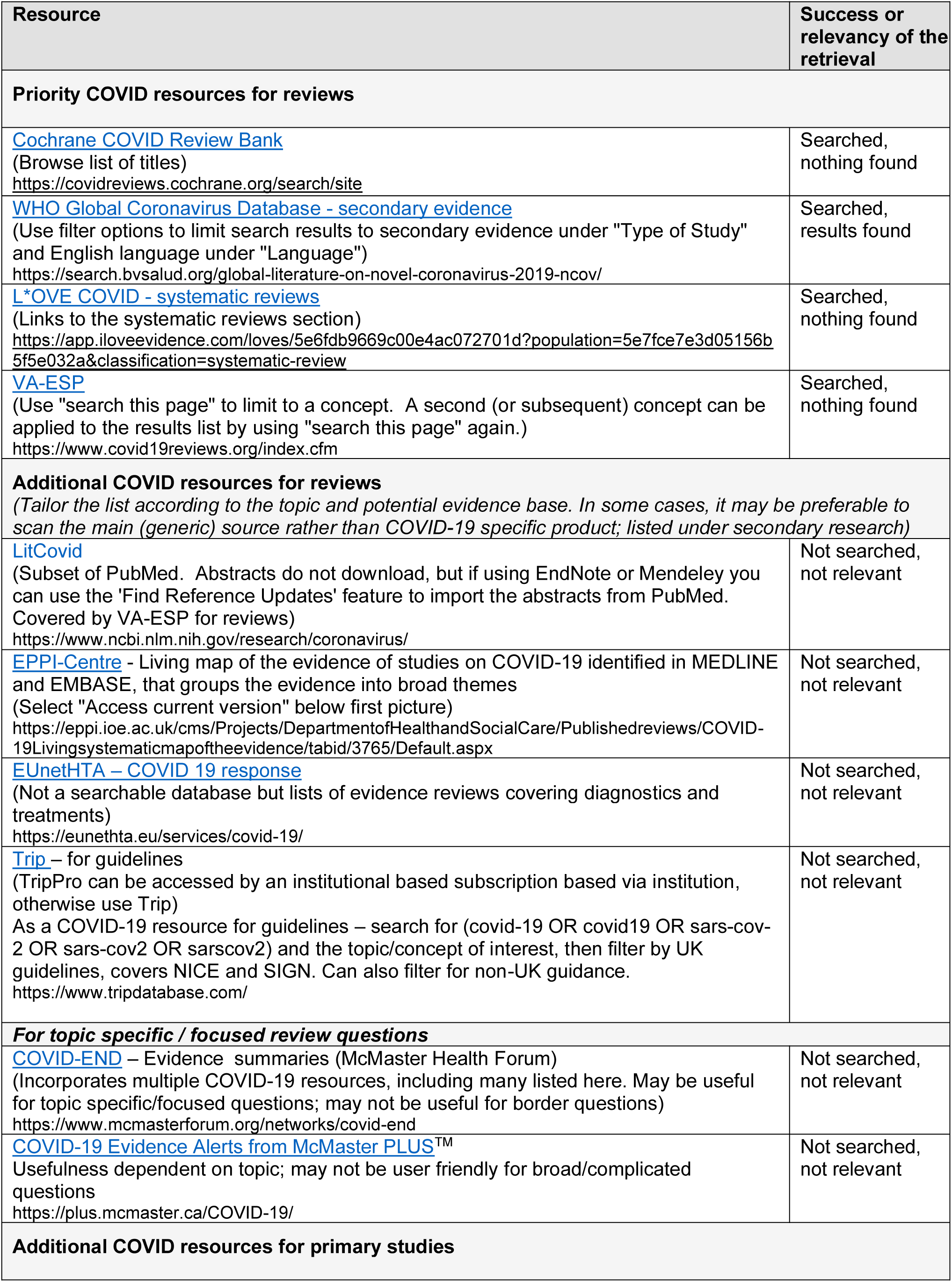
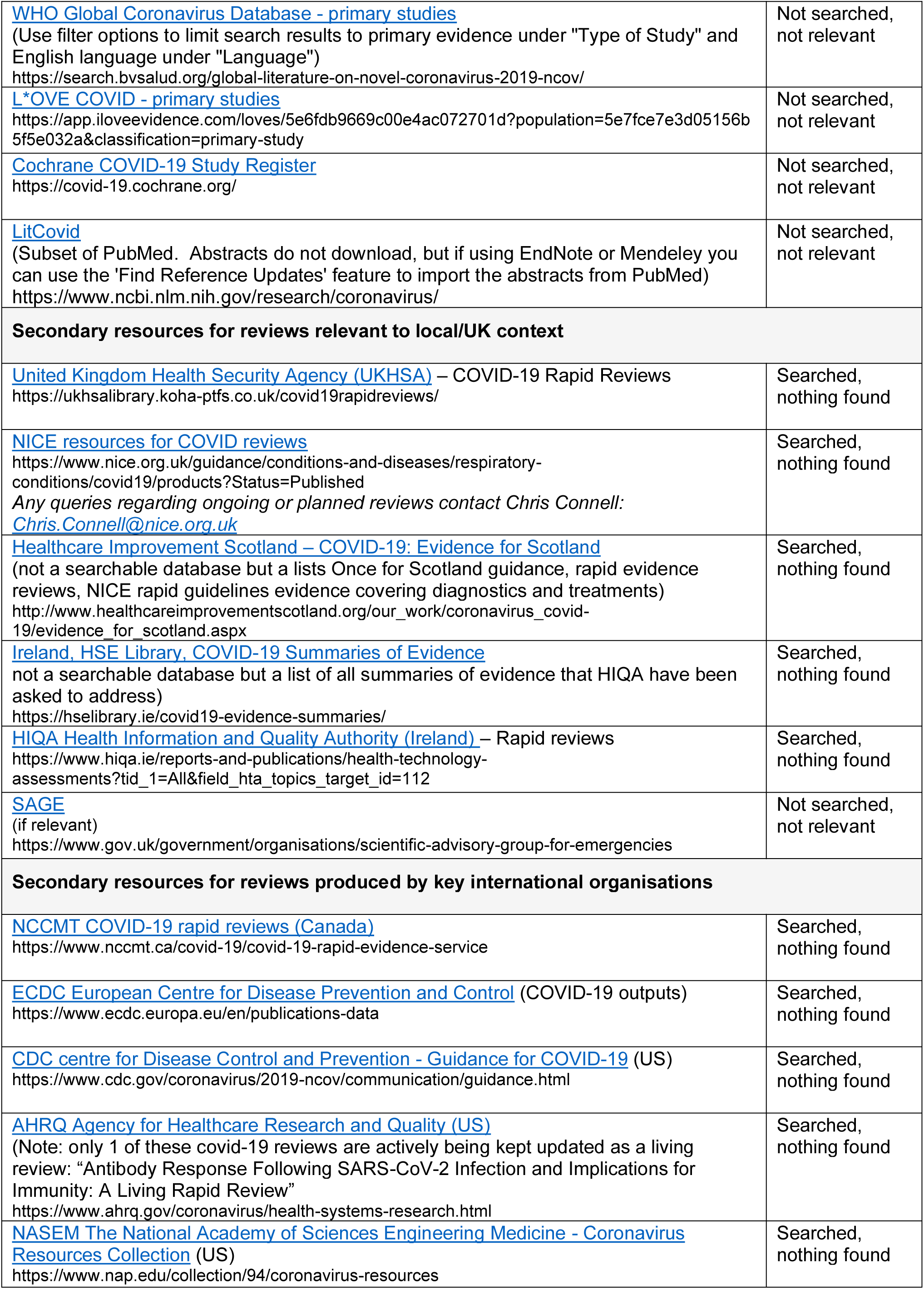
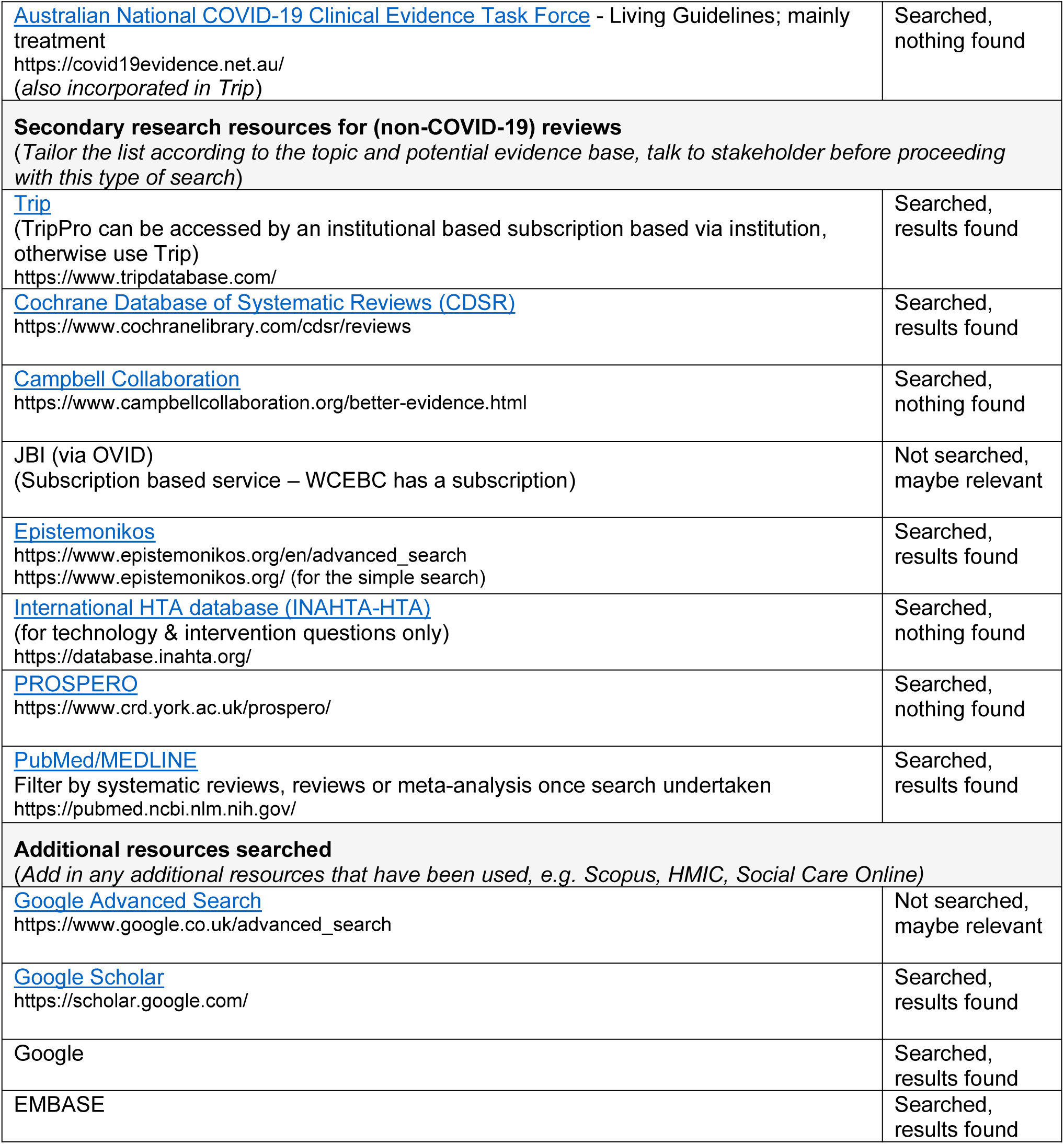
Resources searched

## Appendix 4: Search strategy used for MEDLINE

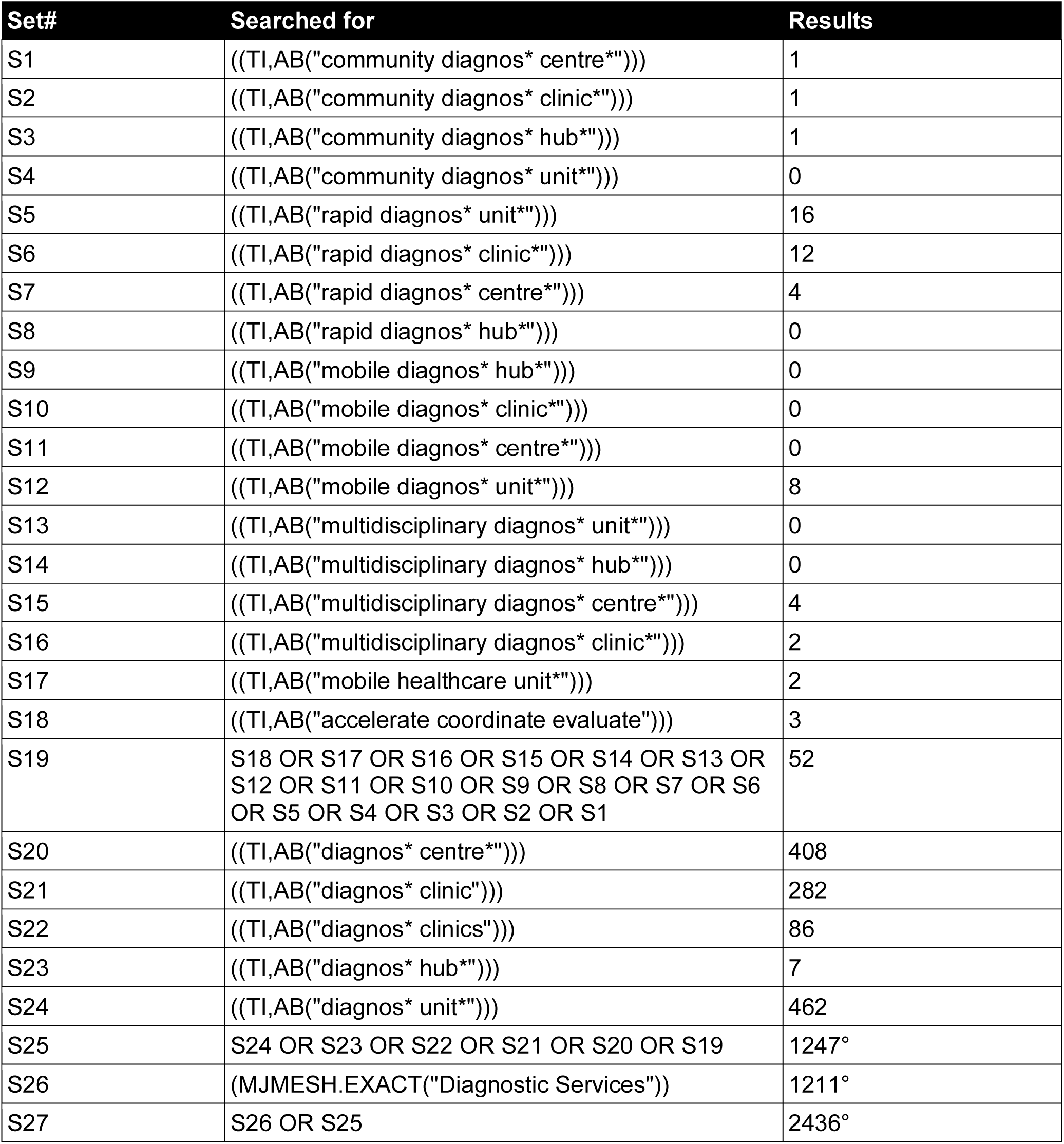

| Set# | Searched for | Results |
| --- | --- | --- |
| S1 | ((TI,AB("community diagnos* centre*"))) | 1 |
| S2 | ((TI,AB("community diagnos* clinic*"))) | 1 |
| S3 | ((TI,AB("community diagnos* hub*"))) | 1 |
| S4 | ((TI,AB("community diagnos* unit*"))) | 0 |
| S5 | ((TI,AB("rapid diagnos* unit*"))) | 16 |
| S6 | ((TI,AB("rapid diagnos* clinic*"))) | 12 |
| S7 | ((TI,AB("rapid diagnos* centre*"))) | 4 |
| S8 | ((TI,AB("rapid diagnos* hub*"))) | 0 |
| S9 | ((TI,AB("mobile diagnos* hub*"))) | 0 |
| S10 | ((TI,AB("mobile diagnos* clinic*"))) | 0 |
| S11 | ((TI,AB("mobile diagnos* centre*"))) | 0 |
| S12 | ((TI,AB("mobile diagnos* unit*"))) | 8 |
| S13 | ((TI,AB("multidisciplinary diagnos* unit*"))) | 0 |
| S14 | ((TI,AB("multidisciplinary diagnos* hub*"))) | 0 |
| S15 | ((TI,AB("multidisciplinary diagnos* centre*"))) | 4 |
| S16 | ((TI,AB("multidisciplinary diagnos* clinic*"))) | 2 |
| S17 | ((TI,AB("mobile healthcare unit*"))) | 2 |
| S18 | ((TI,AB("accelerate coordinate evaluate"))) | 3 |
| S19 | S18 OR S17 OR S16 OR S15 OR S14 OR S13 OR S12 OR S11 OR S10 OR S9 OR S8 OR S7 OR S6 OR S5 OR S4 OR S3 OR S2 OR S1 | 52 |
| S20 | ((TI,AB("diagnos* centre*"))) | 408 |
| S21 | ((TI,AB("diagnos* clinic"))) | 282 |
| S22 | ((TI,AB("diagnos* clinics"))) | 86 |
| S23 | ((TI,AB("diagnos* hub*"))) | 7 |
| S24 | ((TI,AB("diagnos* unit*"))) | 462 |
| S25 | S24 OR S23 OR S22 OR S21 OR S20 OR S19 | 1247° |
| S26 | (MJMESH.EXACT("Diagnostic Services")) | 1211° |
| S27 | S26 OR S25 | 2436° |

## REFERENCES

BBC News (2022) Rapid cancer diagnosis centre rollout in Wales a UK first. Available at: [Rapid cancer diagnosis centre rollout in Wales a UK first - BBC News](https://www.bbc.co.uk/news/uk-wales-63145247) [Accessed 06 October 2022].

Department of Health and Social Care (2021) Available at: 40 community diagnostic centres launching across England - GOV.UK (www.gov.uk). [Accessed 13 July 2022].

Health Education England (2022). ‘Community Diagnostic Centres (CDC)’. Available at: https://www.hee.nhs.uk/our-work/cancer-diagnostics/community-diagnostic-centres-cdc. [Accessed 31 October 2022].

Jensen H, Tørring M.L, Olesen F, et al. (2015). Diagnostic intervals before and after implementation of cancer patient pathways–a GP survey and registry based comparison of three cohorts of cancer patients. BMC cancer, 15(1), pp.1–10.

Leatherdale S.T. (2019) Natural experiment methodology for research: a review of how different methods can support real-world research, International Journal of Social Research Methodology, 22:1, 19–35, DOI: 10.1080/13645579.2018.1488449.

NHS England (2020) Diagnostic: Recovery and Renewal. Report of the Independent Review of Diagnostic Services for NHS England. Available at: https://www.england.nhs.uk/wp-content/uploads/2020/11/diagnostics-recovery-and-renewal-independent-review-of-diagnostic-services-for-nhs-england-2.pdf [Accessed 12 July 2022].

NHS England (2022) One million checks delivered by NHS ‘one stop shops’. News. Available at: [NHS England » One million checks delivered by NHS ‘one stop shops’](https://www.england.nhs.uk/2022/06/one-million-checks-delivered-by-nhs-one-stop-shops/) [Accessed 12 July 2022].

NHS (2022) Document 3 - Community Diagnostic Hub (CDH) Draft Qualification Specification. Available at: https://www.ardengemcsu.nhs.uk/media/2585/document-3-cdh-framework-specification-v111.pdf [Accessed 12 July 2022].

The King’s Fund (2022). ‘Are community diagnostic centres really moving care closer to home?’ Available at: https://www.kingsfund.org.uk/blog/2022/10/are-community-diagnostic-centres-really-moving-care-closer-home. [Accessed 03 November 2022].

Welsh Government (2021) NHS activity and performance summary: July and August 2021. Statistics. Available at: https://gov.wales/nhs-activity-and-performance-summary-july-and-august-2021-html [Accessed 12 July 2022].

Welsh Government (2022) Our programme for transforming and modernising planned care and reducing waiting lists in Wales. Available at: https://gov.wales/sites/default/files/publications/2022-04/our-programme-for-transforming--and-modernising-planned-care-and-reducing-waiting-lists-in-wales.pdf [Accessed 13 July 2022].

StatsWales (2022) Waiting times by month. Available at: https://statswales.gov.wales/Catalogue/Health-and-Social-Care/NHS-Hospital-Waiting-Times/Diagnostic-and-Therapy-Services/waitingtimes-by-month [Accessed 24 October 2022].

